# Change in irritable bowel syndrome symptom severity among members of a multi-omic digital food-as-medicine program: a retrospective single-arm cohort study with estimates corrected for regression to the mean

**DOI:** 10.64898/2026.10.01.26364558

**Authors:** Inti Pedroso, Shreyas V. Kumbhare, Santosh K. Saravanan, Ronesh Sinha, Daniel E. Almonacid, Ranjan Sinha

## Abstract

**Background:** Real-world evidence on food-as-medicine programs for irritable bowel syndrome (IBS) is scarce, and single-arm evaluations overstate improvement because of regression to the mean (RTM). We estimated change in the IBS Symptom Severity Scale (IBS-SSS, 0-500) in a multi-omics personalized digital health food-as-medicine program, unadjusted and RTM-corrected.

**Methods:** Retrospective single-arm cohort. Of 15,933 members with a valid baseline, 2,254 had a follow-up reading and 1,683 one at least 14 days after baseline. Change at 4, 12 (primary) and 26 weeks was estimated by baseline severity stratum, unadjusted and RTM-corrected by analysis of covariance. A responder fell by at least 50 points (the minimal clinically important difference benchmark). Sensitivity analyses covered equity, inverse-probability weighting and delta-based tipping-point analyses for members without a reading in each landmark window, a negative-control stratum and medication. The association of weight loss and dietary nutrient density with symptom change was evaluated within and between members.

**Results:** At 12 weeks (n = 430) IBS-SSS fell by 43.88 points unadjusted (95% confidence interval [CI] – 52.44 to –35.32; p < 0.001) and by 47.73 RTM-corrected (–55.48 to –39.98; p < 0.001); 720 of 1,339 symptomatic members (53.8%; 95% CI 51.1 to 56.4) fell by at least 50 points, and the proportion of responders rose with baseline severity, from 41.0% in the mild stratum (226/551) and 59.4% in the moderate stratum (336/566) to 71.2% in the severe stratum (158/222), staying at 73% to 76% in the severe stratum at each landmark. Larger improvement in more severe members persisted after RTM correction (severe stratum, n = 37: unadjusted –127.43, RTM-corrected –60.71, 95% CI –97.75 to –23.68, p = 0.001). A higher meal nutrient density was associated with lower IBS-SSS within a member’s own trajectory (p = 0.018) while weight change was not (p = 0.361). Follow-up differed by age, sex, income and ethnicity; subgroup differences in RTM-corrected 12-week change were not significant (age p = 0.458, sex 0.749, income 0.486, ethnicity 0.170). RTM-corrected estimates moved by less than 2 points with weighting for members without a reading, and their change would have had to be 39 to 73 points worse than observed before the mean change reached zero. Baseline medication use and starting a GLP-1 receptor agonist did not explain symptom changes.

**Conclusions:** IBS-SSS fell significantly among program participants, with steeper improvement in the more severe group, even after RTM correction. Improvement occurred in all subgroups, with mostly non-significant differences, though follow-up differed by subgroup and the ethnicity result depended on missing-data handling; overall estimates were robust to the assumptions evaluated. These associations from a self-selected single-arm cohort warrant replication in independent studies.

## Background

Irritable bowel syndrome (IBS) is a disorder of gut-brain interaction [1,2] that affects roughly one in nine people worldwide [3,4], impairs health-related quality of life, raises direct and indirect costs [5,6] and is more prevalent in women [7,8]. Obesity is associated with gastrointestinal symptoms in general [9,10], and a conservative weight-loss intervention relieved bowel symptoms in people with morbid obesity [11].

Diet is the main self-management lever, and better dietary interventions are needed. Low-FODMAP (fermentable oligo-, di-, mono-saccharides and polyols) diets reduce symptoms relative to habitual diet [12–15], with a fall of about 45 IBS-SSS points in diet trials that used this scale [16]. Guidance describes staged restriction, reintroduction and personalisation [17,18], and a recent trial reported benefit from a Mediterranean diet as an alternative [19]. Because gut microbiome composition differs between people, diets personalised to microbiome signals are a plausible next step. A multicentre randomised trial compared a microbiome-based, artificial-intelligence-assisted personalised diet with a low-FODMAP diet over 6 weeks and found similar falls in IBS-SSS in both groups (–112.7 versus –99.9 points; between-group p = 0.29) [20]; in an open-label 12-month follow-up of participants who completed the intervention, the personalised-diet group maintained a fall of 78.3 points whereas the low-FODMAP group did not maintain its early benefit (between-group p = 0.001), with 62.5% versus 34.5% responders, clearest in constipation-predominant and mixed IBS and not significant in diarrhoea-predominant IBS, in an analysis the authors describe as hypothesis-generating [21]. Evidence on the effect of personalised dietary guidance on IBS symptoms therefore remains limited. Real-world evidence can fill part of this gap. Systematic reviews of digital interventions for IBS report improvement [22], and real-world single-arm evaluations of digital programs describe the same [23–25], whereas controlled digital trials report smaller between-arm differences [26–28] and a dietary app trial found no significant difference in a small, high-attrition sample [29]. An independent health-technology assessment concluded that wraparound virtual gastrointestinal programs deliver clinically meaningful improvements in IBS symptoms and quality of life, while calling for evidence on the durability of outcomes and on performance in diverse and underserved populations [30].

Importantly, improving patient access to support is a driving motivation for wider and easier access to digital health support for IBS, but the equitability of health outcomes is often not investigated, with the concomitant risk of deepening existing structural health inequalities. Health informatics interventions can generate inequalities when they reach, engage or work better for more advantaged people [31]. Prevalence, care seeking and symptom burden differ by sex, age, body-mass index, income and ethnicity [32,33], and participants in IBS drug trials are predominantly White and female [34]. Digital delivery can widen access, but uptake and use of mobile health vary with age, income and sex [35–37]. A systematic review of ten qualitative studies grouped strategies to advance digital health equity into outreach programs and co-design approaches, and recommended user-friendly and culturally appropriate design, supportive infrastructure and digital-literacy support, while noting that their feasibility and impact still need to be tested in practice [38]. Food-is-medicine programs are proposed as a route to nutrition security and health equity, although most supporting studies are quasi-experimental or pre-post designs [39]. Whether improvement under a digital dietary program differs across demographic groups is therefore an outcome in its own right.

In this study we report health outcomes of a digital health program using multi-omic personalised digital food-as-medicine therapeutics that has been described in earlier studies and shown to improve health outcomes for weight loss, functional gastrointestinal symptoms, gut microbiome composition, mental health symptoms, and reduction in medical claims costs, including digestive disorders [40–44]. The current study presents a single-arm, within-member change in IBS-SSS across baseline severity strata in a contemporary cohort of this program. Our objective is to estimate the unadjusted change alongside an RTM-corrected estimate, and to describe responders, trajectories, equity across demographic groups, robustness to attrition and medication, and the association of weight and diet quality with symptoms.

## Methods

### Study design

We conducted a retrospective, single-arm cohort study of routinely collected app and questionnaire data from members of a commercial digital food-as-medicine program in the United States. The primary outcome was change in IBS-SSS. The report follows the STROBE statement [45] and its RECORD extension [46]. The Institutional Review Board of E&I Review Services reviewed and approved the study protocol (protocol code #18053, approved 05/22/2018). Before being included in the study, all participants provided electronic informed consent as part of their written consent process. Research consent was an inclusion criterion for every analytic population.

### Program

Members were adults eligible for the program if they met any one of the following: a body-mass index (BMI) of 30 or more; a BMI of 25 to below 30 with a cardiometabolic comorbidity; a diagnosis of type 2 diabetes; a self-reported gastrointestinal or digestive condition; self-reported hypertension or high cholesterol; or a current prescription for medication for obesity, diabetes or digestive illness. The program combines app-based tracking, personalised dietary guidance and health coaching, and has been described previously [40–42,44]. Members with IBS, reflux, other functional gastrointestinal disorders or other digestive conditions in which diet is the main treatment follow a digestive-health pathway. They complete the IBS-SSS during onboarding, with a food-trigger history and a symptom diary, and receive an IBS curriculum. A Low FODMAP Trial (elimination, reintroduction, personalisation [17]) is applied via educational material, AI-based recommendations and human coaches’ personalized feedback. Coaching is gastrointestinal-focused, with group coaching sessions and content, and members receive food guides, including a low-FODMAP meal plan. Meal feedback uses a nutrient-density (ND) score (below and Supplementary Methods). Genetic and microbiome personalisation is not evaluated here. Components are delivered together, so none can be credited individually.

### Participants, baseline and flow

Members were eligible if they consented to research use and answered IBS-SSS at least once (all five answered). The index date was the first app download. The baseline was the closest scored reading from 30 days before to 14 days after the index date. This window follows the logic of regulatory guidance for IBS trials, which specifies a baseline measured over a short screening period of one to two weeks before treatment [47,48], widened to the pragmatic scale of routine onboarding, in which the questionnaire may be completed shortly before or after app download. Members whose first scorable reading fell later were considered late entrants and were excluded. A respondent had a valid baseline and at least one follow-up reading of any timing. Readings were read as follows. Follow-up readings less than 14 days after baseline served only to estimate test-retest reliability; readings at 14 days or later (day 14 included) were longitudinal. The longitudinal sample was every respondent with at least one reading at 14 days or later, and it alone defined the centring mean baseline used in the ANCOVA models. The IBS-SSS asks about the previous 10 days [49], so a reading taken less than 14 days after baseline overlaps the baseline recall period, and published retest studies of the scale used comparable intervals [50,51].

### Instruments, strata and covariates

IBS-SSS severity strata were remission (below 75), mild (75-174), moderate (175-299) and severe (300 or more) [49]. Covariates were age, sex, BMI, engagement days (distinct app-activity days in the 90 days after the index date), self-reported ethnicity and household income (with “not answered” retained), and medication.

Meal nutrient density was scored by the program. Members photograph meals in the app, and each meal is scored by a coach or by an artificial-intelligence meal-rating pipeline against three categories of infractions (insulin, inflammation and fibre diversity). A meal starts from a maximum score of 14 and loses one or two points for each matched infraction, so that a higher ND-score is better; the points lost to each category are stored with the meal. For every member-day we derived the ND-score, the share of the day’s scored meals with at least one infraction in each category (penalisation rate) and the mean points lost per scored meal in each category; readings on the same day were averaged, and each IBS-SSS reading was matched to the nearest ND-score reading within 14 days. Details are in the Supplementary Methods.

### Medication sources

Medication information came from two sources. First, the program’s intake form, where members listed medications in structured entries, read as a baseline snapshot. Second, pharmacy claims, which are available for a subset of participants depending on data availability. Claims files were received monthly, parsed with custom scripts following each layout file, matched to members by name and date of birth and de-identified before analysis, and the claims-processing rules of an earlier study were applied [44]. Each drug claim carried the service date and the package national drug code (NDC), which was linked to the openFDA drug-label reference [52,53], downloaded on 10 July 2026, to obtain the product generic and brand names, active ingredients and FDA Established Pharmacologic Class (EPC). A fill counted toward a drug class if the claim product name or the matched openFDA names contained a class keyword (case-insensitive substring), or if the NDC carried the class’s EPC; the same keywords were applied to the intake-form list. Drug classes investigated were antidepressants (selective and serotonin-norepinephrine reuptake inhibitors, tricyclics and others), glucagon-like peptide-1 (GLP-1) receptor agonists and proton-pump inhibitors. Baseline use was a claims fill in the 180 days before baseline or a list entry at baseline. A start was a first fill on or after baseline with none before, assessed only for members whose pharmacy feed covered the period from 180 days before baseline through the last reading. Unobserved medication was counted as not exposed, so that medication analyses describe observed exposure. Keyword and class lists are in the Supplementary Methods.

### Landmarks and estimands

Landmarks were 4, 12 and 26 weeks after baseline (days 14-35, 70-98 and 154-210); the reading closest to day 28, 84 or 182 was used (ties to the earlier). The 12-week landmark was primary. The pooled estimate used each longitudinal member’s first reading at 14 days or later. Change was follow-up minus baseline (negative = improvement). The primary estimand was the unadjusted mean change (mean paired difference with a t interval), always reported beside the RTM-corrected estimate. The RTM-corrected estimate was the change at the mean baseline of the longitudinal sample (171.42) from an ANCOVA mixed model of change on centred baseline, stratum, landmark and their interaction with a member random intercept, fitted by restricted maximum likelihood [54,55] with Satterthwaite degrees of freedom and estimated marginal means [56] (R formula change ∼ centred_baseline + stratum * landmark + (1 | member); centring as in [57–59]), namely

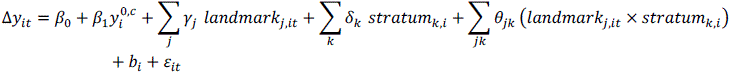

where y^0,*c*^ is the centred baseline score, *b_i_* the member random intercept and all-stratum estimates weight strata by cell size. Pooled estimates used one row per member and ordinary least squares (change ∼ 0 + stratum + centred_baseline), and the heterogeneity of the baseline slope across strata was tested by a nested F test. Because analysis of covariance can differ from change-score analysis outside randomised designs [59], both are shown with 95% CIs and p values. As an independent cross-check, the expected RTM shift is E[X2 – X1] = –(1 – ρ)(xˉbaseline – μ), where ρ is the test-retest reliability of the scale (assumed 0.719) [60–62] and μ the mean baseline of the longitudinal sample; each member’s change was corrected by adding (1 – ρ)(baseline – μ), with a t interval that treats ρ as fixed. Estimates at each stratum’s own mean baseline, a baseline-by-stratum slope test and a negative-control reading of the remission stratum (members near the floor of the scale, expected to show an apparent rise from RTM alone) were sensitivity analyses.

A responder was a symptomatic member (baseline 75 or more) whose score fell by 50 points or more [49]; remission (follow-up score below 75) was a secondary definition. Proportions carry Wilson 95% CIs. Responder and remission proportions were also computed within baseline severity strata (mild 75 to 174, moderate 175 to 299, severe 300 or above) and for the moderate and severe strata combined, at each landmark; these were descriptive, unadjusted and not tested.

### Sensitivity, trajectories, equity and correlates

Respondents were compared with non-respondents, and longitudinal with reliability-only respondents, by standardised mean differences of the baseline characteristics. In a drop-one analysis, age, sex, BMI or the baseline term was removed in turn on a fixed complete-case sample (raw_change ∼ baseline_c + age_c + sex + bmi_c, ordinary least squares). Missing landmark readings were examined within each landmark among the 1,683 longitudinal members: a member was observed at a landmark if they had a reading in its window and missing if they had a reading at 14 days or later but none in that window. Inverse-probability weights [63] came from a logistic model of being observed (observed ∼ baseline_score + age + bmi + sex, weight set A, or with log(1 + engagement days before the baseline reading) added, weight set B), were stabilised and truncated at the 1st and 99th percentiles (a third set, C, added log(1 + distinct app-activity days in the first 14 days after the baseline reading) to set B, as a sensitivity analysis because early engagement is measured after baseline), and were applied to the observed members in a weighted least squares regression of follow-up on centred baseline (followup ∼ I(baseline – 171.42)) with heteroscedasticity-robust (HC1) intervals; for comparison, the same weighting was applied to dropout after baseline among all 15,933 in-window members (IPW-A, responding at all; IPW-B, membership of the longitudinal sample). A delta-based tipping-point sensitivity analysis [64] for data missing not at random imputed the change of the missing members as the observed mean plus a fixed delta; we report the delta at which the mean change over all 1,683 members reached zero and compare it with the 50-point benchmark. Medication terms were added to the RTM-corrected model (change ∼ centred_baseline + stratum * landmark + medication terms + (1 | member)), and, because GLP-1 receptor agonists cause gastrointestinal adverse events [65,66], GLP-1 starters were compared with non-starters on RTM-corrected change (change ∼ 0 + stratum + centred_baseline + starter) and on worsening of 50 points or more (Fisher test). Because the baseline reading could be taken up to 14 days after the index date, the baseline offset from the index date was described among the longitudinal members, and the 4-, 12- and 26-week estimates were re-estimated in members whose baseline was on or before the index date (0 days or less) and in those whose baseline was at most 7 days after it. Landmark windows were defined from each member’s own baseline reading, as in the primary analysis; the RTM-corrected estimates used the same model as the primary analysis, centred at the full longitudinal-sample mean (171.42), and estimates recentred at the mean of each restricted sample were computed as a further sensitivity analysis. Trajectories used a stratum-by-monthly-bin Gaussian mixed model for the unadjusted estimates (glmmTMB [67]; score ∼ stratum * bin + diag(0 + stratum | member), dispersion varying by stratum and baseline indicator), a linear mixed model for the RTM-corrected estimates (change ∼ centred_baseline + stratum * bin + (1 | member), follow-up readings only), and smooth additive mixed models (score ∼ stratum + s(time, by = stratum) with a member random intercept, fitted with gamm4 on mgcv [68–70]), each unadjusted and RTM-corrected. For equity, change was modelled as centred baseline plus a subgroup term by least squares at each landmark (change ∼ 0 + subgroup + centred_baseline), separately for age, BMI, sex, income and ethnicity, with an omnibus F test; the equity models were repeated with the landmark weights (sets A and B) among members with complete age, BMI and sex, by weighted least squares with HC1 intervals, with the difference between weighted and unweighted estimates given an interval from their joint influence function; follow-up by subgroup was described among all 15,933 members with a valid baseline (rates with Wilson intervals and Pearson chi-square tests across strata); baseline severity and stratum mix by subgroup were tested by analysis of variance and Monte Carlo chi-square. Weight and ND-score were related to IBS-SSS by a within/between decomposition [71] (ibs_sss ∼ baseline_ibs_sss + time_c + x_within + x_between + (1 | member)) with cluster-robust variance [72,73] and Satterthwaite degrees of freedom [74], with a sensitivity analysis adding member-level covariates. In an exploratory item analysis, each IBS-SSS item (0-100) and a pain domain (severity plus frequency, 0-200) was analysed as the total score, with landmark change unadjusted and RTM-corrected (each item’s own baseline as the covariate, centred at its longitudinal-sample mean), continuous-time models (monthly-bin and smooth) in the longitudinal sample, and a stacked item-by-time test of whether profiles (change as a percentage of each item’s baseline mean) differed between items, overall and within each baseline severity stratum (Supplementary Methods). All analyses were run in R 4.6.1 [75]; the pooled, equity, weighting and reliability analyses used ordinary least squares and related routines written in Python (version 3.12 or later). Full formulas are in the Supplementary Methods. All analyses were exploratory, without multiplicity correction; counts are exact, and no causal inference is warranted.

## Results

### Cohort

Of 16,250 research-consented participants who answered the IBS-SSS at least once, 317 were late entrants, leaving 15,933; 13,679 had no follow-up reading, leaving 2,254 respondents (14.15%; 95% CI 13.61 to 14.70) (Figure 1). The longitudinal sample had 1,683 members and 571 respondents had reliability readings only; 787 members had a reading under 14 days (216 also had one at 14 days or later). Landmark samples were 337 (4 weeks), 430 (12 weeks) and 345 (26 weeks). Among 12,852 symptomatic members with a valid baseline, 1,812 were respondents and 1,339 were in the longitudinal sample.

**FIGURE 1.**
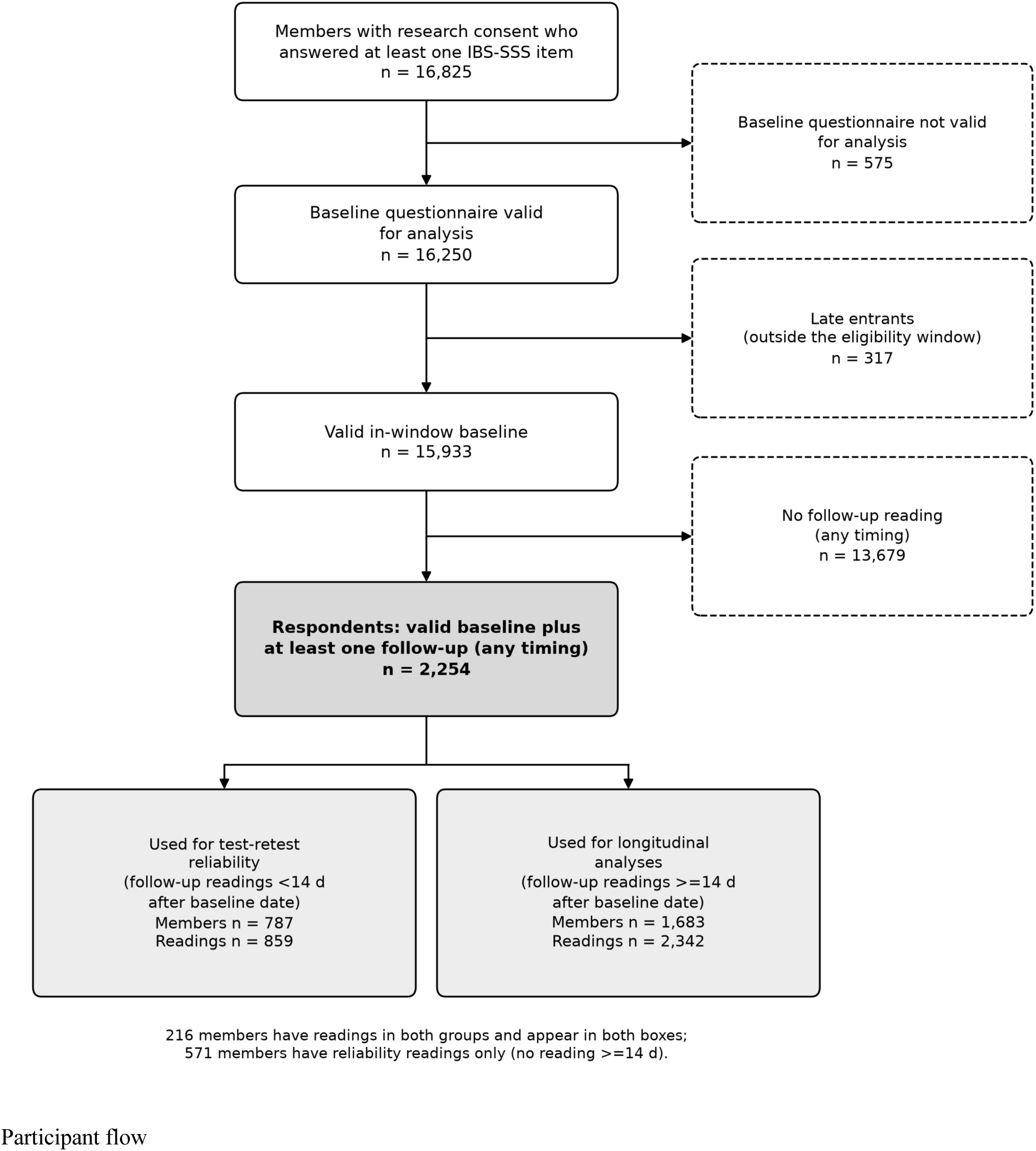
Participant flow. Of 16,250 research-consented participants who answered the IBS-SSS (all five items) at least once, 317 were excluded as late entrants and 13,679 for having no follow-up reading of any timing, leaving 2,254 respondents. Follow-up readings under 14 days were used for test-retest reliability (787 members, 859 readings) and readings of 14 days or more for longitudinal analyses (1,683 members, 2,342 readings). Of these, 216 members had readings in both groups and appear in both boxes; 571 members had reliability readings only.

Respondents were mostly women with a mean body-mass index of 34 (Table 1; full table in Supplementary Table S1); women and younger members were over-represented in the more severe strata, and older members and men in remission (Figure 2). At 12 weeks, baseline severity differed by age (p = 0.002) and sex (p = 0.006): 42% of members aged 65 or older were in remission and 3% severe, and 38% of men were in remission versus 22% of women. The stratum mix at 12 weeks was borderline for age (Monte Carlo p = 0.074) and sex (p = 0.082) and not different for BMI (p = 0.465), income (p = 0.616) or ethnicity (p = 0.159); in the longitudinal sample it differed by age and sex (both p < 0.001; BMI 0.310, income 0.059, ethnicity 0.077). The follow-up rate ranged from 13.74% to 15.71% across strata (Supplementary Table S2). Respondents differed from non-respondents in engagement (32.59 versus 9.57 days; standardised mean difference +1.03), in not answering ethnicity (2.8% versus 28.4%; –0.75) and income (–0.77), and in age (+0.31), but not in baseline score (175.41 versus 171.73; +0.04) (Supplementary Table S3). The longitudinal sample had lower baseline scores (171.42 versus 187.19; – 0.15) and higher engagement (34.98 versus 25.56 days; +0.36) than reliability-only respondents (Supplementary Table S4). Medication data were sparse and not uniformly available: 31.9% of the longitudinal sample linked to pharmacy claims and 27.0% had pharmacy rows, whereas 81.2% had an intake medication list (Supplementary Table S5; Supplementary Figure S1).

**FIGURE 2.**
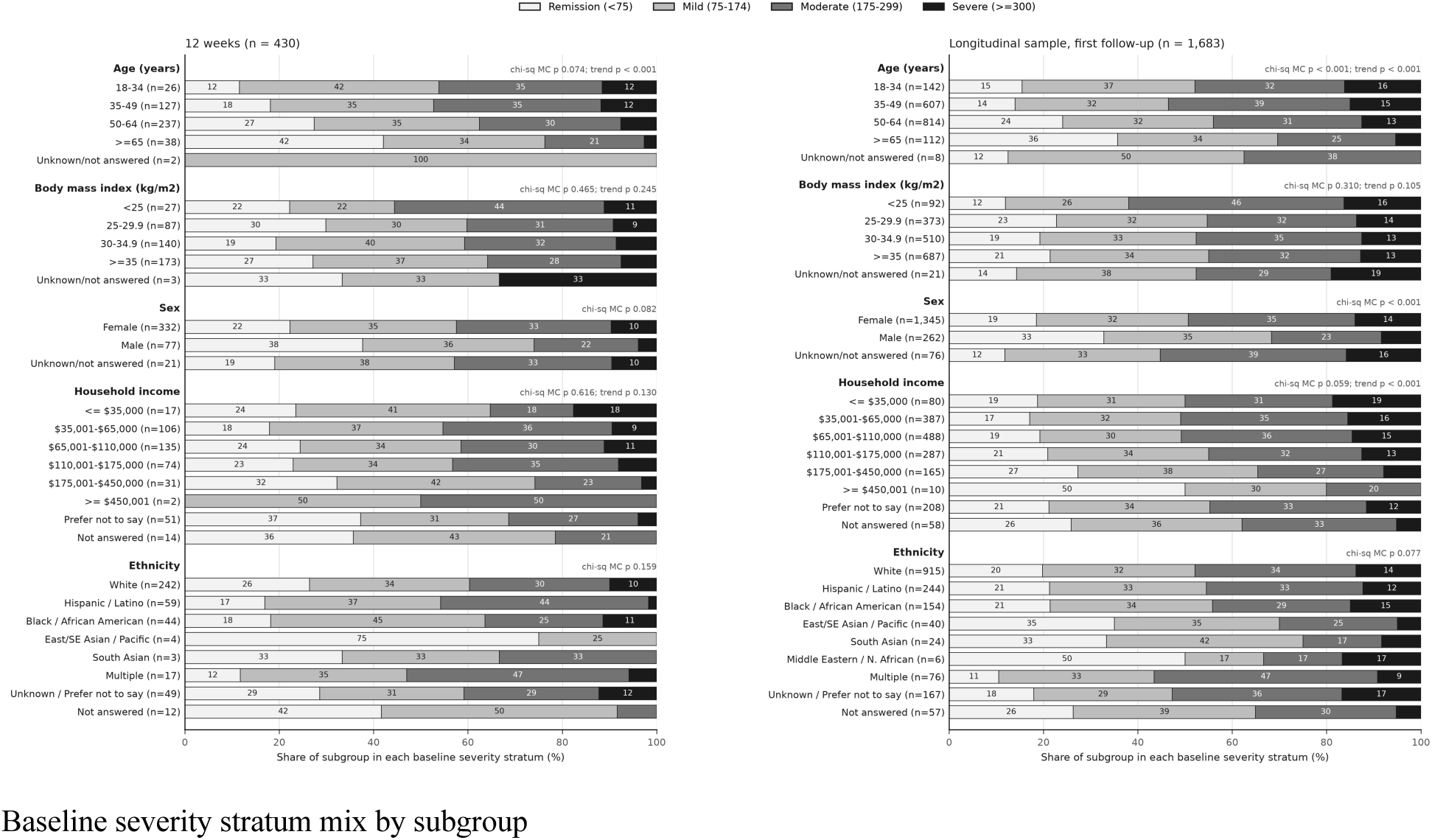
Baseline severity-stratum mix within each subgroup. Each bar is 100% of the subgroup at 12 weeks (n = 430, left) and in the longitudinal sample (n = 1,683, right); numbers are percentages. P values above each factor are Monte Carlo chi-square tests of subgroup by severity stratum and, for ordered factors, linear trend tests. Exploratory; no multiplicity correction.

**TABLE 1.** Baseline characteristics of respondents by baseline IBS-SSS stratum. Cells are n (%) or mean (SD). Medication rows use the headline definition (intake list or pharmacy claims, through follow-up); definitions and the detailed medication and claims breakdown are in Supplementary Tables S1 and S5.

| Characteristic | All respondents (n = 2,254) | Remission, <75 (n = 442) | Mild, 75-174 (n = 733) | Moderate, 175-299 (n = 762) | Severe, ≥300 (n = 317) |
| --- | --- | --- | --- | --- | --- |
| Longitudinal sample, n (%) | 1,683 (74.7%) | 344 (77.8%) | 551 (75.2%) | 566 (74.3%) | 222 (70.0%) |
| Health Status |  |  |  |  |  |
| Baseline IBS-SSS | 175.41 (106.00) | 34.60 (23.14) | 127.09 (27.91) | 228.80 (35.78) | 355.16 (45.11) |
| Body-mass index, kg/m<sup>2</sup> | 34.38 (7.19) | 34.69 (7.38) | 34.71 (7.11) | 34.14 (7.13) | 33.76 (7.17) |
| Demography |  |  |  |  |  |
| Age, years | 49.62 (10.68) | 52.81 (10.43) | 49.67 (10.89) | 48.85 (10.41) | 46.93 (10.18) |
| Female, n (%) | 1,798 (79.8%) | 313 (70.8%) | 577 (78.7%) | 636 (83.5%) | 272 (85.8%) |
| Ethnicity |  |  |  |  |  |
| White, n (%) | 1,239 (55.0%) | 240 (54.3%) | 401 (54.7%) | 414 (54.3%) | 184 (58.0%) |
| Hispanic or Latino, n (%) | 329 (14.6%) | 67 (15.2%) | 107 (14.6%) | 114 (15.0%) | 41 (12.9%) |
| Black or African American, n (%) | 202 (9.0%) | 40 (9.1%) | 70 (9.6%) | 61 (8.0%) | 31 (9.8%) |
| Engagement |  |  |  |  |  |

| **Characteristic** | **All respondents (n = 2,254)** | **Remission, <75 (n = 442)** | **Mild, 75-174 (n = 733)** | **Moderate, 175-299 (n = 762)** | **Severe, ≥300 (n = 317)** |
| --- | --- | --- | --- | --- | --- |
| Engagement, app-activity days in the 90 days after the index date, mean (SD) | 32.59 (27.51) | 35.74 (28.28) | 32.10 (27.34) | 32.17 (27.69) | 30.35 (26.11) |
| Median day of first follow-up reading | 44 | 91 | 60 | 33 | 19 |
| Medication |  |  |  |  |  |
| Proton-pump inhibitor use, n (%) | 327 (14.5%) | 50 (11.3%) | 98 (13.4%) | 117 (15.3%) | 62 (19.6%) |
| GLP-1 receptor agonist use, n (%) | 440 (19.5%) | 86 (19.5%) | 150 (20.5%) | 149 (19.6%) | 55 (17.4%) |
| Linked to claims data, n (%) | 686 (30.4%) | 127 (28.7%) | 217 (29.6%) | 237 (31.1%) | 105 (33.1%) |

### Test-retest reliability

Among 787 members with a first follow-up reading under 14 days (median gap 4 days), the intraclass correlation ICC(2,1) was 0.709 (bootstrap 95% CI 0.668 to 0.748) and Pearson r 0.729 (0.694 to 0.760). The mean difference was –25.71 points (–31.27 to –20.14; p < 0.001) with limits of agreement –181.67 to 130.25 (Supplementary Figure S2).

### Responders

In the longitudinal sample, 720 of 1,339 symptomatic members (53.8%; 95% CI 51.1 to 56.4) fell by at least 50 points. By landmark the proportions were 49.8% (139/279; 44.0 to 55.7) at 4 weeks, 52.3% (169/323; 46.9 to 57.7) at 12 weeks and 63.0% (172/273; 57.1 to 68.5) at 26 weeks (Supplementary Table S6; Supplementary Figure S3). In the longitudinal sample 371 of 1,339 (27.7%) reached remission (below 75); 712 members improved by at least one severity stratum, 700 stayed in the same stratum and 271 worsened (Supplementary Table S7).

Responder proportions were higher in the more severe strata at every reading set (Supplementary Table S35). Using each member’s first reading at 14 days or later, 226/551 (41.0%; 95% CI 37.0 to 45.2) of the mild stratum, 336/566 (59.4%; 55.3 to 63.3) of the moderate stratum and 158/222 (71.2%; 64.9 to 76.7) of the severe stratum fell by at least 50 points. At 12 weeks the proportions were 41.8% (64/153; 34.3 to 49.8), 57.9% (77/133; 49.4 to 65.9) and 75.7% (28/37; 59.9 to 86.6). The mild stratum rose from 33.3% (31/93; 24.6 to 43.4) at 4 weeks to 49.2% (58/118; 40.3 to 58.1) at 26 weeks and the moderate stratum from 51.5% (67/130; 43.0 to 60.0) to 73.5% (86/117; 64.9 to 80.7), while the severe stratum stayed near 73% to 76% (73.2%, 75.7% and 73.7% at 4, 12 and 26 weeks; 41/56, 28/37 and 28/38), with wide intervals because the stratum is small at the later landmarks. Among members with a baseline of 175 or more (moderate and severe combined), 58.1% (108/186; 50.9 to 64.9), 61.8% (105/170; 54.3 to 68.7) and 73.5% (114/155; 66.1 to 79.9) responded at 4, 12 and 26 weeks. The mild stratum had a ceiling effect: more of its members ended below 75, so remission at 12 weeks fell from 43.8% (67/153) in the mild stratum to 22.6% (30/133) in the moderate and 10.8% (4/37) in the severe stratum, and a fall of 50 points was less often reachable from a low baseline. Different members contribute at each landmark, so the percentages are not the trajectory of one group.

### Longitudinal changes in IBS symptom severity

At 12 weeks (n = 430), IBS-SSS fell by 43.88 points unadjusted (95% CI –52.44 to –35.32; p < 0.001) and by 47.73 after RTM correction (–55.48 to –39.98; p < 0.001) (Figure 3). The reliability-formula cross-check for all strata was –48.66 (–56.28 to –41.05), which is consistent with the RTM correction estimate. By stratum, unadjusted change was +9.93 in remission (n = 107; –1.24 to +21.09; p = 0.081), –30.19 in mild (n = 153; –41.78 to –18.61), –79.66 in moderate (n = 133; –96.24 to –63.08) and –127.43 in severe (n = 37; –159.80 to –95.06), all p < 0.001 except remission. RTM-corrected change was –37.49 (–63.57 to –11.42; p = 0.005), –44.85 (–58.80 to –30.89), –55.67 (–71.54 to –39.80) and –60.71 (–97.75 to –23.68; p = 0.001), all p < 0.001 except those two. RTM correction thus halved the fall in the severe stratum and turned the rise in the remission stratum into a fall, but a gradient of larger improvement with higher baseline severity remained.

**FIGURE 3.**
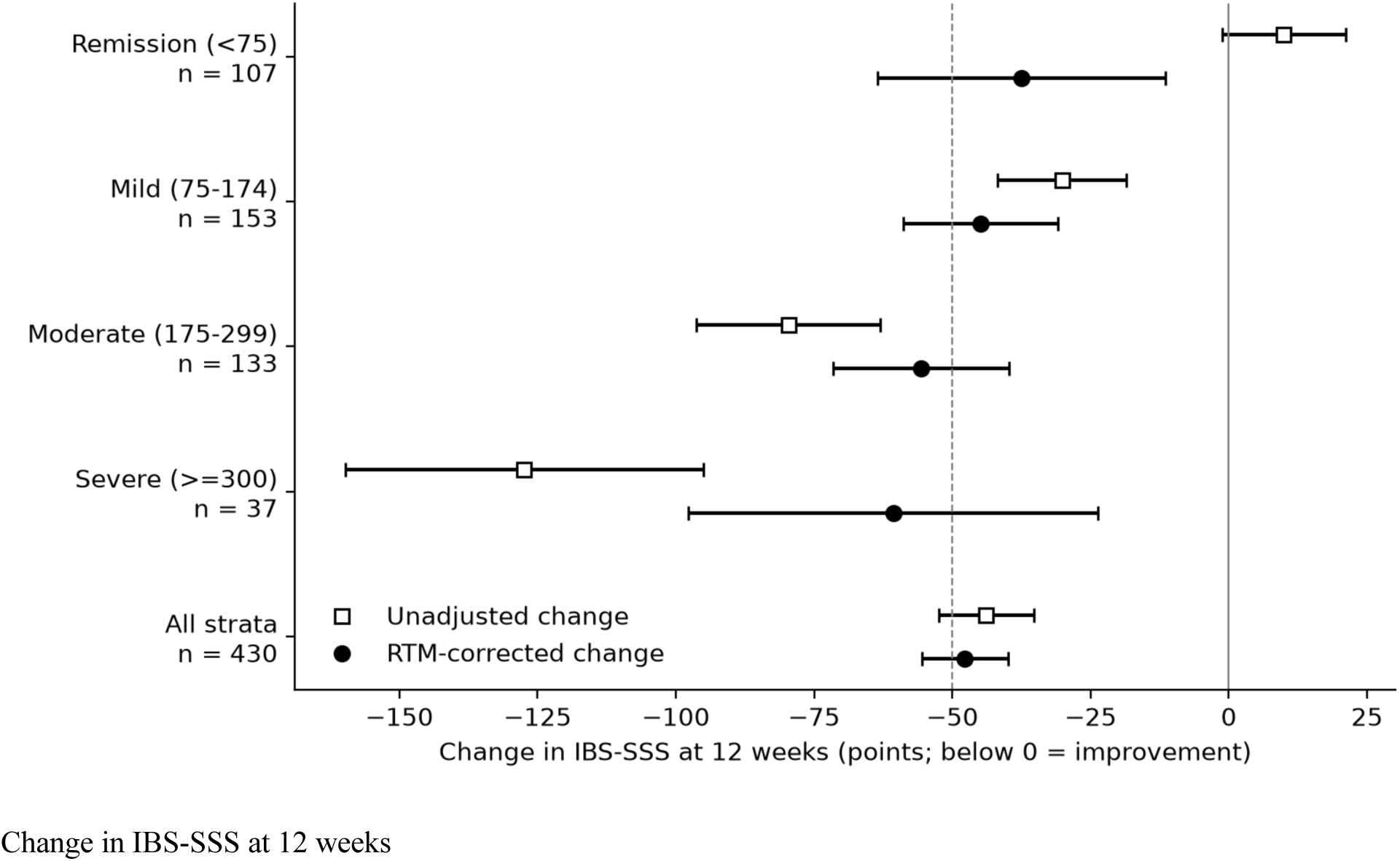
Change in IBS-SSS at 12 weeks by baseline severity stratum. Mean change from baseline with 95% CIs, unadjusted and RTM-corrected, for remission (n = 107), mild (n = 153), moderate (n = 133) and severe (n = 37) strata and for all strata (n = 430). Negative values indicate improvement; the dashed line marks a fall of 50 points, the benchmark for a minimal clinically important difference. RTM-corrected estimates are centred at the mean baseline of the longitudinal sample (171.42). Associations only.

At 4 weeks (n = 337) the change was –39.25 unadjusted (–49.30 to –29.20; p < 0.001) and –33.58 RTM-corrected (–42.41 to –24.75; p < 0.001); at 26 weeks (n = 345) it was –54.69 (–64.06 to –45.32; p < 0.001) and –55.82 (–63.90 to –47.75; p < 0.001). Pooled across the longitudinal sample (n = 1,683) the change was –40.54 unadjusted (–45.12 to –35.96; p < 0.001) and –40.54 RTM-corrected (–44.59 to –36.48; p < 0.001); with a first reading at 30 days or later (n = 1,482) it was –42.32 and –43.72 (Table 2; p values, the reliability-formula cross-check and the 30-day results by stratum in Supplementary Table S8).

**TABLE 2.** Primary results: change in IBS-SSS from baseline (points; negative = improvement), unadjusted and RTM-corrected, by baseline severity stratum at 12 weeks (primary landmark) and overall at each landmark and pooled. RTM-corrected estimates are centred at the mean baseline of the longitudinal sample (171.42). Estimates for strata at 4 and 26 weeks and pooled are in Supplementary Table S8.

| Landmark | Stratum | n | Unadjusted change [95% CI] | RTM-corrected change [95% CI] |
| --- | --- | --- | --- | --- |
| 12 weeks | remission (<75) | 107 | +9.93 [-1.24, +21.09] | -37.49 [-63.57, -11.42] |
| 12 weeks | mild (75-174) | 153 | -30.19 [-41.78, -18.61] | -44.85 [-58.80, -30.89] |
| 12 weeks | moderate (175-299) | 133 | -79.66 [-96.24, -63.08] | -55.67 [-71.54, -39.80] |
| 12 weeks | severe (≥300) | 37 | -127.43 [-159.80, -95.06] | -60.71 [-97.75, -23.68] |
| 12 weeks | all strata | 430 | -43.88 [-52.44, -35.32] | -47.73 [-55.48, -39.98] |
| 4 weeks | all strata | 337 | -39.25 [-49.30, -29.20] | -33.58 [-42.41, -24.75] |
| 26 weeks | all strata | 345 | -54.69 [-64.06, -45.32] | -55.82 [-63.90, -47.75] |
| Pooled, first reading at 14 days or later | all strata | 1,683 | -40.54 [-45.12, -35.96] | -40.54 [-44.59, -36.48] |
| Pooled, first reading at 30 days or later | all strata | 1,482 | -42.32 [-47.26, -37.37] | -43.72 [-48.09, -39.35] |

The empirical slope of follow-up on baseline was 0.577 (95% CI 0.538 to 0.616) pooled and 0.555 (0.479 to 0.630) at 12 weeks, so neither interval contained the assumed retest reliability of 0.719, indicating more regression than the formula assumes. We evaluated if the ANCOVA RTM model with a single baseline mean model could be improved by using strata specific baseline mean, which may suggest differential response by baseline severity level. The baseline-by-stratum slope differed at 12 weeks (F(3, 422) = 2.869, p = 0.036) but not at 4 weeks (p = 0.647), 26 weeks (p = 0.304) or pooled (p = 0.105). Estimates at each stratum’s own mean baseline are in Supplementary Table S9.

### Longitudinal IBS symptoms trajectories

The unadjusted monthly-bin model used 2,254 respondents (4,596 rows) and the RTM-corrected model 1,683 members (2,342 follow-up rows). The stratum-by-month interaction was large in the unadjusted model (chi-square(36) = 710.4; p < 0.001; Figure 4) and much smaller after RTM correction (chi-square(33) = 50.9; p = 0.024; Supplementary Figure S4). At 12 weeks the monthly-bin RTM-corrected falls were –34.7 (remission), –39.6 (mild), –51.5 (moderate) and –42.3 (severe), and the smooth additive model gave similar estimates, –24.6, –37.1, –50.9 and –47.0 (Supplementary Table S10; Supplementary Figure S5). The RTM-corrected remission estimates extrapolate beyond that stratum’s baseline range.

**FIGURE 4.**
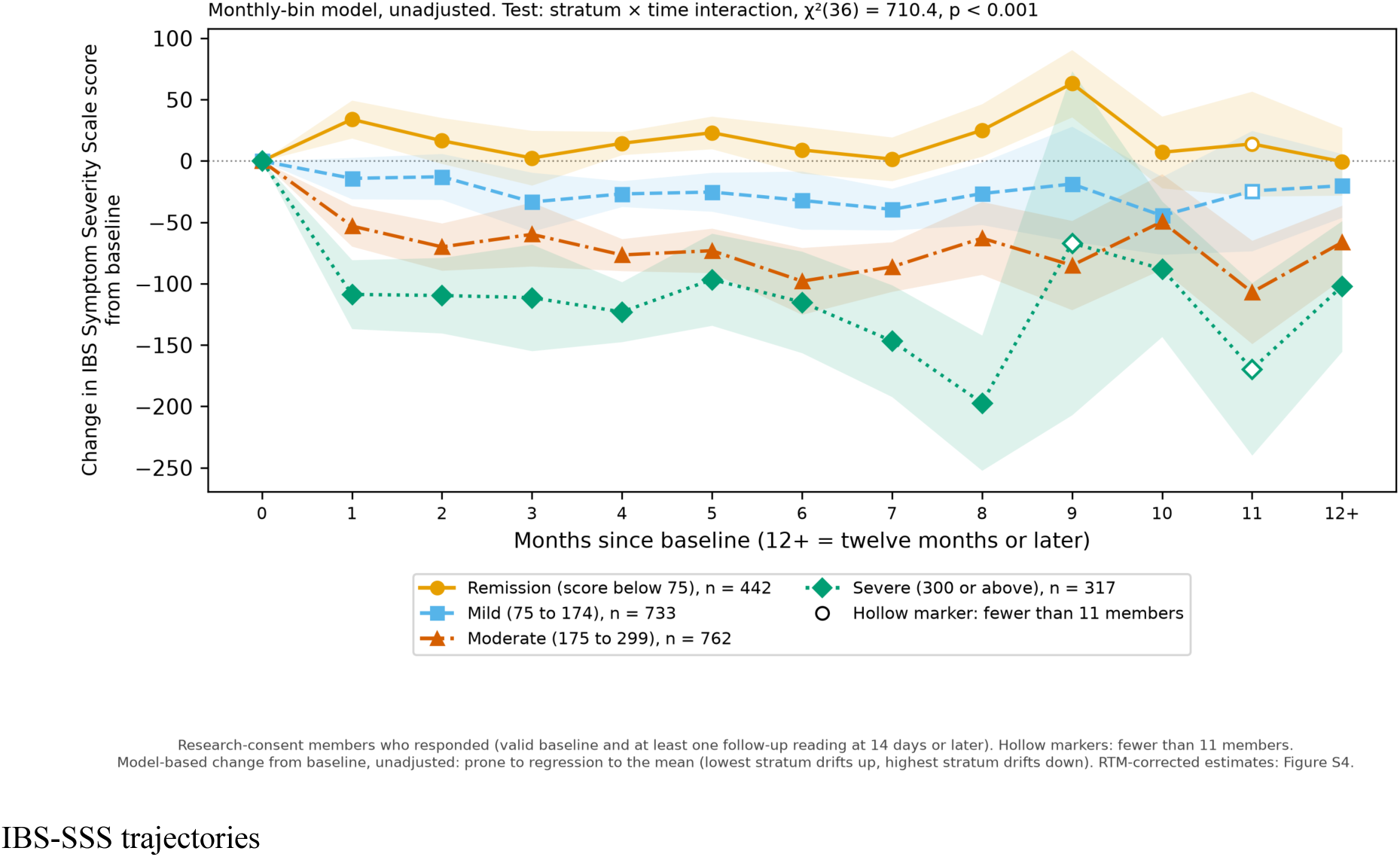
Change in IBS-SSS by baseline severity stratum over 12 months. Unadjusted monthly-bin mixed-model estimates (2,254 respondents, 4,596 rows; stratum-by-time chi-square(36) = 710.4, p < 0.001) with 95% CIs. Hollow markers indicate months with fewer than 11 members. The unadjusted trajectories are prone to regression to the mean; RTM-corrected estimates (1,683 members, 2,342 follow-up rows; chi-square(33) = 50.9, p = 0.024) are in Supplementary Figure S4.

### Equitability of improvements in IBS symptom severity

Baseline severity differed by age and sex, and stratum mix differed by age and sex in the longitudinal sample (Cohort; Figure 2). Follow-up was also uneven (Supplementary Table S33): of 15,933 valid-baseline members, the share with any follow-up reading was 8.6% at 18 to 34 years and 21.4% at 65 or older, 15.8% in women and 9.6% in men, and 1.6% both for members who did not answer income and for those who did not answer ethnicity, and it differed by age, BMI, sex, income and ethnicity (chi-square p < 0.001 for each; the same for membership of the longitudinal sample). For a reading in the 12-week window, rates differed by age, BMI, income and ethnicity (p < 0.001) but not clearly by sex (p = 0.071). Among the 430 members with a 12-week reading, RTM-corrected change did not differ significantly by age (F(4, 424) = 0.91, p = 0.458), BMI (p = 0.328), sex (p = 0.749) or household income (p = 0.486), and the ethnicity test was not significant in all 430 (p = 0.170) (Figure 5; Supplementary Table S11) but was sensitive to the handling of members with missing covariates: among the 407 members with complete age, BMI and sex it was p = 0.033 unweighted and p = 0.023 with inverse-probability weights (set A; set B 0.025) (Supplementary Table S34). The equity model has no stratum terms, so its overall RTM-corrected change is –51.47 (–59.04 to –43.89; unadjusted –43.88) rather than –47.73. Of 29 strata, five had a lower baseline but did not differ detectably in change, one (Hispanic or Latino members, –74.7) did not differ detectably in baseline but differed in change, and 23 did not differ detectably on either. At other landmarks the pooled RTM-corrected change differed by age (p = 0.002) and sex (p = 0.038), and at 26 weeks by BMI (p = 0.023); all other omnibus tests were not significant (Supplementary Tables S12, S13, S14 and S15; Supplementary Figure S6). The 12-week landmark has 430 members against 1,683 in the pooled sample, so a non-significant omnibus test at 12 weeks and significant tests at other landmarks partly reflect precision. Weighting the 12-week equity analysis for non-response left the subgroup estimates close to the unweighted ones: RTM-corrected differences between weighted and unweighted estimates were between –3.2 and +3.9 points in every stratum with at least 11 members (set A), and the weighted omnibus tests were p = 0.312 for age, 0.359 for BMI, 0.369 for sex and 0.638 for income. At 26 weeks the weighted omnibus test for age was p = 0.016 (set A; unweighted in the same members 0.038; full sample 0.052; Supplementary Table S34). The weighted overall 12-week estimate was –51.3 (–59.7 to –43.0) against –51.9 (–60.2 to –43.6) unweighted, but the unadjusted estimate moved from –43.8 (–52.7 to – 35.0) to –50.7 (–60.2 to –41.2) (Supplementary Table S34). The differences between subgroups were exploratory and, other than for ethnicity and age as above, not statistically significant: by sex the 12-week RTM-corrected change was –50.7 (–59.3 to –42.2) in women and –57.0 (–75.1 to –38.9) in men, and –39.0 (–43.6 to –34.5) and –51.9 (–62.2 to –41.5) in the pooled sample; by age the pooled change ranged from –22.3 (–36.3 to –8.4) at 18-34 years to –62.8 (–78.5 to –47.0) at 65 years or older; and at 26 weeks by BMI from –18.2 (–51.5 to 15.1; n = 20) below 25 kg/m^2^ to –68.1 (–80.4 to –55.8) at 35 or above. In every subgroup with more than 10 members, RTM-corrected change was an improvement at all landmarks.

**FIGURE 5.**
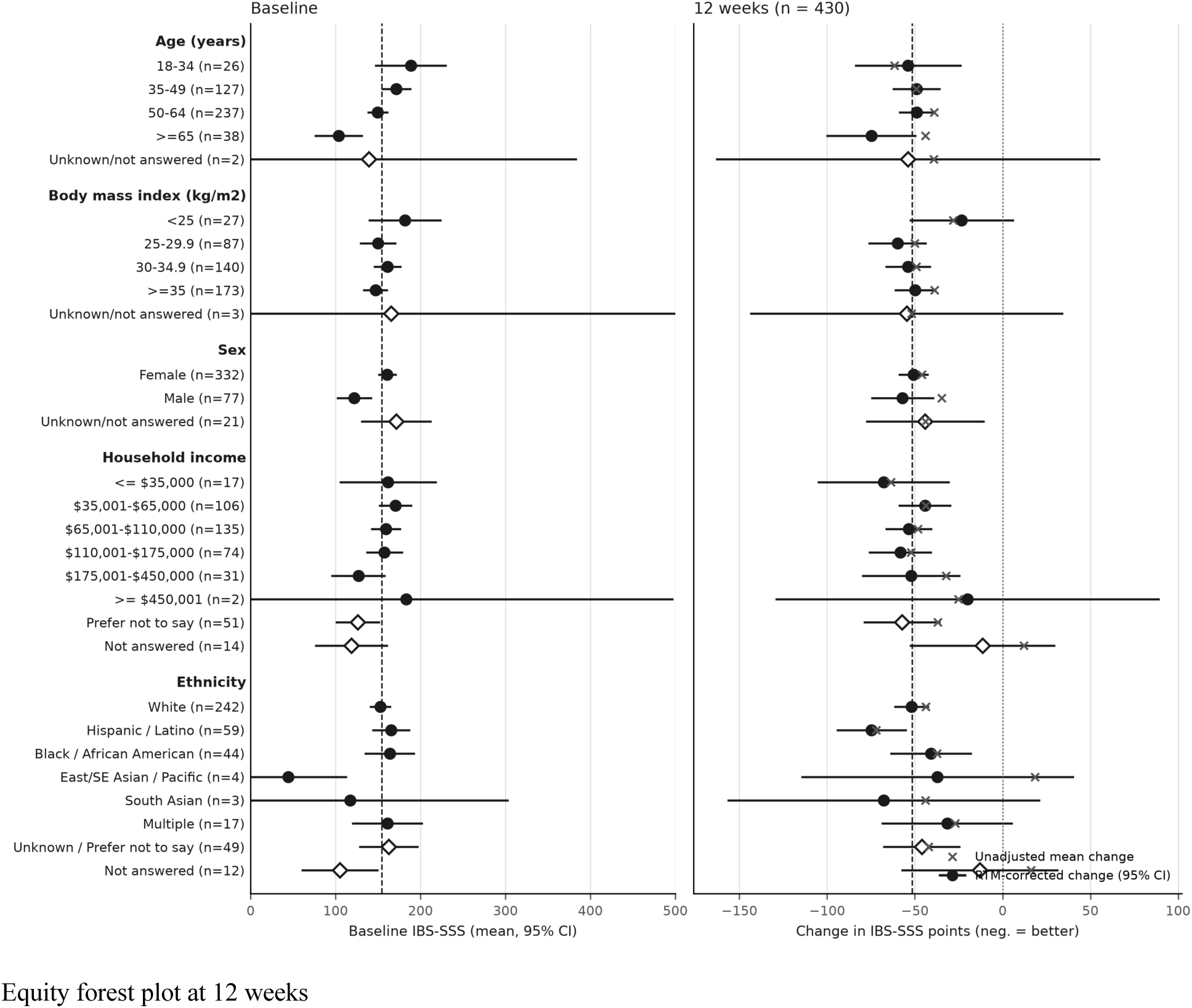
Baseline severity and 12-week change in IBS-SSS by subgroup. Left, mean baseline IBS-SSS with 95% CIs; right, RTM-corrected change (dots) and unadjusted change (crosses) with 95% CIs, for age, body-mass index, sex, household income and ethnicity (n = 430). The dashed line is the overall RTM-corrected change (–51.5; the equity model has no stratum terms). Omnibus tests of differences in RTM-corrected change: age p = 0.458, body-mass index p = 0.328, sex p = 0.749, income p = 0.486, ethnicity p = 0.170. Open diamonds mark members who did not answer. Exploratory; no multiplicity correction.

### Weight and ND-score

Weight change did not differ detectably between responders and non-responders: at 12 weeks –7.60 lb (95% CI –10.07 to –5.13; n = 140) versus –5.82 lb (–8.17 to –3.46; n = 121), difference –1.78 (–5.18 to 1.62; p = 0.303), with no difference at 4 weeks (p = 0.543) or 26 weeks (p = 0.337). Across all members weight fell by 3.53 lb at 4 weeks (–4.01 to –3.04; n = 1,564), 6.66 lb at 12 weeks (–7.40 to –5.93; n = 1,395) and 10.34 lb at 26 weeks (–11.40 to –9.28; n = 1,153), all p < 0.0001 (Supplementary Table S16). The ND-score rose by 0.47 (0.37 to 0.56; n = 1,313), 0.69 (0.57 to 0.81; n = 812) and 0.70 (0.53 to 0.88; n = 471) points at 4, 12 and 26 weeks (all p < 0.0001), with no difference by responder status (12 weeks: 0.91 versus 0.70; p = 0.445; Supplementary Table S17).

We carried out a within versus between members decomposition of the longitudinal IBS-SSS score changes. Within members, weight was not associated with IBS-SSS (0.236 points per lb, –0.295 to 0.768; p = 0.361), but a higher ND-score was associated with lower IBS-SSS (–6.05 points per ND point; –11.02 to –1.08; p = 0.018), as was a higher between-member score (–4.68; –8.26 to –1.09; p = 0.011), without evidence that the two differ (contrast p = 0.663) (Supplementary Table S18). When analysing the ND-score’s by penalisation domain, a higher within-member rate of inflammation penalties was associated with higher IBS-SSS (+2.48 points per 10 percentage points of meals; 0.66 to 4.31; p = 0.008), whereas fibre-diversity and insulin penalisation rates were not (p = 0.219 and 0.802; Supplementary Table S18). In a sensitivity analysis adding member-level covariates, the within-member coefficients did not change (ND-score –5.84 points per ND point, –10.81 to –0.87, p = 0.022, at base versus –5.83, –10.80 to –0.85, p = 0.022, with all covariates; n = 1,365 observations from 1,036 members; weight 0.245, –0.290 to 0.779, p = 0.348, versus 0.258, –0.278 to 0.794, p = 0.323; n = 1,823 from 1,308), whereas the between-member ND-score coefficient moved from –4.60 (–8.20 to –1.00; p = 0.012) to –1.02 (–4.71 to 2.66; p = 0.585) and the weight coefficient from –0.005 (–0.099 to 0.089; p = 0.916) to 0.059 (–0.126 to 0.245; p = 0.529) (Supplementary Table S19). The covariates improved model fit (likelihood-ratio p < 0.01 for each model) but did not narrow the between-member interval for the ND-score (width +0.1% to +2.5%) and widened it for weight (about 98%), and the Mundlak contrast was not significant for weight or ND-score in any model. Between-member associations were therefore sensitive to member-level covariates, whereas within-member associations were not.

### Sensitivity analyses

Pooled and 12-week estimates under each sensitivity analysis are summarised in Supplementary Table S20. The remission stratum served as a negative control, because members near the floor of the scale are expected to show an apparent rise from RTM alone. In the pooled longitudinal sample (n = 344) it rose by 24.08 points unadjusted (95% CI 17.32 to 30.84) and fell by 19.12 RTM-corrected (–38.71 to +0.47; p = 0.056). The reliability formula expects a shift of +38.72 at the retest reliability of 0.719, giving a corrected change of –14.64 (–21.25 to –8.02; Supplementary Table S21). The observed rise has the direction RTM predicts, but the correction cannot be verified independently, and each 0.1 of reliability moves the pooled expected shift by 13.78 points. Removing one covariate at a time from the RTM ANCOVA left the estimates stable: at 12 weeks (n = 407) the model-based estimate was –50.95 (–59.53 to –42.38), moving to –45.84 without the baseline term and by 1.2 points or less without age, sex or BMI; pooled (n = 1,592) it was –39.06 (–43.61 to –34.52), with shifts of at most 2.24 (Supplementary Table S22).

To evaluate attrition within the longitudinal sample, we weighted the members observed in each landmark window (337, 430 and 345 of 1,683) by the inverse probability of being observed (Supplementary Table S23). Weighting had little impact on the RTM-corrected estimates: change moved from –30.91 to –32.38 at 4 weeks, from –51.94 to –51.35 at 12 weeks and from –59.45 to –57.82 at 26 weeks (all moves under 2 points; weight set B –32.59, –51.67 and –57.82), with an effective sample of 93.8% to 98.1% of the complete cases. Non-response mattered more for the unadjusted estimate: at 12 weeks the unadjusted change of –43.83 became –50.70 with set A (–51.12 with B), whereas at 4 and 26 weeks it moved from – 39.15 to –31.77 and from –55.43 to –56.87, so the unadjusted figure depends on who responded and the RTM-corrected figure does not. Balancing early engagement (set C, adding the number of distinct app-activity days in the first 14 days after baseline) barely moved the estimates further: RTM-corrected change was –32.87, –51.23 and –56.12 at 4, 12 and 26 weeks, a difference from set A of –0.49, +0.12 and +1.70 points (unadjusted –0.64, –0.02 and +1.53), with a weight range of 0.54 to 2.06 and an effective sample of 92.8% to 95.9%; early engagement was imbalanced between observed and all members at 12 weeks (standardised difference 0.178) and was balanced by set C (0.019) (Supplementary Table S23). Weights built from measured characteristics cannot remove selection on unmeasured engagement or on symptom course after baseline. For all members with a reading, unadjusted –40.50 became –37.39 and RTM-corrected –41.04 became –39.48 under IPW-A (–43.82 to –35.13), with IPW-B giving –38.81 and – 39.12 (–43.51 to –34.73) (Supplementary Table S23). In the delta-based tipping-point sensitivity analysis for data missing not at random, the change of the members without a reading in a landmark window was set to the observed mean plus a delta. The RTM-corrected mean change over all 1,683 members reached zero at deltas of 39.36 (4 weeks), 69.13 (12 weeks) and 73.31 points (26 weeks); the unadjusted deltas were 49.08, 58.93 and 68.79. Only at 4 weeks was zero reached within the 50-point benchmark (Supplementary Table S24).

For medication, antidepressant users (n = 440 in the longitudinal sample, of whom 420 by intake list and 67 by claims) had higher baseline scores (188.0 versus 165.6; difference +22.4, 95% CI 11.1 to 33.7; p = 0.0001), but their RTM-corrected 12-week change did not differ from non-users (–48.54 versus –52.73; difference +3.76, –13.71 to +21.24; p = 0.672; Supplementary Table S25). Baseline GLP-1 receptor agonist users (n = 260) had similar baseline scores (+1.1; p = 0.873), and adding antidepressant and GLP-1 terms moved the 12-week estimate from –47.73 to –47.58 (–55.31 to –39.85). Because GLP-1 receptor agonists can cause gastrointestinal adverse events, we asked whether starting one after enrolment explained symptom increases. Among members with a complete pharmacy window who were not baseline users, 72 who started a GLP-1 receptor agonist after baseline and 193 who did not were compared. Starters improved more: RTM-corrected difference –50.53 points (–72.53 to –28.53; p < 0.0001; unadjusted –44.49, –70.34 to –18.64; p = 0.0009), with imprecise landmark differences. Worsening of at least 50 points occurred in 8 of 72 starters (11.1%) and 38 of 193 non-starters (19.7%; risk difference –0.086, –0.178 to +0.006; Fisher p = 0.144), and was no more frequent in the claims-linked sensitivity analysis (9/82 versus 59/352; p = 0.238) (Supplementary Table S26; Supplementary Figure S7). Starting a GLP-1 receptor agonist therefore did not explain symptom increases.

Baseline readings were taken a median of 0 days from the index date (interquartile range 0 to 0): of 1,683 longitudinal members, 1,334 (79.3%) had a baseline on or before the index date, 333 (19.8%) 1 to 7 days after and 16 (1.0%) 8 to 14 days after. At 12 weeks, the change in all strata was –43.88 points unadjusted (95% CI –52.44 to –35.32) and –47.73 RTM-corrected (–55.48 to –39.98) in the full sample (n = 430), – 47.11 (–57.11 to –37.12) and –49.30 (–57.96 to –40.64) with baselines on or before the index date (n = 340), and –44.25 (–52.86 to –35.64) and –47.81 (–55.55 to –40.06) with baselines at most 7 days after it (n = 427). The 95% confidence intervals overlapped throughout, for every stratum and landmark, and the estimates were close (Supplementary Table S27).

### Analysis by IBS-SSS item

At 12 weeks (n = 430) all five items fell (Table 3; the unadjusted item changes sum to the total change of –43.88). Unadjusted, the largest fall was in interference with daily life (–14.52 points; 95% CI –17.43 to – 11.62), followed by pain frequency (–9.88; –12.74 to –7.02), distension (–9.57; –12.30 to –6.83), bowel-habit dissatisfaction (–5.58; –9.51 to –1.65) and pain severity (–4.32; –6.16 to –2.48); the pain domain (severity plus frequency, 0-200) fell by –14.20 (–17.98 to –10.43). After RTM correction the falls were – 17.06 (–19.39 to –14.72) for interference, –12.40 (–14.65 to –10.16) for distension, –11.29 (–13.26 to –9.31) for pain frequency, –7.32 (–10.41 to –4.24) for bowel-habit dissatisfaction and –6.31 (–7.82 to –4.80) for pain severity, and –17.06 (–19.95 to –14.18) for the pain domain. The ordering of items was therefore similar but not identical before and after correction. Items differ in baseline mean (pain severity 11.70, bowel-habit dissatisfaction 45.07 at 12 weeks), so points are not directly comparable across items, and pain frequency is a day count multiplied by 10. Results at 4 and 26 weeks are in Supplementary Table S28 and Supplementary Figure S8.

**TABLE 3.** Change in IBS-SSS by item and domain at 12 weeks (points; negative = improvement), unadjusted and RTM-corrected. Each item is scored 0-100; the pain domain (severity plus frequency) is an exploratory grouping and the total is shown for reference. RTM-corrected estimates are centred at each outcome’s longitudinal-sample mean baseline (total 171.42). Exploratory; items are correlated and no multiplicity correction was applied.

| Outcome (scale) | n | Unadjusted change [95% CI] | RTM-corrected change [95% CI] |
| --- | --- | --- | --- |
| Pain severity (0-100) | 430 | -4.32 [-6.16, -2.48] | -6.31 [-7.82, -4.80] |
| Pain frequency (0-100) | 430 | -9.88 [-12.74, -7.02] | -11.29 [-13.26, -9.31] |
| Distension (bloating) (0-100) | 430 | -9.57 [-12.30, -6.83] | -12.40 [-14.65, -10.16] |
| Bowel-habit dissatisfaction (0-100) | 430 | -5.58 [-9.51, -1.65] | -7.32 [-10.41, -4.24] |
| Interference with daily life (0-100) | 430 | -14.52 [-17.43, -11.62] | -17.06 [-19.39, -14.72] |
| Pain domain (severity + frequency; exploratory) (0-200) | 430 | -14.20 [-17.98, -10.43] | -17.06 [-19.95, -14.18] |
| IBS-SSS total (0-500) | 430 | -43.88 [-52.44, -35.32] | -47.73 [-55.48, -39.98] |

In continuous-time models of the 1,683 longitudinal members (2,342 follow-up readings), each item improved over the first months and then plateaued (Supplementary Table S29; Supplementary Figure S9). By point estimates, pain severity and pain frequency had the largest share of their 26-week fitted change already present at 4 weeks (0.92 and 0.81, against 0.53 to 0.63 for the other items), and these two items and the pain domain (0.83) ranked first descriptively; this is exploratory, the intervals overlapped widely, and no pairwise difference in this share or in the day at which 75% of the 26-week change was reached had a 95% interval excluding zero (Supplementary Table S30). Pain severity has a low baseline mean, so a floor may contribute. Continuous-time models showed no distinguishable difference in trajectory between items (item-by-time likelihood-ratio test p = 0.23; cluster-robust p = 0.62). The same held within each baseline severity stratum (quarterly-bin likelihood-ratio tests p = 0.68 to 0.98; Supplementary Figure S10 and Supplementary Tables S31 and S32); in the remission, mild and moderate strata pain severity and pain frequency were the two items with the largest early share by point estimate, and the severe stratum could not be assessed on RTM-corrected curves because no individual item had a fitted 26-week change below zero there. The cluster-robust test within strata was unreliable where time cells were sparse (p = 0.004 in the mild stratum, Supplementary Table S32).

## Discussion

### Principal findings

In this single-arm cohort, IBS-SSS fell by 43.88 points at 12 weeks unadjusted and by 47.73 after RTM correction, close to the 50-point benchmark, and just over half of symptomatic members met it. Members with higher baseline severity improved more even after correction, and RTM-corrected improvement was seen in all subgroups with more than 10 members, with omnibus tests of subgroup differences mostly not significant despite baseline differences (the ethnicity test depended on how missing covariates were handled), while follow-up itself differed by subgroup. These are associations in a self-selected program without a comparator.

### Change, trajectories and regression to the mean

The unadjusted fall is within the range of real-world evidence for digital IBS programs [24,25] and above the roughly 45-point fall in dietary trials [16]. RTM correction reduced the change in the severe stratum and reversed the rise in the remission stratum, yet a severity gradient remained, so the remission stratum can serve as a negative control for RTM correction in real-world studies. The early fall and later plateau in our trajectories resemble the time-course reported for diet and digital programs, in which IBS-SSS fell by about 100 points within 6 weeks in a diet trial [20] and by 81 and 104 points at the half-way point and the end of a digital cognitive behavioural therapy program among those who reached those sessions [25], with benefit persisting at 6 and 12 months only in the arm that received personalised guidance [21]. Our falls are smaller (RTM-corrected –34.7 to –51.5 at 12 weeks by stratum), as expected for an unselected real-world cohort analysed with RTM correction. Monthly-bin and smooth trajectory models agree in shape; the RTM-corrected remission estimates extrapolate, and the smooth model should be read as description.

### Equity

Informatics and digital interventions can widen inequalities when they work better for advantaged groups [31,38], and virtual gastrointestinal programs have been asked to show performance in diverse and underserved populations [30]. Older members and men began with milder disease, and women and younger members began more often in severe strata (the stratum mix differed by age and sex but not by body-mass index), yet RTM-corrected change did not differ significantly by age, BMI, sex or income at 12 weeks. The ethnicity test was not significant in all 430 members with a reading (p = 0.170) but was in the 407 with complete covariates (p = 0.033 unweighted, 0.023 weighted), and the weighted age test was significant at 26 weeks (p = 0.016), so we do not read the non-significant tests as showing equal benefit; the analyses were not designed to show equivalence and are exploratory. Subgroup estimates changed little when weighted for non-response, and improvement in every subgroup with more than 10 members encourages adapting them for additional subgroups and communities, for example with culturally tailored education and partnership-based implementation [39,76]. Our 26-week follow-up and subgroup analyses address two evidence gaps identified for virtual gastrointestinal programs, the durability of benefit and performance across diverse populations [30], although follow-up remains short and the equity groups are limited to members who answered demographic questions. Differences by age and sex at other landmarks were modest and exploratory. Follow-up was heterogeneous by age (higher at older ages), sex (higher in women) and whether income and ethnicity were answered (very low when not answered), so the equity estimates describe the members who were followed up, and weights for measured characteristics cannot correct selection on unmeasured factors.

### Cohort, reliability and responders

Respondents were 14.15% of valid-baseline members, more engaged and more often answering demographic questions than non-respondents, with similar baseline scores, so our inferences are biased towards more engaged participants, which is expected in real-world evidence studies. Test-retest agreement of the IBS-SSS (ICC(2,1) 0.709, 95% CI 0.668 to 0.748, median interval 4 days) is moderate by conventional bands [77] and close to the 0.72 (95% CI 0.50 to 0.85) reported for a 7 to 9 day retest of the Chinese version [50]; next-day repetitions in the original validation and in a Greek version were described as highly reproducible [49,51], and a German validation reported it as reproducible [78], so a shorter interval and a clinic setting may explain higher agreement. The bias of –25.71 points between baseline and a reading within 14 days is itself a reason to keep early readings out of change estimates. The responder proportion at 12 weeks (52.3%; 95% CI 46.9 to 57.7) is similar to the 50% reported for a 4-week low-FODMAP diet in a randomised trial, whose traditional-advice arm reached 46% [79], while placebo response in IBS drug trials is 27% to 38% [80,81]; responder proportions depend on baseline severity [82]. The overall 12-week proportion averages over mixed severity and is diluted by the mild stratum, in which more members fall below 75 (remission) and a 50-point fall is less often possible; among members with a baseline of 175 or more (moderate and severe combined), 61.8% (95% CI 54.3 to 68.7) responded at 12 weeks and 73.5% (66.1 to 79.9) at 26 weeks, closer to the severity of most trial populations (Supplementary Table S35). Because responder proportions depend on baseline severity [82], comparison with trials should be made within severity.

### Robustness

Estimates were stable to dropping covariates (2 points or less except for the baseline term, which does the correcting) and to weighting for members without a reading in each landmark window (RTM-corrected estimates moved by less than 2 points). The missing members’ change would have had to be 39 to 73 points worse than observed before the mean change reached zero, beyond the 50-point benchmark at 12 and 26 weeks; weighting covers measured characteristics only, including early program engagement in the third weight set, which moved estimates by less than 2 points but cannot capture unmeasured engagement, and single-imputation tipping intervals are anti-conservative. Formal sensitivity analyses for inverse-probability weighting that bound the effect of unmeasured selection [83] are a direction for future work. Antidepressant use marked more severe baseline symptoms without changing improvement. Starters of GLP-1 receptor agonists improved more and did not worsen more often, so these drugs, which cause gastrointestinal adverse events [65,66], do not explain the symptom increases we saw; but this rests on small groups with pharmacy data, confounded by indication.

### Weight and diet quality

Weight loss did not differ between responders and non-responders and was not associated with IBS-SSS within or between members, while a higher ND-score was associated with lower IBS-SSS at both levels, and more inflammation-penalised meals accompanied higher IBS-SSS within members; these are exploratory associations. IBS is associated with low-grade mucosal inflammation, altered immune function and microbiome differences from controls, although whether these are cause or consequence is unknown [84–87]. Pro-inflammatory dietary patterns have been associated with a higher risk of incident IBS [88] and with IBS in case-control and cross-sectional studies, although one cross-sectional study found no association with severity [89,90]. Our within-member finding is consistent with this literature; if it reflects a pathway it would link food choices to the microbiome and inflammation proposed to contribute to IBS, but this study cannot establish direction. The program also personalises avoidance of pro-inflammatory and trigger foods using genetics, for example lactose: the lactase persistence variant determines lactase expression in adults [91], the non-persistence genotype has been associated with diarrhoea-predominant IBS and symptoms after dairy in one study [92], and self-reported dairy sensitivity in IBS agrees poorly with genotype [93]. Genotype-informed avoidance is therefore a plausible but untested contributor to symptom change here. The between-member associations were sensitive to member-level covariates and the within-member associations were not; most of the shift came from engagement days, which are counted over the 90 days after the index date and include food-post days, so they are not a clean baseline covariate (without them the between-member ND-score coefficient was – 3.23, –6.82 to 0.36; p = 0.078).

### Items, inflammation, pain and the gut microbiome

The within-member association between a higher ND-score, driven mainly by its inflammation-penalty domain, and lower IBS-SSS is compatible with a hypothesised pathway in which less exposure to pro-inflammatory foods accompanies less low-grade mucosal immune activation and microbiome-related signalling to visceral afferents. IBS is associated with more colonic mast cells and higher circulating cytokines and faecal calprotectin than in controls [94–96], although calprotectin does not distinguish individuals with IBS from controls [97]; mast-cell mediators and faecal microbiota from patients induced visceral hypersensitivity in preclinical models [98,99]; microbial metabolites such as short-chain fatty acids have been proposed to link microbiota, barrier function and immunity [100]; and pro-inflammatory or ultra-processed dietary patterns have been associated with incident IBS [88,101]. Our descriptive finding that the pain items had the largest early share of change, although the item trajectories could not be statistically distinguished, is consistent with this idea but does not test it, because we measured no mucosal, immune, microbiome or pain-pathway outcome; item-level improvements in pain and bloating have been reported with a low-FODMAP diet [102], and pain and bloating improved at similar rates by week 2 in one randomised trial [103], so earlier improvement in pain items here should not be read as a feature of diet therapy in general. In our earlier longitudinal study of this program, time in the program was associated with shifts in gut microbial pathways, including GABA, nitric oxide and succinate metabolism [42], and Mediterranean-diet change has likewise been associated with microbiome shifts towards short-chain fatty acid producers and taxa inversely related to inflammatory markers [104], but in a weight-loss programme such time effects are difficult be separated from weight loss in real world evidence studies.

Whether personalising food avoidance with genetic and microbiome information reduces inflammatory exposure and, through it, abdominal pain should be examined in trials that measure faecal calprotectin, cytokines and microbiome profiles together with item-level IBS-SSS and pain over time, with comparisons against non-personalised diet advice [20,21,105].

### Limitations

As with any research setting, this real-world evidence study has limitations. Due to its setting the program lacks a comparator arm, so placebo and natural-history effects cannot be separated; most valid-baseline members had no follow-up, and follow-up differed by age, sex and whether income and ethnicity were answered, so inverse-probability weights for measured characteristics move the unadjusted estimate (12-week unadjusted change –50.70 weighted against –43.83 unweighted) but cannot address unmeasured selection, and the missing data; reliability, centring values and slopes come from the same cohort; outcomes and covariates are self-reported; claims are available for a minority of the sample and not uniformly; the program bundles components, so none can be credited; and generalisability is limited to members of a commercial program with self-enrollment who met any one of its several eligibility criteria (Methods). Baseline readings taken up to 14 days after the index date could include early program effects; results were similar when restricted to baselines on or before the index date or within 7 days after it. IBS subtype (constipation-, diarrhoea- or mixed-predominant) was not avialable. The ND-score is a proprietary artificial-intelligence-based rating, and its inter-rater reliability and validation against dietitian assessment are not reported here, so its association with IBS-SSS may reflect properties of the scoring. The item-level analysis is exploratory: items are correlated, differ in floor and granularity (pain frequency is a day count multiplied by 10), and the study was not designed to rank them. Genetic and microbiome guided personalization effect modification is not evaluated.

## Conclusions

In a digital food-as-medicine program, IBS-SSS fell by 43.88 points at 12 weeks (95% CI –52.44 to – 35.32) and by about 48 points after RTM correction (–55.48 to –39.98), more in members with higher baseline severity, and with mostly non-significant differences between demographic groups, although follow-up differed by subgroup. A higher meal nutrient density, and fewer pro-inflammatory meals, accompanied lower symptom scores within members. The result held under sensitivity and robustness analyses, and starting a GLP-1 receptor agonist did not explain symptom increases.

## Abbreviations

ANCOVA, analysis of covariance; BMI, body-mass index; CI, confidence interval; EPC, Established Pharmacologic Class; FODMAP, fermentable oligo-, di-, mono-saccharides and polyols; GLP-1, glucagon-like peptide-1; IBS, irritable bowel syndrome; IBS-SSS, IBS Symptom Severity Scale; ICC, intraclass correlation coefficient; IPW, inverse-probability weighting; ND, nutrient density; NDC, national drug code; RTM, regression to the mean.

## Declarations

### Ethics approval and consent to participate

The Institutional Review Board of E&I Review Services reviewed and approved the study protocol (protocol code #18053, approved 05/22/2018). Before being included in the study, all participants provided electronic informed consent as part of their written consent process.

## Consent for publication

Not applicable. No individual-level data, images or identifiable information are reported.

## Availability of data and materials

The data analysed in this study are member-level records from a commercial digital health program and cannot be shared publicly due to consent restrictions.

## Authors’ contributions

IP: Conceptualization, Data curation, Formal analysis, Investigation, Methodology, Resources, Software, Validation, Visualization, Writing – original draft, Writing – review & editing.

SVK: Writing – review & editing.

SKS: Data Collection, Methodology, Writing – review & editing.

Ronesh Sinha: Writing review & editing.

DEA: Conceptualization, Writing review & editing.

RS: Conceptualization, Funding acquisition, Project administration, Supervision, Writing – review & editing.

## Acknowledgements

We are grateful to subjects participating in Digbi Health research studies without whom it would not have been possible to perform this research.

## Supplementary material

### Supplementary methods

#### Nutrient-density score

Members photograph meals in the app. Each meal is scored by a coach or by an artificial-intelligence meal-rating pipeline against three categories of infractions (insulin, inflammation and fibre diversity). A meal starts from a maximum score of 14 and loses one or two points for each matched infraction, so that a higher ND-score is better, and the points lost to each category are stored with the meal. For every member-day we derived the ND-score, the share of the day’s scored meals with at least one infraction in each category (penalisation rate, percent of meals) and the mean points lost per scored meal in each category (penalty points). Readings on the same day were averaged; each IBS-SSS reading was matched to the nearest ND-score reading within 14 days, and changes were computed between the baseline and landmark readings.

#### Medication processing

Intake-form medication lists were matched to drug classes by the keywords in Table SM2 applied to the free text and structured entries. Pharmacy claims were received monthly in several layouts; each layout was parsed with a custom script, members were linked by name and date of birth, and records were de-identified in-house under the HIPAA safe-harbor method before analysis (the approach used for medical claims in an earlier Digbi study [44]). For each drug claim we kept the service date, the product name and the national drug code (NDC) of the package. The NDC was linked to the openFDA drug-label reference [52,53], downloaded on 10 July 2026, to obtain generic and brand names, active ingredients and the FDA Established Pharmacologic Class (EPC) strings. A claim or list entry counted toward a class if any of three routes matched: (1) a class keyword (Table SM2; case-insensitive substring) in the claim product name; (2) a class keyword in the openFDA generic or brand names or active ingredients matched by NDC; (3) the NDC carried the class EPC string (Table SM1; exact match, except the GLP-1 class, matched by substring). No code or class was assigned by hand. Class groups were antidepressants (selective serotonin reuptake inhibitors, serotonin-norepinephrine reuptake inhibitors, tricyclics and others), GLP-1 receptor agonists and proton-pump inhibitors; other classes in Table SM2 were screened but not analysed here. The claims window before baseline was 180 days before baseline to the day before; the window after baseline ran from baseline to 30 days after the last reading. A list entry dated more than 14 days after baseline counted as a later entry. A start was a first fill on or after baseline with none before, assessed only for members whose pharmacy feed covered the period from 180 days before baseline through the last reading. Members without an observed source were counted as not exposed.

**TABLE SM1.**
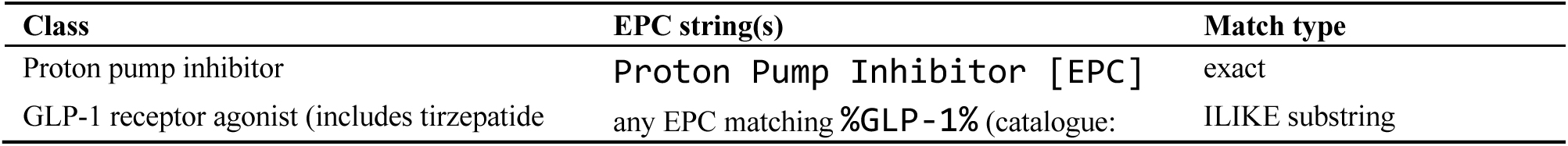
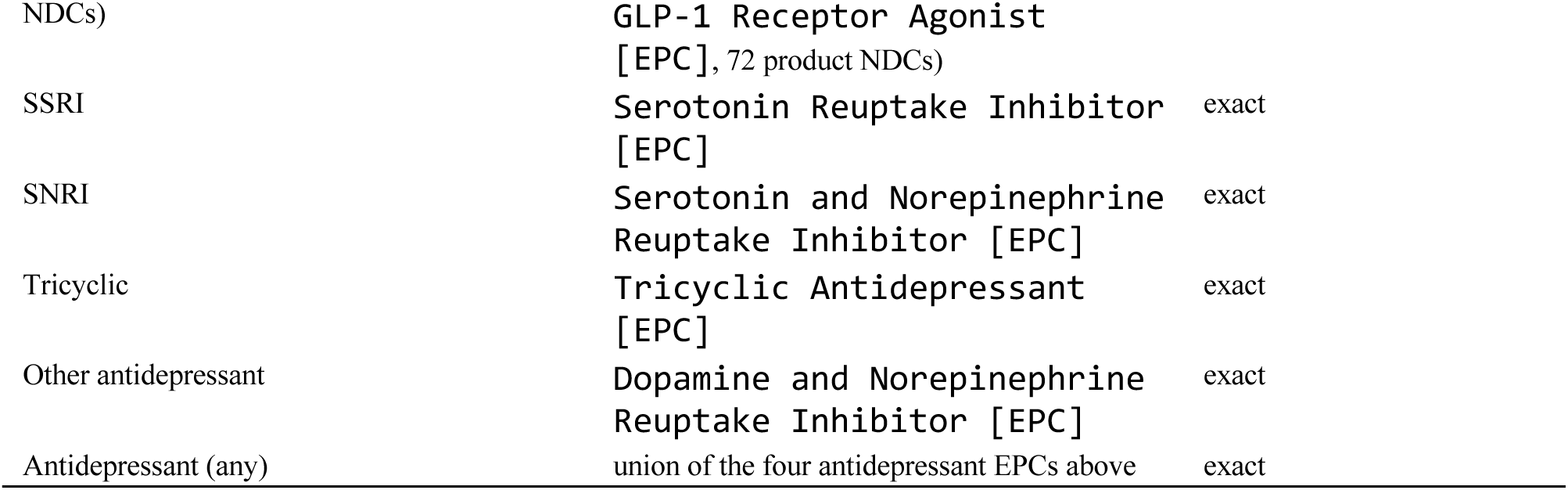
FDA Established Pharmacologic Class strings used to assign drug classes to national drug codes.

**TABLE SM2.**
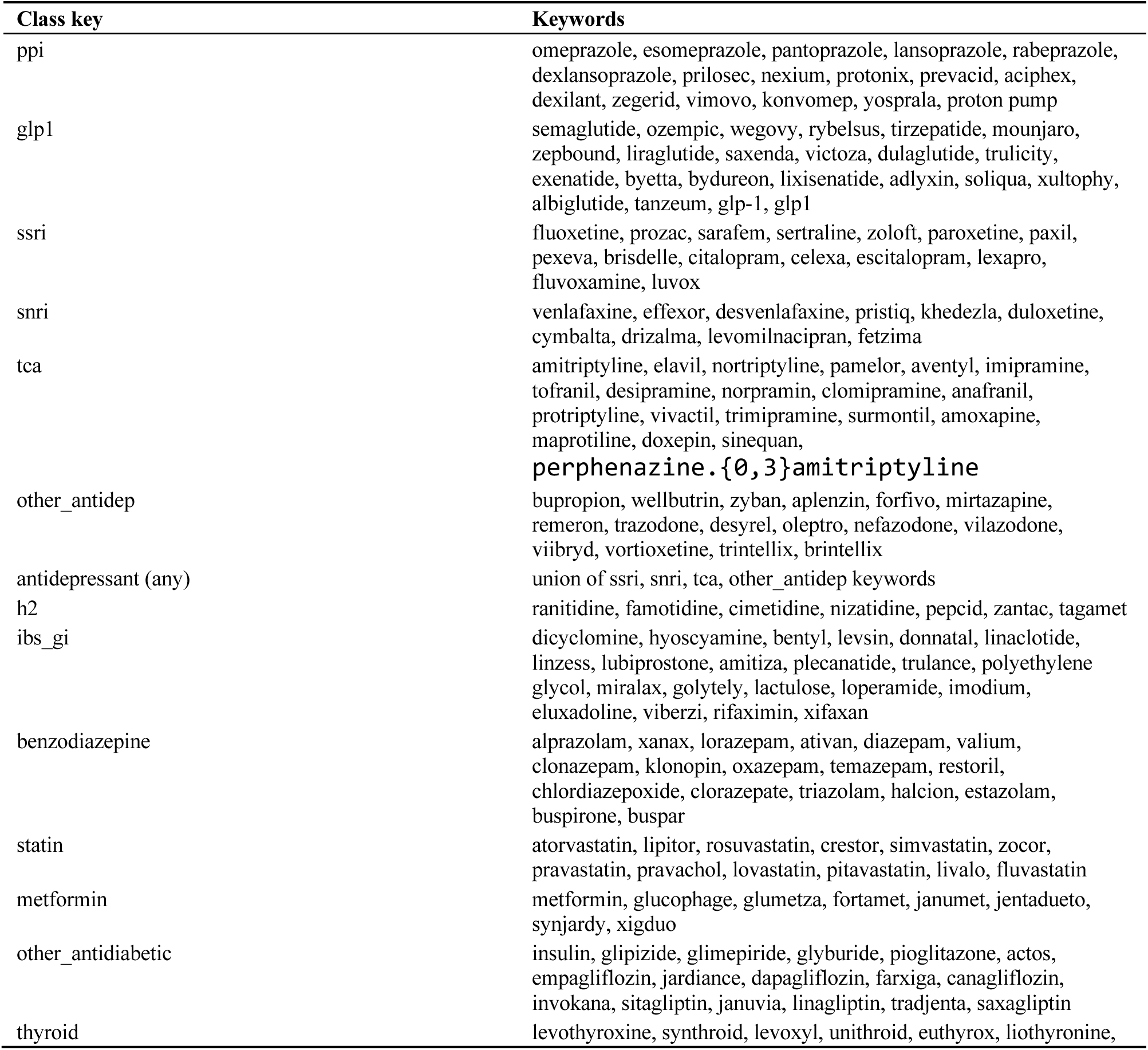
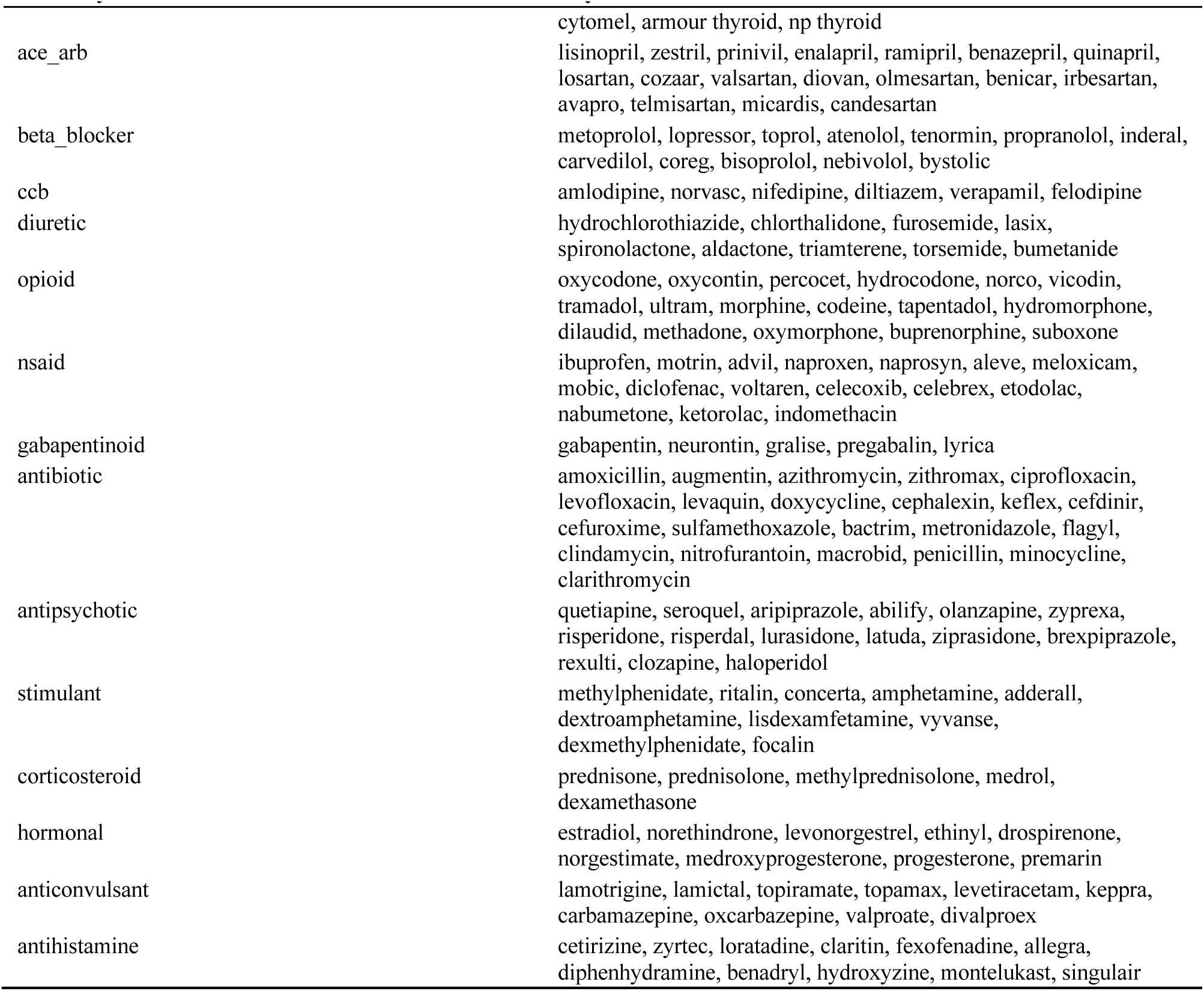
Class keywords applied to claim product names, openFDA names and intake-form medication lists (case-insensitive substrings; the tricyclic entry includes one regular expression).

#### Statistical models

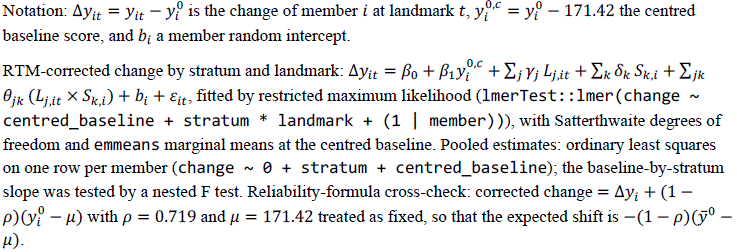

Baseline-timing sensitivity analysis: same model as primary (the RTM-corrected model above), fitted separately on the longitudinal members with a baseline offset from the index date of 0 days or less and of 7 days or less, with landmark windows defined from each member’s own baseline reading and the centring value fixed at the full-sample mean (171.42); the recentred sensitivity analysis replaced it by the mean baseline of the restricted sample.

Inverse-probability weighting at each landmark: among the 1,683 longitudinal members with a complete-case baseline (n = 1,592; weight set A uses baseline score, age, BMI and sex, set B adds log(1 + pre-baseline engagement days), set C adds log(1 + distinct app-activity days in days 0 to 13 from the baseline reading day), which precede every landmark window but are measured after baseline), Pr(observed in window) = logit^-1^(*α*_0_ + *α*_1_score + *α*_2_age + *α*_3_BMI + *α*_4_ male); weights *w_i_* = Pr(*R* = 1)/*p*^_*i*_ were stabilised and truncated at the 1st and 99th percentiles, and the weighted least squares regression followup ∼ I(baseline – 171.42) used HC1 robust variance. The unadjusted IPW estimate is the weighted mean change. Tipping-point analysis at each landmark: members without a reading in the window were assigned the observed mean change plus a delta, the mean over all 1,683 members was 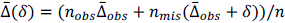, and the delta at which it reaches zero is reported. The pooled IPW-A and IPW-B models (responding at all; longitudinal membership) were fitted among 14,797 in-window members with a complete-case baseline in the same way.

Within/between decomposition: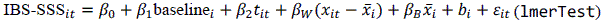, with CR2 cluster-robust variance (clubSandwich) and the Mundlak contrast *β*_2_ − *β*_1_. Three covariate-extended models were fitted on the occasions of the base model with a recorded age, plausible baseline BMI (18.5 to 80 kg/m2) and engagement count: (a) adding age, sex and baseline BMI; (b) adding ethnicity, household-income stratum and engagement days (distinct days with any app activity in the 90 days from the index date); and (c) adding baseline IBS-SSS stratum and baseline medication from the intake-form list only (list present, antidepressant, GLP-1 receptor agonist). Not-recorded and not-answered levels were kept as their own categories. Models were compared with the base model by likelihood-ratio tests on maximum-likelihood refits, with AIC and Nakagawa marginal and conditional R2.

#### Item analysis

The five IBS-SSS items were taken from the same complete submissions as the total score (each item 0-100; pain frequency is the number of days with pain in the last 10 multiplied by 10), so the five unadjusted item changes add to the total change exactly (checked at every landmark). The pain domain (severity plus frequency, 0-200) is an exploratory grouping, not a validated sub-scale. Landmark change per item used the same readings, strata and models as the total score, with the item’s own baseline as covariate and centring at the item’s longitudinal-sample baseline mean. Continuous-time models were fitted to the 1,683 longitudinal members: monthly-bin models (glmmTMB, unadjusted; lmerTest, RTM-corrected change ∼ centred_baseline + bin + (1 | member)) and smooth models (gamm4, thin-plate basis, k = 8, change ∼ centred_baseline + s(time_days) with a member random intercept, and the same without the baseline term for unadjusted change). Within each baseline severity stratum the smooth models used a stratum-specific curve with a common baseline slope. The early share was the fitted change at day 28 divided by the fitted change at day 182; days to 25%, 50%, 75% and 90% of the 26-week change were the first day in days 14 to 182 at which the fitted change reached that fraction (a value of 14 means reached by the first observable day). These fractions are exploratory descriptors, and their intervals come from a member-level bootstrap (400 replicates) that refits all items on the same resampled members. Whether items differed over time was tested in a stacked five-item model of change as a percentage of each item’s baseline mean (chg_pct ∼ item + item:centred_baseline + bin + item:bin + (1 | member) + (1 | reading)), by a likelihood-ratio test and a cluster-robust Wald test, overall (monthly bins) and within each stratum (quarterly bins, because monthly cells are sparse in small strata). The test addresses the shape and relative size of profiles together, not timing alone.

## Supplementary Tables

**TABLE S1.**
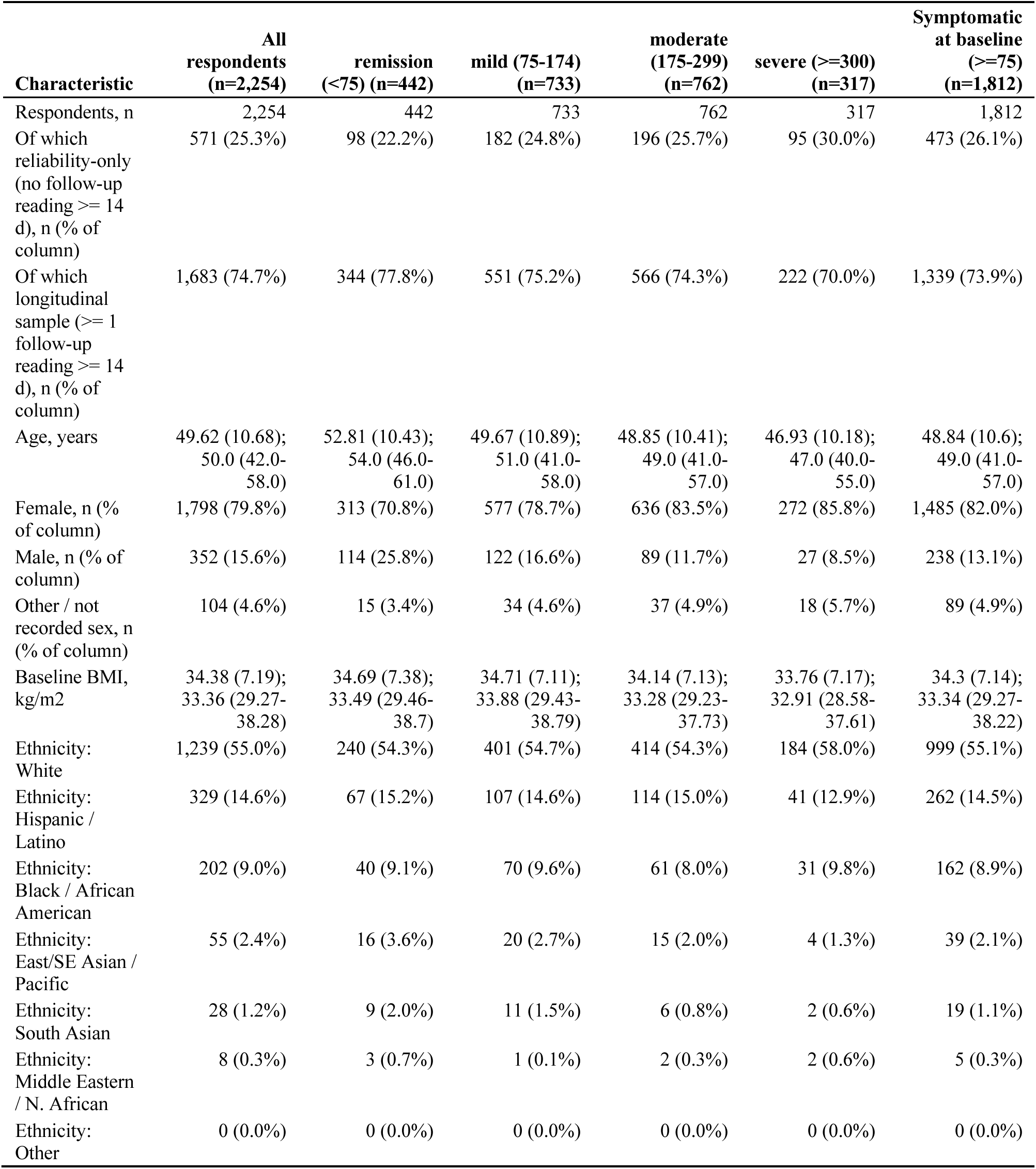
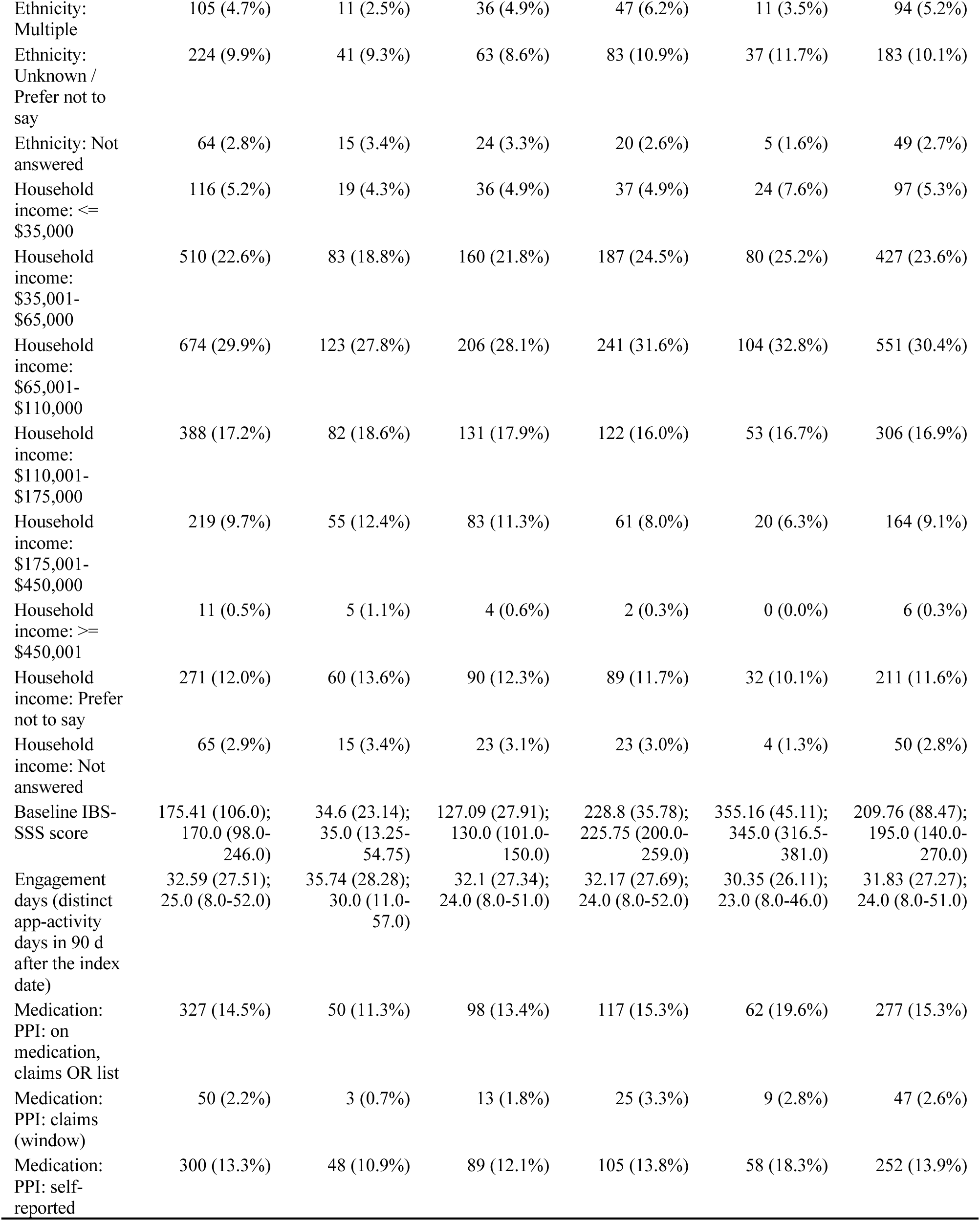
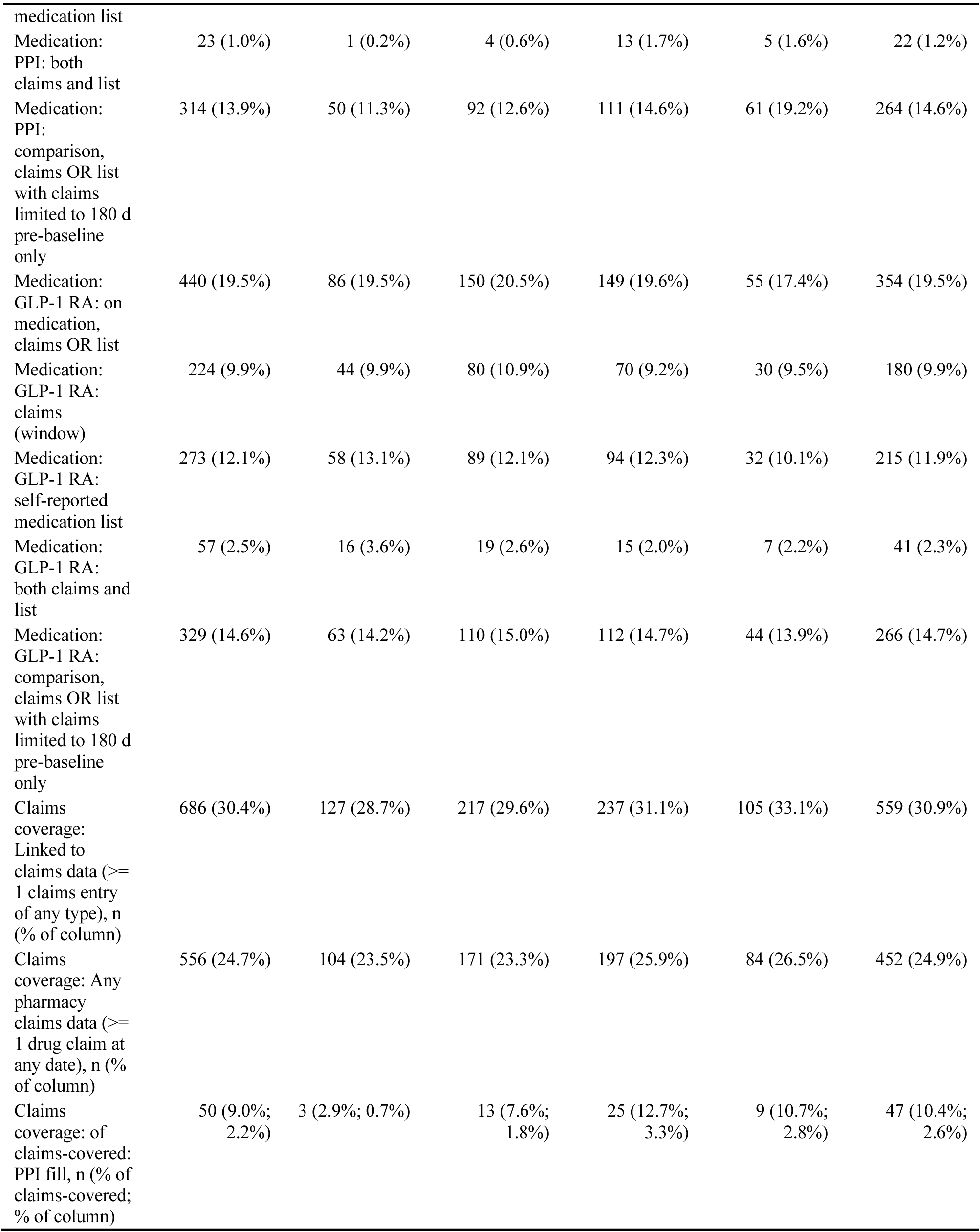
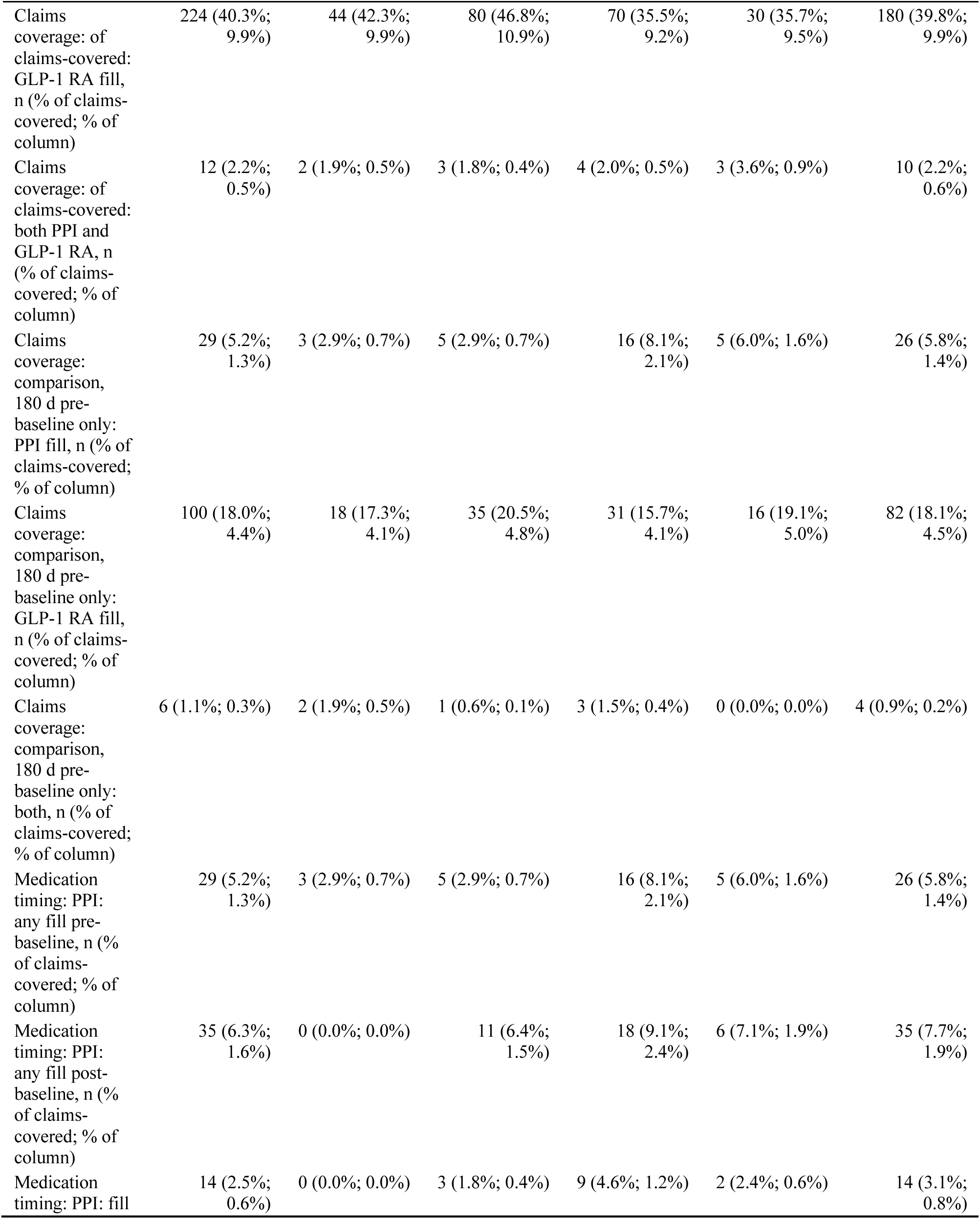
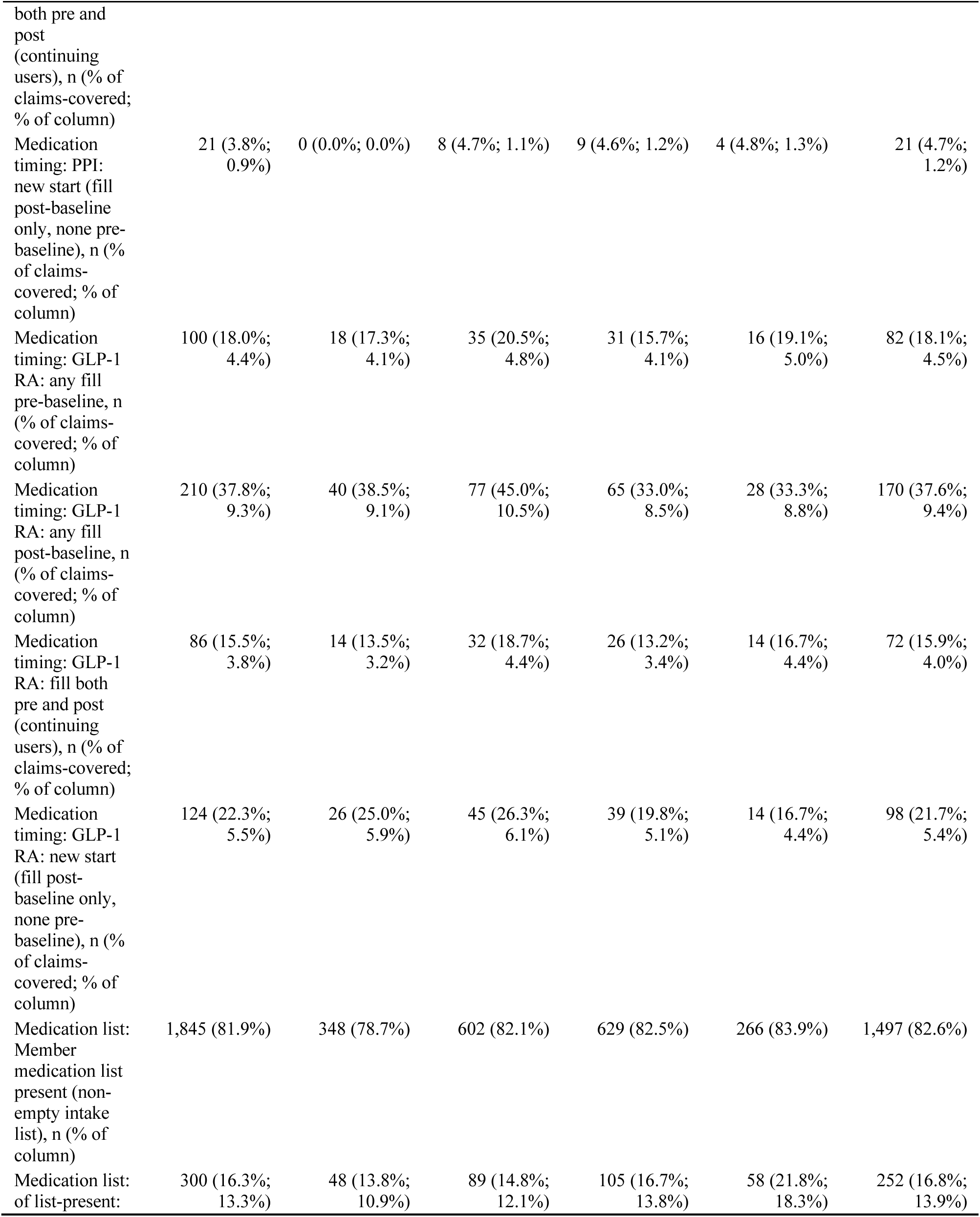
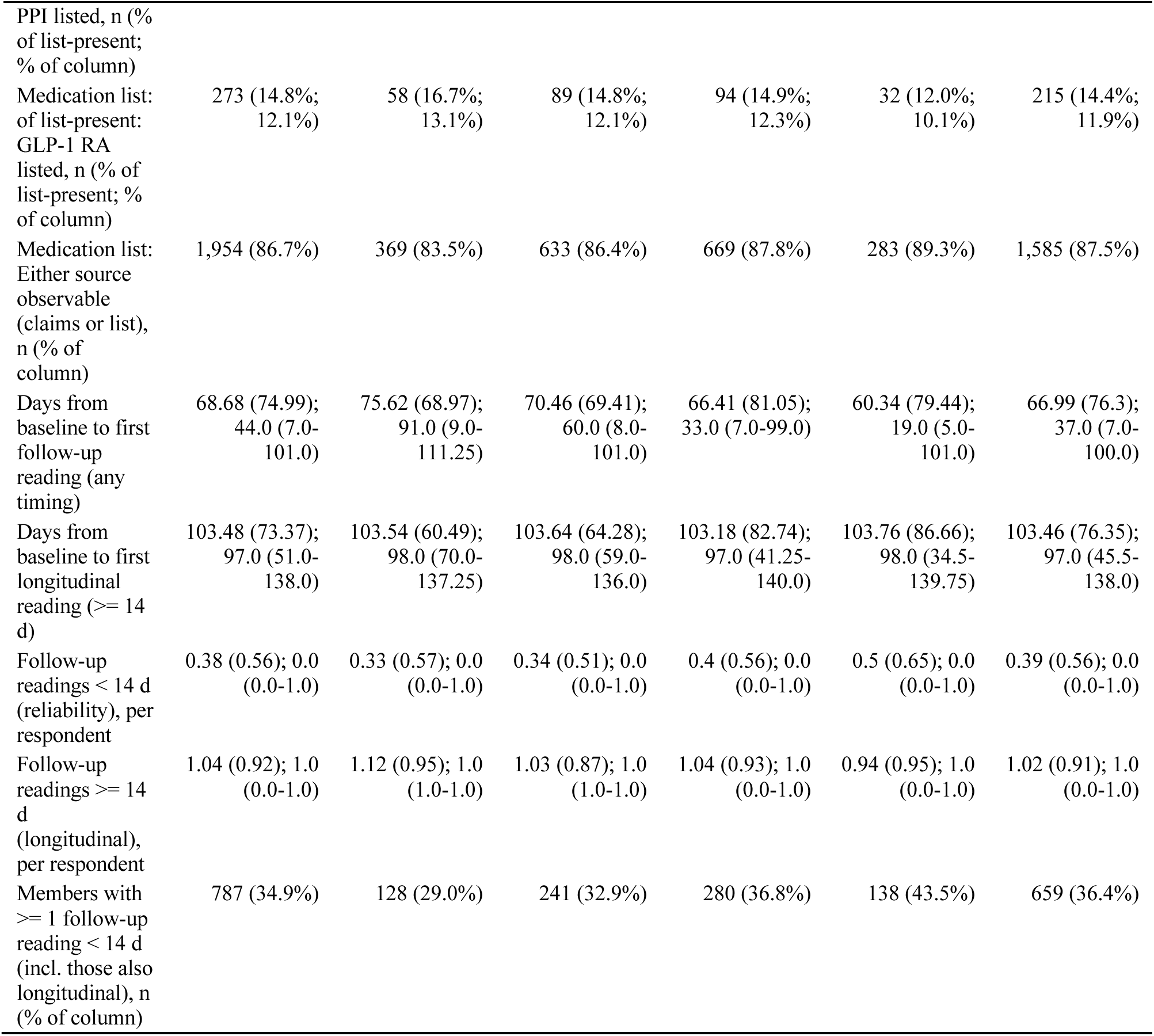
Baseline characteristics, medication and claims detail, by baseline IBS-SSS stratum. Cells are n (%) or mean (SD); median (interquartile range).

**TABLE S2.** Follow-up rate by baseline IBS-SSS stratum.

| Baseline stratum | In-window baseline n | Respondents n | Follow-up rate % (95% CI) | Longitudinal sample n (>= 1 reading >= 14 d) | % of in-window | Reliability-only n |
| --- | --- | --- | --- | --- | --- | --- |
| remission (<75) | 3,081 | 442 | 14.35 (13.15-15.63) | 344 | 11.17 | 98 |
| mild (75-174) | 5,333 | 733 | 13.74 (12.85-14.69) | 551 | 10.33 | 182 |
| moderate (175-299) | 5,501 | 762 | 13.85 (12.96-14.79) | 566 | 10.29 | 196 |
| severe (>=300) | 2,018 | 317 | 15.71 (14.19-17.36) | 222 | 11.0 | 95 |
| All in-window baseline members | 15,933 | 2,254 | 14.15 (13.61-14.7) | 1,683 | 10.56 | 571 |

**TABLE S3.**
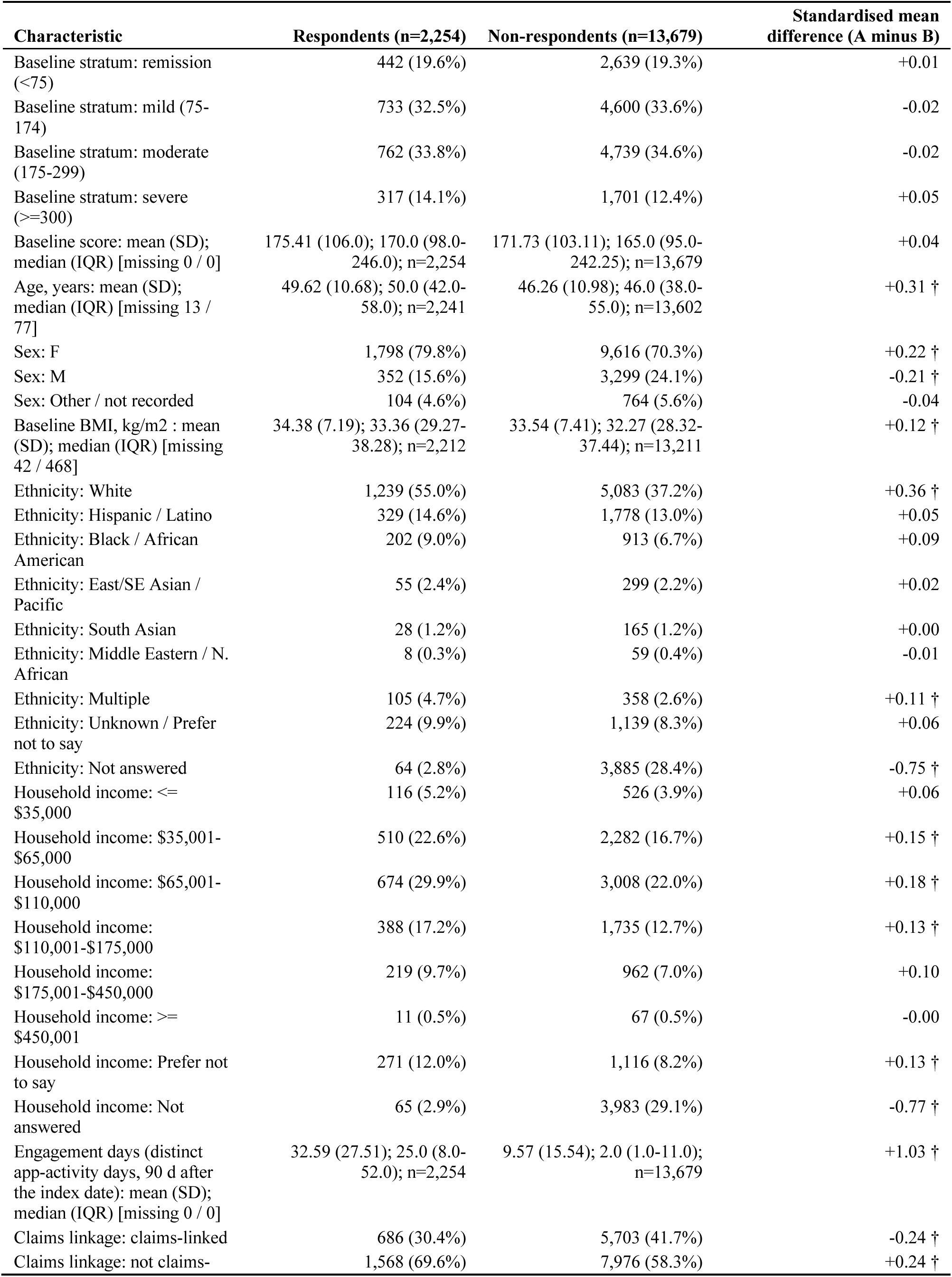
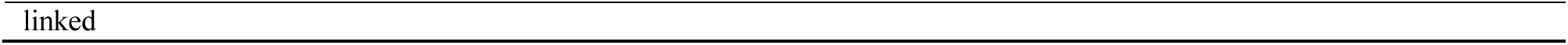
Respondents versus non-respondents (members with a valid baseline and no follow-up reading). Standardised mean differences (respondents minus non-respondents); a dagger marks absolute values above 0.1.

**TABLE S4.** Longitudinal sample versus reliability-only respondents. Standardised mean differences (longitudinal minus reliability-only); a dagger marks absolute values above 0.1.

| Characteristic | Longitudinal sample (n=1,683) | Reliability-only (n=571) | Standardised mean difference (A minus B) |
| --- | --- | --- | --- |
| Baseline stratum: remission (<75) | 344 (20.4%) | 98 (17.2%) | +0.08 |
| Baseline stratum: mild (75-174) | 551 (32.7%) | 182 (31.9%) | +0.02 |
| Baseline stratum: moderate (175-299) | 566 (33.6%) | 196 (34.3%) | -0.01 |
| Baseline stratum: severe (>=300) | 222 (13.2%) | 95 (16.6%) | -0.10 |
| Baseline score: mean (SD); median (IQR) [missing 0 / 0] | 171.42 (104.72); 165.0 (93.0-240.0); n=1,683 | 187.19 (108.91); 175.0 (104.5-270.0); n=571 | -0.15 † |
| Age, years: mean (SD); median (IQR) [missing 8 / 5] | 50.11 (10.59); 51.0 (42.0-58.0); n=1,675 | 48.18 (10.83); 48.0 (41.0-56.0); n=566 | +0.18 † |
| Sex: F | 1,345 (79.9%) | 453 (79.3%) | +0.01 |
| Sex: M | 262 (15.6%) | 90 (15.8%) | -0.01 |
| Sex: Other / not recorded | 76 (4.5%) | 28 (4.9%) | -0.02 |
| Baseline BMI, kg/m2 : mean (SD); median (IQR) [missing 27 / 15] | 34.38 (7.06); 33.32 (29.32-38.42); n=1,656 | 34.37 (7.56); 33.46 (29.09-37.76); n=556 | +0.00 |
| Ethnicity: White | 915 (54.4%) | 324 (56.7%) | -0.05 |
| Ethnicity: Hispanic / Latino | 244 (14.5%) | 85 (14.9%) | -0.01 |
| Ethnicity: Black / African American | 154 (9.2%) | 48 (8.4%) | +0.03 |
| Ethnicity: East/SE Asian / Pacific | 40 (2.4%) | 15 (2.6%) | -0.02 |
| Ethnicity: South Asian | 24 (1.4%) | 4 (0.7%) | +0.07 |
| Ethnicity: Middle Eastern / N. African | 6 (0.4%) | 2 (0.3%) | +0.00 |
| Ethnicity: Multiple | 76 (4.5%) | 29 (5.1%) | -0.03 |
| Ethnicity: Unknown / Prefer not to say | 167 (9.9%) | 57 (10.0%) | -0.00 |
| Ethnicity: Not answered | 57 (3.4%) | 7 (1.2%) | +0.14 † |
| Household income: <= \$35,000 | 80 (4.8%) | 36 (6.3%) | -0.07 |
| Household income: \$35,001-\$65,000 | 387 (23.0%) | 123 (21.5%) | +0.04 |
| Household income: \$65,001-\$110,000 | 488 (29.0%) | 186 (32.6%) | -0.08 |
| Household income: \$110,001-\$175,000 | 287 (17.1%) | 101 (17.7%) | -0.02 |
| Household income: \$175,001-\$450,000 | 165 (9.8%) | 54 (9.5%) | +0.01 |
| Household income: >= \$450,001 | 10 (0.6%) | 1 (0.2%) | +0.07 |
| Household income: Prefer not to say | 208 (12.4%) | 63 (11.0%) | +0.04 |
| Household income: Not answered | 58 (3.5%) | 7 (1.2%) | +0.15 † |

| Characteristic | Longitudinal sample<br>(n=1,683) | Reliability-only (n=571) | Standardised mean<br>difference (A minus B) |
| --- | --- | --- | --- |
| Engagement days (distinct app-activity days, 90 d after the index date): mean (SD); median (IQR) [missing 0 / 0] | 34.98 (28.11); 28.0 (10.0-56.0); n=1,683 | 25.56 (24.35); 18.0 (5.0-39.0); n=571 | +0.36 † |
| Claims linkage: claims-linked | 537 (31.9%) | 149 (26.1%) | +0.13 † |
| Claims linkage: not claims-linked | 1,146 (68.1%) | 422 (73.9%) | -0.13 † |

**TABLE S5.**
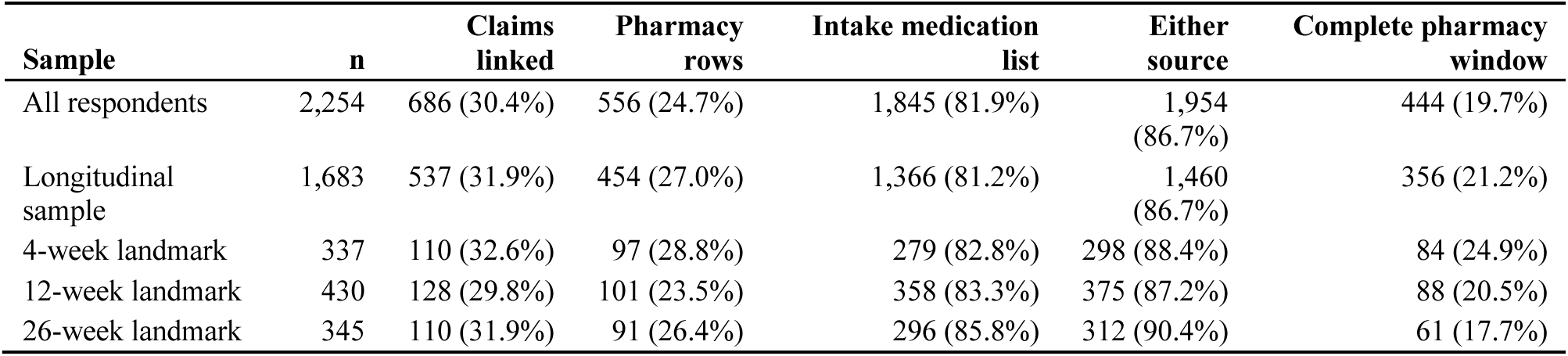
Medication data coverage by sample. Complete pharmacy window means the pharmacy feed starts at least 180 days before baseline and runs to at least the last reading.

**TABLE S6.** Responder proportions, n/N = % [Wilson 95% CI], by reading set; no p value is given because none is defined for a single proportion.

| Criterion | Longitudinal sample | Any timing | First reading at 30 days or later | 4 weeks | 12 weeks | 26 weeks |
| --- | --- | --- | --- | --- | --- | --- |
| ≥ 50-point decrease (benchmark threshold) | 720/1,339 = 53.8% [51.1, 56.4] | 886/1,812 = 48.9% [46.6, 51.2] | 644/1,165 = 55.3% [52.4, 58.1] | 139/279 = 49.8% [44.0, 55.7] | 169/323 = 52.3% [46.9, 57.7] | 172/273 = 63.0% [57.1, 68.5] |
| Follow-up score < 75 (remission) | 371/1,339 = 27.7% [25.4, 30.2] | 416/1,812 = 23.0% [21.1, 25.0] | 333/1,165 = 28.6% [26.1, 31.2] | 61/279 = 21.9% [17.4, 27.1] | 101/323 = 31.3% [26.5, 36.5] | 86/273 = 31.5% [26.3, 37.2] |

**TABLE S7.** Severity-stratum shifts (improved, same, worsened) by reading set.

| Reading set | Improved | Same | Worsened |
| --- | --- | --- | --- |
| Longitudinal sample: first reading at ≥ 14 days | 712 | 700 | 271 |
| All respondents: first follow-up reading, any timing | 873 | 1,017 | 364 |
| Pooled: first reading at ≥ 30 days (sensitivity definition) | 639 | 605 | 238 |
| Landmark 4 weeks (days 14-35) | 135 | 152 | 50 |
| Landmark 12 weeks (days 70-98) | 174 | 192 | 64 |
| Landmark 26 weeks (days 154-210) | 166 | 141 | 38 |

**TABLE S8.**
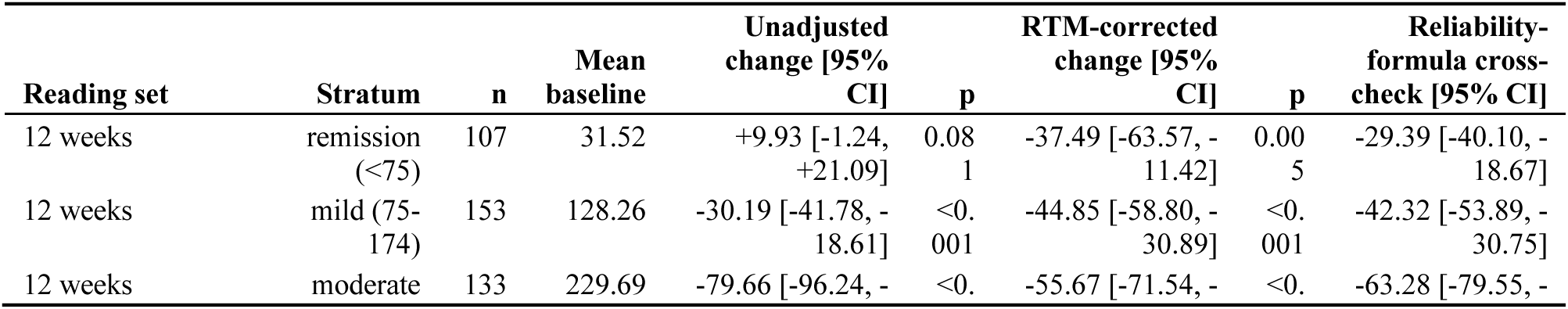
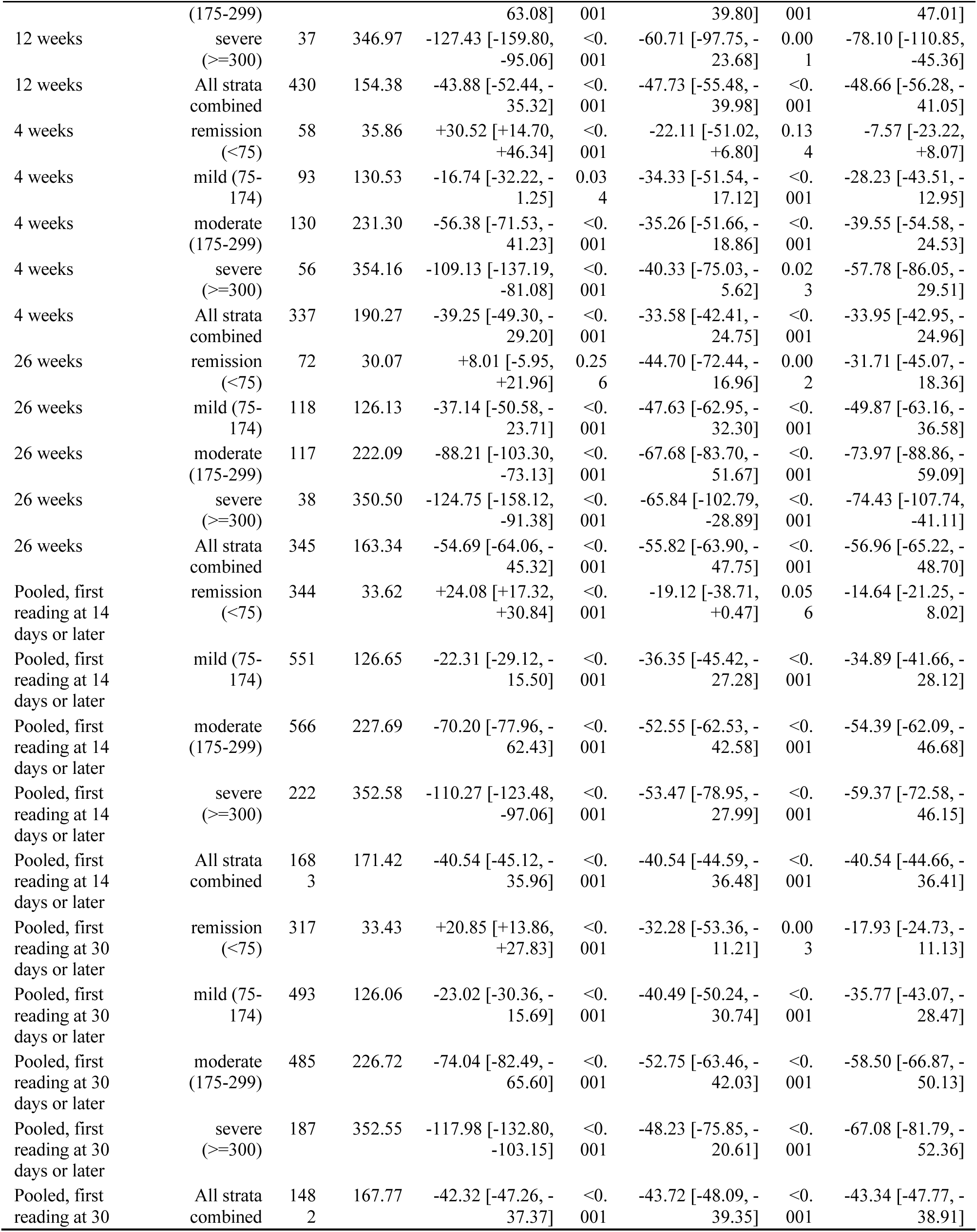
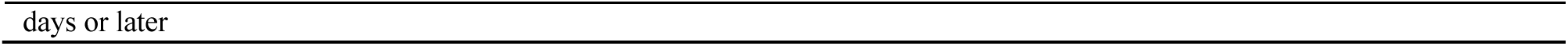
Change in IBS-SSS by baseline stratum at each landmark and pooled. Change is follow-up minus baseline (negative = improvement); RTM-corrected change is centred at 171.42. The last column is the reliability-formula cross-check, with the reliability given in the Results.

**TABLE S9.** Change estimated at the overall mean baseline (171.42) and at each stratum’s own mean baseline, with 95% CIs.

| Stratum | Landmark (weeks) | n | RTM-corrected at overall mean baseline [95% CI] | RTM-corrected at stratum own mean baseline [95% CI] |
| --- | --- | --- | --- | --- |
| remission (<75) | 4 | 58 | -22.11 [-51.02, +6.80] | +28.21 [+8.44, +47.99] |
| remission (<75) | 12 | 107 | -37.49 [-63.57, -11.42] | +12.83 [-1.80, +27.46] |
| remission (<75) | 26 | 72 | -44.70 [-72.44, -16.96] | +5.62 [-11.63, +22.88] |
| mild (75-174) | 4 | 93 | -34.33 [-51.54, -17.12] | -17.98 [-33.99, -1.97] |
| mild (75-174) | 12 | 153 | -44.85 [-58.80, -30.89] | -28.50 [-40.73, -16.26] |
| mild (75-174) | 26 | 118 | -47.63 [-62.95, -32.30] | -31.28 [-44.94, -17.62] |
| moderate (175-299) | 4 | 130 | -35.26 [-51.66, -18.86] | -55.81 [-69.32, -42.30] |
| moderate (175-299) | 12 | 133 | -55.67 [-71.54, -39.80] | -76.22 [-89.28, -63.16] |
| moderate (175-299) | 26 | 117 | -67.68 [-83.70, -51.67] | -88.23 [-102.02, -74.44] |
| severe (≥300) | 4 | 56 | -40.33 [-75.03, -5.62] | -106.49 [-126.90, -86.07] |
| severe (≥300) | 12 | 37 | -60.71 [-97.75, -23.68] | -126.87 [-151.98, -101.76] |
| severe (≥300) | 26 | 38 | -65.84 [-102.79, -28.89] | -132.00 [-156.56, -107.44] |
| remission (<75) | pooled longitudinal | 344 | -19.12 [-38.71, +0.47] | +24.08 [+15.12, +33.05] |
| mild (75-174) | pooled longitudinal | 551 | -36.35 [-45.42, -27.28] | -22.31 [-29.40, -15.23] |
| moderate (175-299) | pooled longitudinal | 566 | -52.55 [-62.53, -42.58] | -70.20 [-77.19, -63.21] |
| severe (≥300) | pooled longitudinal | 222 | -53.47 [-78.95, -27.99] | -110.27 [-121.44, -99.11] |

**TABLE S10.** Twelve-week change in IBS-SSS by baseline stratum from the monthly-bin and smooth trajectory models, unadjusted and RTM-corrected (all respondents, n = 2,254). Values are mean change [95% CI] and p.

| Stratum | Monthly-bin, unadjusted | Monthly-bin, RTM-corrected | Smooth, unadjusted | Smooth, RTM-corrected |
| --- | --- | --- | --- | --- |
| Remission | +2.4 [-19.9, +24.7], 0.83 | -34.7 [-68.1, -1.3], 0.042 | +10.1 [+3.9, +16.2], 0.001 | -24.6 [-43.5, -5.7], 0.011 |
| Mild | -33.4 [-57.4, -9.4], 0.006 | -39.6 [-63.5, -15.6], 0.001 | -21.0 [-27.3, -14.6], <0.001 | -37.1 [-46.1, -28.0], <0.001 |
| Moderate | -59.8 [-85.8, -33.7], <0.001 | -51.5 [-74.7, -28.4], <0.001 | -72.6 [-80.3, -64.9], <0.001 | -50.9 [-60.7, -41.2], <0.001 |
| Severe | -111.4 [-154.8, -68.0], <0.001 | -42.3 [-81.4, -3.2], 0.034 | -124.1 [-137.8, -110.4], <0.001 | -47.0 [-72.8, -21.2], <0.001 |

**TABLE S11.**
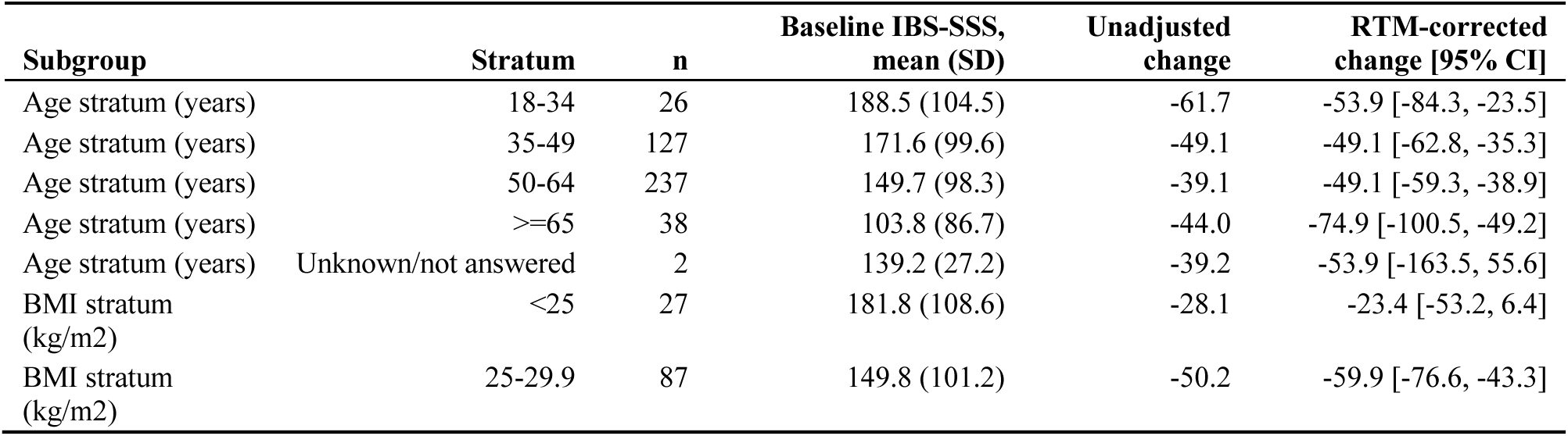
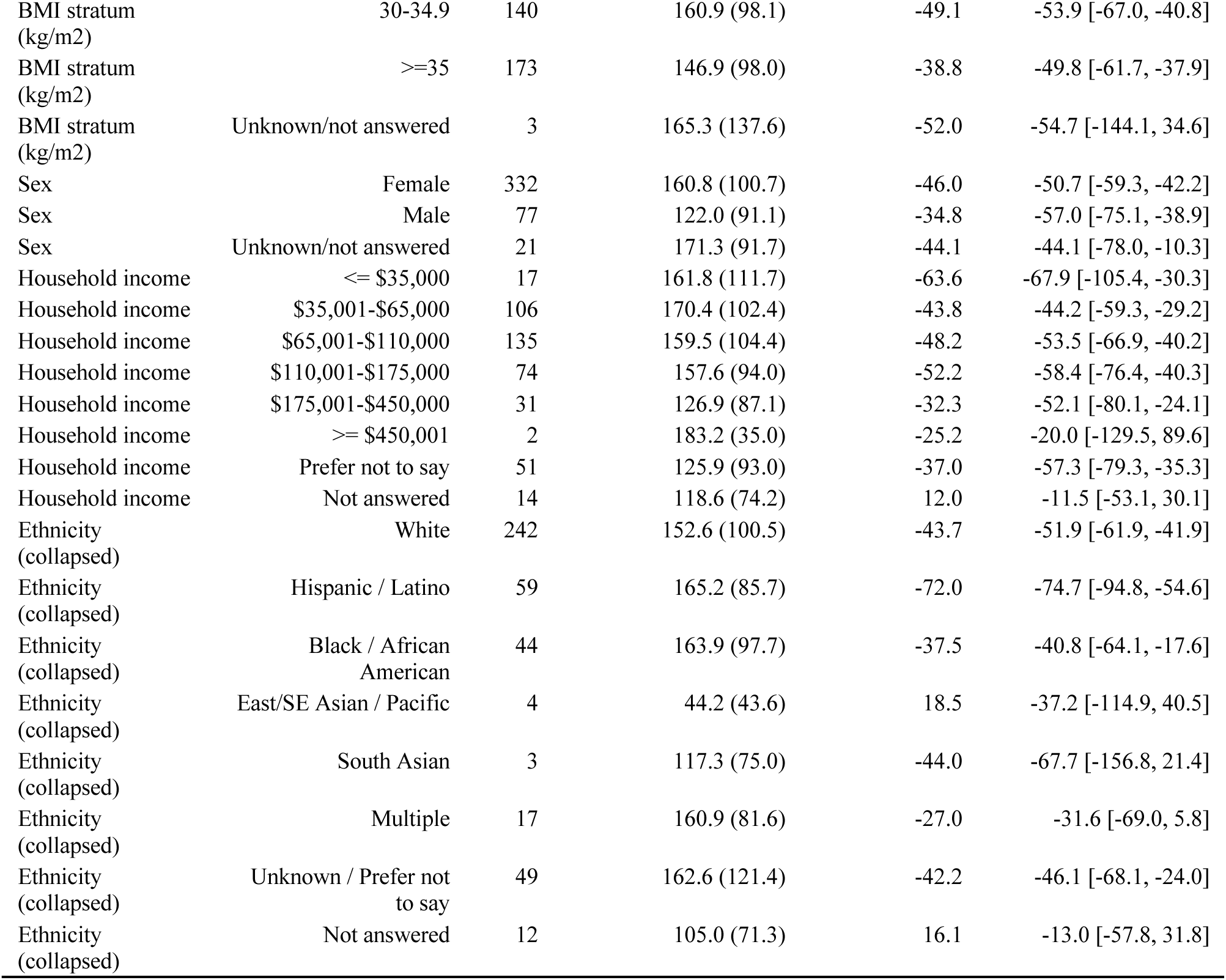
Change in IBS-SSS at 12 weeks by subgroup (n = 430). RTM-corrected change is centred at 171.42; the unadjusted change is the raw mean.

| Subgroup | Stratum | n | Baseline IBS-SSS, mean (SD) | Unadjusted change | RTM-corrected change [95% CI] |
| --- | --- | --- | --- | --- | --- |
| Age stratum (years) | 18-34 | 26 | 188.5 (104.5) | -61.7 | -53.9 [-84.3, -23.5] |
| Age stratum (years) | 35-49 | 127 | 171.6 (99.6) | -49.1 | -49.1 [-62.8, -35.3] |
| Age stratum (years) | 50-64 | 237 | 149.7 (98.3) | -39.1 | -49.1 [-59.3, -38.9] |
| Age stratum (years) | ≥65 | 38 | 103.8 (86.7) | -44.0 | -74.9 [-100.5, -49.2] |
| Age stratum (years) | Unknown/not answered | 2 | 139.2 (27.2) | -39.2 | -53.9 [-163.5, 55.6] |
| BMI stratum (kg/m2) | <25 | 27 | 181.8 (108.6) | -28.1 | -23.4 [-53.2, 6.4] |
| BMI stratum (kg/m2) | 25-29.9 | 87 | 149.8 (101.2) | -50.2 | -59.9 [-76.6, -43.3] |

| Subgroup | Stratum | n | Baseline IBS-SSS,<br>mean (SD) | Unadjusted<br>change | RTM-corrected<br>change [95% CI] |
| --- | --- | --- | --- | --- | --- |
| BMI stratum<br>(kg/m2) | 30-34.9 | 140 | 160.9 (98.1) | -49.1 | -53.9 [-67.0, -40.8] |
| BMI stratum<br>(kg/m2) | >=35 | 173 | 146.9 (98.0) | -38.8 | -49.8 [-61.7, -37.9] |
| BMI stratum<br>(kg/m2) | Unknown/not answered | 3 | 165.3 (137.6) | -52.0 | -54.7 [-144.1, 34.6] |
| Sex | Female | 332 | 160.8 (100.7) | -46.0 | -50.7 [-59.3, -42.2] |
| Sex | Male | 77 | 122.0 (91.1) | -34.8 | -57.0 [-75.1, -38.9] |
| Sex | Unknown/not answered | 21 | 171.3 (91.7) | -44.1 | -44.1 [-78.0, -10.3] |
| Household income | <= \$35,000 | 17 | 161.8 (111.7) | -63.6 | -67.9 [-105.4, -30.3] |
| Household income | \$35,001-\$65,000 | 106 | 170.4 (102.4) | -43.8 | -44.2 [-59.3, -29.2] |
| Household income | \$65,001-\$110,000 | 135 | 159.5 (104.4) | -48.2 | -53.5 [-66.9, -40.2] |
| Household income | \$110,001-\$175,000 | 74 | 157.6 (94.0) | -52.2 | -58.4 [-76.4, -40.3] |
| Household income | \$175,001-\$450,000 | 31 | 126.9 (87.1) | -32.3 | -52.1 [-80.1, -24.1] |
| Household income | >= \$450,001 | 2 | 183.2 (35.0) | -25.2 | -20.0 [-129.5, 89.6] |
| Household income | Prefer not to say | 51 | 125.9 (93.0) | -37.0 | -57.3 [-79.3, -35.3] |
| Household income | Not answered | 14 | 118.6 (74.2) | 12.0 | -11.5 [-53.1, 30.1] |
| Ethnicity<br>(collapsed) | White | 242 | 152.6 (100.5) | -43.7 | -51.9 [-61.9, -41.9] |
| Ethnicity<br>(collapsed) | Hispanic / Latino | 59 | 165.2 (85.7) | -72.0 | -74.7 [-94.8, -54.6] |
| Ethnicity<br>(collapsed) | Black / African<br>American | 44 | 163.9 (97.7) | -37.5 | -40.8 [-64.1, -17.6] |
| Ethnicity<br>(collapsed) | East/SE Asian / Pacific | 4 | 44.2 (43.6) | 18.5 | -37.2 [-114.9, 40.5] |
| Ethnicity<br>(collapsed) | South Asian | 3 | 117.3 (75.0) | -44.0 | -67.7 [-156.8, 21.4] |
| Ethnicity<br>(collapsed) | Multiple | 17 | 160.9 (81.6) | -27.0 | -31.6 [-69.0, 5.8] |
| Ethnicity<br>(collapsed) | Unknown / Prefer not<br>to say | 49 | 162.6 (121.4) | -42.2 | -46.1 [-68.1, -24.0] |
| Ethnicity<br>(collapsed) | Not answered | 12 | 105.0 (71.3) | 16.1 | -13.0 [-57.8, 31.8] |

**TABLE S12.** Change in IBS-SSS at 4 weeks by subgroup. RTM-corrected change is centred at 171.42.

| Subgroup | Stratum | n | Unadjusted change | RTM-corrected change<br>[95% CI] |
| --- | --- | --- | --- | --- |
| Age stratum (years) | 18-34 | 36 | -34.4 | -25.2 [-52.5, 2.1] |
| Age stratum (years) | 35-49 | 132 | -37.1 | -23.7 [-38.2, -9.3] |
| Age stratum (years) | 50-64 | 151 | -41.2 | -37.9 [-51.2, -24.6] |
| Age stratum (years) | >=65 | 15 | -50.1 | -45.5 [-87.7, -3.3] |
| Age stratum (years) | Unknown/not answered | 3 | -40.0 | -37.8 [-132.1, 56.5] |
| BMI stratum (kg/m2) | <25 | 18 | -39.7 | -7.5 [-46.5, 31.5] |
| BMI stratum (kg/m2) | 25-29.9 | 87 | -43.1 | -35.4 [-53.0, -17.8] |
| BMI stratum (kg/m2) | 30-34.9 | 94 | -41.3 | -32.4 [-49.3, -15.5] |
| BMI stratum (kg/m2) | >=35 | 136 | -33.8 | -30.1 [-44.1, -16.1] |
| BMI stratum (kg/m2) | Unknown/not answered | 2 | -141.8 | -115.4 [-230.8, 0.1] |
| Sex | Female | 285 | -38.3 | -27.8 [-37.7, -18.0] |
| Sex | Male | 35 | -47.7 | -55.7 [-83.2, -28.1] |
| Sex | Unknown/not answered | 17 | -37.3 | -39.0 [-78.4, 0.5] |
| Household income | <= \$35,000 | 16 | -49.0 | -48.0 [-89.0, -7.0] |
| Household income | \$35,001-\$65,000 | 86 | -36.2 | -23.4 [-41.2, -5.5] |
| Household income | \$65,001-\$110,000 | 95 | -42.1 | -32.1 [-49.1, -15.2] |
| Household income | \$110,001-\$175,000 | 53 | -50.1 | -40.2 [-62.9, -17.6] |
| Household income | \$175,001-\$450,000 | 38 | -23.0 | -23.2 [-49.8, 3.4] |
| Household income | >= \$450,001 | 4 | 30.0 | -0.4 [-82.5, 81.8] |
| Household income | Prefer not to say | 42 | -45.5 | -40.6 [-65.9, -15.3] |
| Household income | Not answered | 3 | -3.3 | -22.8 [-117.5, 71.9] |
| Ethnicity (collapsed) | White | 202 | -31.5 | -23.9 [-35.4, -12.3] |
| Ethnicity (collapsed) | Hispanic / Latino | 54 | -61.8 | -56.7 [-78.9, -34.5] |
| Ethnicity (collapsed) | Black / African American | 21 | -39.8 | -32.7 [-68.2, 2.8] |
| Ethnicity (collapsed) | East/SE Asian / Pacific | 5 | -23.2 | -40.2 [-113.0, 32.7] |
| Ethnicity (collapsed) | South Asian | 4 | -49.2 | -30.0 [-111.4, 51.5] |
| Ethnicity (collapsed) | Middle Eastern / N. African | 1 | 22.0 | 37.1 [-125.6, 199.8] |
| Ethnicity (collapsed) | Multiple | 12 | -58.2 | -43.0 [-90.1, 4.0] |
| Ethnicity (collapsed) | Unknown / Prefer not to say | 34 | -53.1 | -38.9 [-67.0, -10.9] |
| Ethnicity (collapsed) | Not answered | 4 | 29.2 | 20.4 [-60.9, 101.8] |

**TABLE S13.**
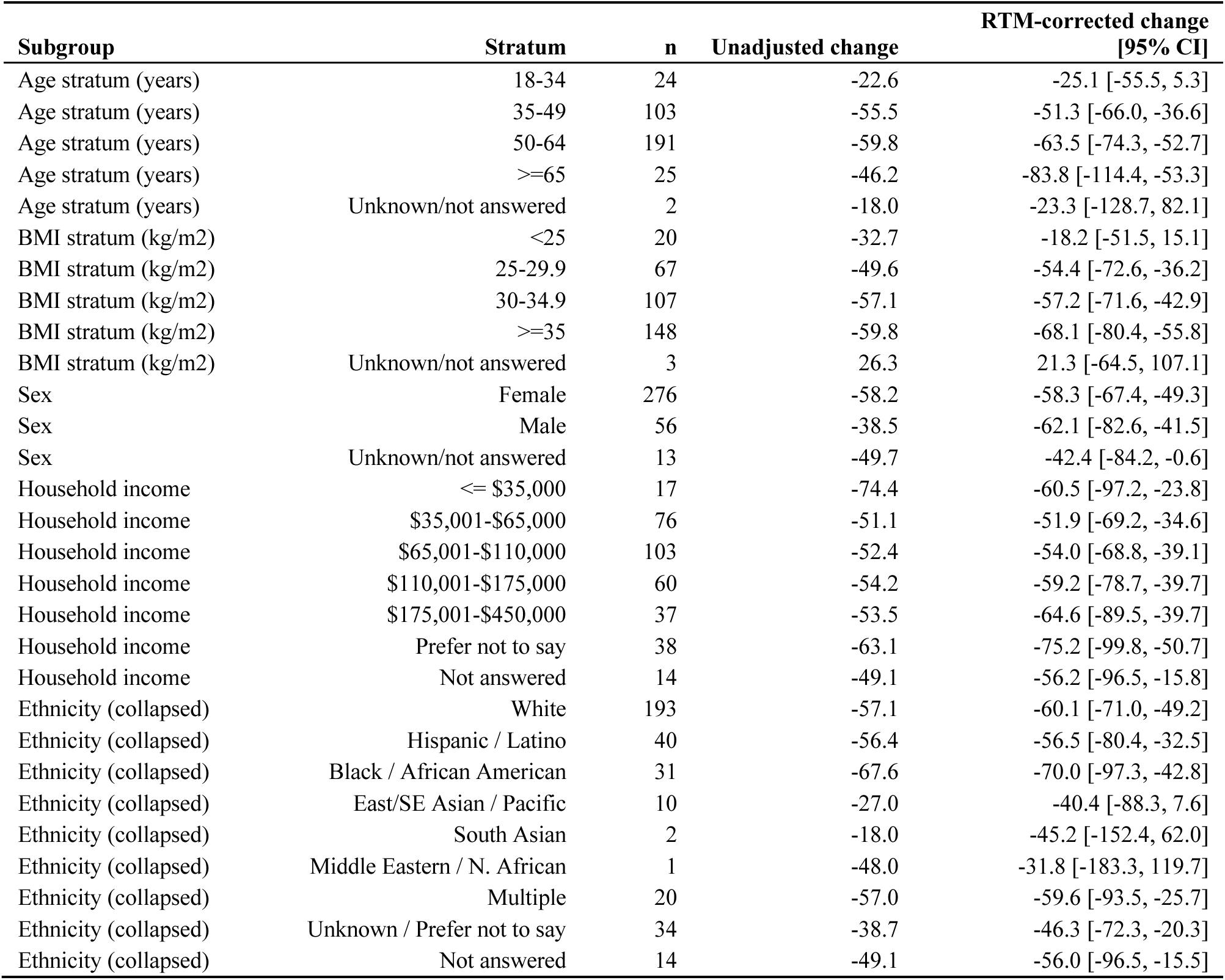
Change in IBS-SSS at 26 weeks by subgroup. RTM-corrected change is centred at 171.42

| Subgroup | Stratum | n | Unadjusted change | RTM-corrected change<br>[95% CI] |
| --- | --- | --- | --- | --- |
| Age stratum (years) | 18-34 | 24 | -22.6 | -25.1 [-55.5, 5.3] |
| Age stratum (years) | 35-49 | 103 | -55.5 | -51.3 [-66.0, -36.6] |
| Age stratum (years) | 50-64 | 191 | -59.8 | -63.5 [-74.3, -52.7] |
| Age stratum (years) | >=65 | 25 | -46.2 | -83.8 [-114.4, -53.3] |
| Age stratum (years) | Unknown/not answered | 2 | -18.0 | -23.3 [-128.7, 82.1] |
| BMI stratum (kg/m2) | <25 | 20 | -32.7 | -18.2 [-51.5, 15.1] |
| BMI stratum (kg/m2) | 25-29.9 | 67 | -49.6 | -54.4 [-72.6, -36.2] |
| BMI stratum (kg/m2) | 30-34.9 | 107 | -57.1 | -57.2 [-71.6, -42.9] |
| BMI stratum (kg/m2) | >=35 | 148 | -59.8 | -68.1 [-80.4, -55.8] |
| BMI stratum (kg/m2) | Unknown/not answered | 3 | 26.3 | 21.3 [-64.5, 107.1] |
| Sex | Female | 276 | -58.2 | -58.3 [-67.4, -49.3] |
| Sex | Male | 56 | -38.5 | -62.1 [-82.6, -41.5] |
| Sex | Unknown/not answered | 13 | -49.7 | -42.4 [-84.2, -0.6] |
| Household income | <= \$35,000 | 17 | -74.4 | -60.5 [-97.2, -23.8] |
| Household income | \$35,001-\$65,000 | 76 | -51.1 | -51.9 [-69.2, -34.6] |
| Household income | \$65,001-\$110,000 | 103 | -52.4 | -54.0 [-68.8, -39.1] |
| Household income | \$110,001-\$175,000 | 60 | -54.2 | -59.2 [-78.7, -39.7] |
| Household income | \$175,001-\$450,000 | 37 | -53.5 | -64.6 [-89.5, -39.7] |
| Household income | Prefer not to say | 38 | -63.1 | -75.2 [-99.8, -50.7] |
| Household income | Not answered | 14 | -49.1 | -56.2 [-96.5, -15.8] |
| Ethnicity (collapsed) | White | 193 | -57.1 | -60.1 [-71.0, -49.2] |
| Ethnicity (collapsed) | Hispanic / Latino | 40 | -56.4 | -56.5 [-80.4, -32.5] |
| Ethnicity (collapsed) | Black / African American | 31 | -67.6 | -70.0 [-97.3, -42.8] |
| Ethnicity (collapsed) | East/SE Asian / Pacific | 10 | -27.0 | -40.4 [-88.3, 7.6] |
| Ethnicity (collapsed) | South Asian | 2 | -18.0 | -45.2 [-152.4, 62.0] |
| Ethnicity (collapsed) | Middle Eastern / N. African | 1 | -48.0 | -31.8 [-183.3, 119.7] |
| Ethnicity (collapsed) | Multiple | 20 | -57.0 | -59.6 [-93.5, -25.7] |
| Ethnicity (collapsed) | Unknown / Prefer not to say | 34 | -38.7 | -46.3 [-72.3, -20.3] |
| Ethnicity (collapsed) | Not answered | 14 | -49.1 | -56.0 [-96.5, -15.5] |

**TABLE S14.** Pooled change in IBS-SSS by subgroup (first reading at 14 days or later). RTM-corrected change is centred at 171.42.

| Subgroup | Stratum | n | Unadjusted change | RTM-corrected change<br>[95% CI] |
| --- | --- | --- | --- | --- |
| Age stratum (years) | 18-34 | 142 | -26.7 | -22.3 [-36.3, -8.4] |
| Age stratum (years) | 35-49 | 607 | -43.3 | -36.5 [-43.3, -29.8] |
| Age stratum (years) | 50-64 | 814 | -40.9 | -43.9 [-49.7, -38.0] |
| Age stratum (years) | >=65 | 112 | -42.8 | -62.8 [-78.5, -47.0] |
| Age stratum (years) | Unknown/not answered | 8 | -13.6 | -16.2 [-74.8, 42.5] |
| BMI stratum (kg/m2) | <25 | 92 | -33.8 | -18.8 [-36.3, -1.4] |
| BMI stratum (kg/m2) | 25-29.9 | 373 | -39.8 | -40.4 [-49.0, -31.8] |
| BMI stratum (kg/m2) | 30-34.9 | 510 | -42.3 | -42.1 [-49.5, -34.8] |
| BMI stratum (kg/m2) | >=35 | 687 | -40.6 | -42.5 [-48.9, -36.2] |
| BMI stratum (kg/m2) | Unknown/not answered | 21 | -37.0 | -33.3 [-69.6, 3.0] |
| Sex | Female | 1,345 | -41.3 | -39.0 [-43.6, -34.5] |
| Sex | Male | 262 | -36.8 | -51.9 [-62.2, -41.5] |
| Sex | Unknown/not answered | 76 | -40.4 | -28.2 [-47.3, -9.0] |
| Household income | <= \$35,000 | 80 | -56.0 | -48.6 [-67.2, -29.9] |
| Household income | \$35,001-\$65,000 | 387 | -38.1 | -34.1 [-42.6, -25.6] |
| Household income | \$65,001-\$110,000 | 488 | -41.2 | -38.6 [-46.2, -31.1] |
| Household income | \$110,001-\$175,000 | 287 | -48.4 | -49.1 [-58.9, -39.3] |
| Household income | \$175,001-\$450,000 | 165 | -31.8 | -42.2 [-55.2, -29.2] |
| Household income | >= \$450,001 | 10 | -12.4 | -39.0 [-91.7, 13.8] |
| Household income | Prefer not to say | 208 | -39.5 | -42.9 [-54.5, -31.4] |
| Household income | Not answered | 58 | -24.1 | -33.3 [-55.2, -11.4] |
| Ethnicity (collapsed) | White | 915 | -40.3 | -39.3 [-44.8, -33.8] |
| Ethnicity (collapsed) | Hispanic / Latino | 244 | -49.8 | -51.3 [-61.9, -40.6] |
| Ethnicity (collapsed) | Black / African American | 154 | -41.0 | -41.2 [-54.6, -27.8] |
| Ethnicity (collapsed) | East/SE Asian / Pacific | 40 | -23.4 | -42.2 [-68.6, -15.8] |
| Ethnicity (collapsed) | South Asian | 24 | -16.9 | -34.7 [-68.7, -0.7] |
| Ethnicity (collapsed) | Middle Eastern / N. African | 6 | 57.8 | 48.5 [-19.4, 116.4] |
| Ethnicity (collapsed) | Multiple | 76 | -34.4 | -31.0 [-50.1, -12.0] |
| Ethnicity (collapsed) | Unknown / Prefer not to say | 167 | -48.4 | -42.5 [-55.4, -29.6] |
| Ethnicity (collapsed) | Not answered | 57 | -20.4 | -30.5 [-52.5, -8.4] |

**TABLE S15.** Omnibus tests of whether RTM-corrected IBS-SSS change differs between subgroup strata, at each landmark.

| Subgroup | 4 weeks | 12 weeks | 26 weeks | Pooled |
| --- | --- | --- | --- | --- |
| Age stratum (years) | F(4,331) = 0.67, p = 0.614 | F(4,424) = 0.91, p = 0.458 | F(4,339) = 2.37, p = 0.052 | F(4,1677) = 4.32, p = 0.002 |
| BMI stratum (kg/m2) | F(4,331) = 0.95, p = 0.436 | F(4,424) = 1.16, p = 0.328 | F(4,339) = 2.88, p = 0.023 | F(4,1677) = 1.67, p = 0.154 |
| Sex | F(2,333) = 1.82, p = 0.164 | F(2,426) = 0.29, p = 0.749 | F(2,341) = 0.34, p = 0.710 | F(2,1679) = 3.28, p = 0.038 |
| Household income | F(7,328) = 0.50, p = 0.834 | F(7,421) = 0.93, p = 0.486 | F(6,337) = 0.50, p = 0.811 | F(7,1674) = 0.97, p = 0.455 |
| Ethnicity (collapsed) | F(8,327) = 1.20, p = 0.300 | F(7,421) = 1.49, p = 0.170 | F(8,335) = 0.30, p = 0.965 | F(8,1673) = 1.59, p = 0.124 |

**TABLE S16.** Weight (lb) and ND-score change. Upper block, weight by responder status (responders vs non-responders, symptomatic members). Lower blocks, all respondents with a paired reading (paired t test).

| Landmark | n responders / non-responders | Responders: mean change [95% CI] | Non-responders: mean change [95% CI] | Difference [95% CI] | p |
| --- | --- | --- | --- | --- | --- |
| 4 weeks | 117 / 113 | -2.39 [-4.08, -0.70] | -3.24 [-5.42, -1.06] | +0.85 [-1.89, +3.59] | 0.5427 |
| 12 weeks | 140 / 121 | -7.60 [-10.07, -5.13] | -5.82 [-8.17, -3.46] | -1.78 [-5.18, +1.62] | 0.3031 |
| 26 weeks | 143 / 83 | -12.48 [-15.37, -9.59] | -9.87 [-14.41, -5.32] | -2.61 [-7.97, +2.74] | 0.3367 |

| Measure | Landmark | Paired n | Mean change [95% CI] | Mean % change | p |
| --- | --- | --- | --- | --- | --- |
| Weight (lb) | 4 weeks | 1,564 | -3.53 [-4.01, -3.04] | -1.63 | <0.0001 |
| Weight (lb) | 12 weeks | 1,395 | -6.66 [-7.40, -5.93] | -3.02 | <0.0001 |
| Weight (lb) | 26 weeks | 1,153 | -10.34 [-11.40, -9.28] | -4.61 | <0.0001 |
| ND-score (points) | 4 weeks | 1,313 | +0.47 [+0.37, +0.56] | not applicable | <0.0001 |
| ND-score (points) | 12 weeks | 812 | +0.69 [+0.57, +0.81] | not applicable | <0.0001 |
| ND-score (points) | 26 weeks | 471 | +0.70 [+0.53, +0.88] | not applicable | <0.0001 |

**TABLE S17.**
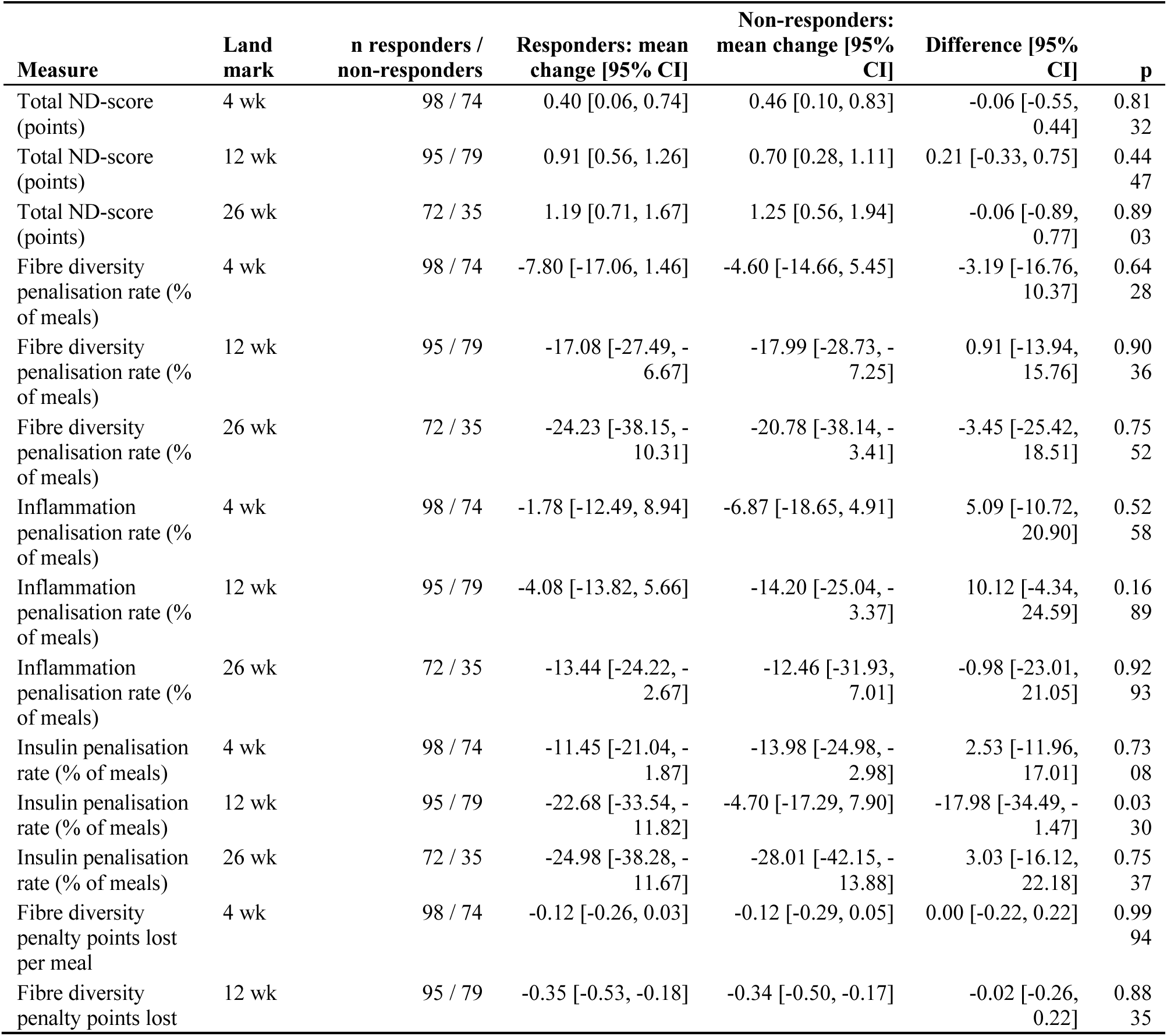
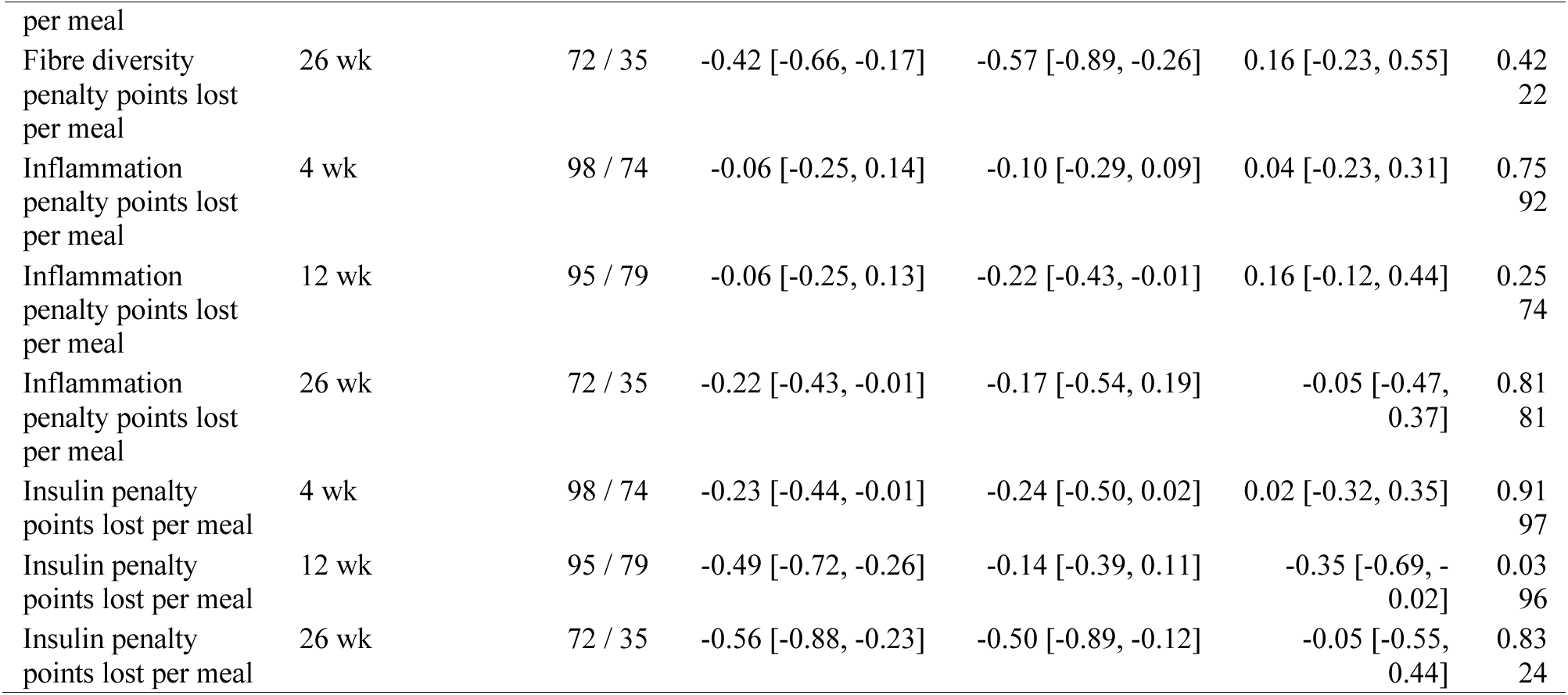
Change in ND-score and its penalisation domains from baseline, by responder status. Rates are percentages of meals; penalty points are points lost per meal.

**TABLE S18.** Within-member and between-member associations of weight and ND-score with IBS-SSS (respondents; occasions matched within 14 days; same-day mean; cluster-robust CR2 errors). Coefficients are IBS-SSS points per lb, per ND point, per 10 percentage points of meals penalised, or per penalty point lost per meal. Domain models are fitted one at a time; the last three rows fit the three penalisation rates jointly.

| Exposure | Observations / members | Within-member coefficient [95% CI] | p | Between-member coefficient [95% CI] | p | Contrast p |
| --- | --- | --- | --- | --- | --- | --- |
| Weight (per lb) | 1,841 / 1,319 | 0.236 [-0.295, 0.768] | 0.361<br>0 | -0.005 [-0.098, 0.088] | 0.923<br>0 | 0.3580 |
| ND-score (per point) | 1,379 / 1,048 | -6.047 [-11.018, -1.075] | 0.017<br>8 | -4.676 [-8.264, -1.088] | 0.010<br>8 | 0.6628 |
| Fibre-diversity penalisation rate | 1,379 / 1,048 | 1.065 [-0.643, 2.772] | 0.218<br>6 | 1.156 [-0.117, 2.429] | 0.075<br>1 | 0.9326 |
| Inflammation penalisation rate | 1,379 / 1,048 | 2.481 [0.656, 4.306] | 0.008<br>4 | 1.200 [-0.174, 2.575] | 0.086<br>8 | 0.2744 |
| Insulin penalisation rate | 1,379 / 1,048 | -0.211 [-1.873, 1.452] | 0.801<br>7 | 1.058 [-0.190, 2.306] | 0.096<br>5 | 0.2261 |
| Fibre-diversity penalty points per meal | 1,379 / 1,048 | 7.817 [-4.888, 20.522] | 0.224<br>4 | 8.578 [0.446, 16.710] | 0.038<br>7 | 0.9212 |
| Inflammation penalty points per meal | 1,379 / 1,048 | 14.038 [4.609, 23.466] | 0.004<br>2 | 4.250 [-2.594, 11.095] | 0.222<br>6 | 0.1034 |
| Insulin penalty points per meal | 1,379 / 1,048 | 1.282 [-5.940, 8.503] | 0.724<br>1 | 4.798 [-0.782, 10.378] | 0.091<br>7 | 0.4451 |
| Joint model: fibre-diversity rate | 1,379 / 1,048 | 0.824 [-0.897, 2.545] | 0.343<br>7 | 1.006 [-0.268, 2.280] | 0.121<br>4 | 0.8665 |
| Joint model: inflammation rate | 1,379 / 1,048 | 2.367 [0.533, 4.201] | 0.012<br>1 | 0.973 [-0.414, 2.360] | 0.168<br>8 | 0.2365 |
| Joint model: insulin rate | 1,379 / 1,048 | -0.239 [-1.873, 1.394] | 0.771<br>6 | 0.864 [-0.389, 2.117] | 0.176<br>0 | 0.2860 |

**TABLE S19.**
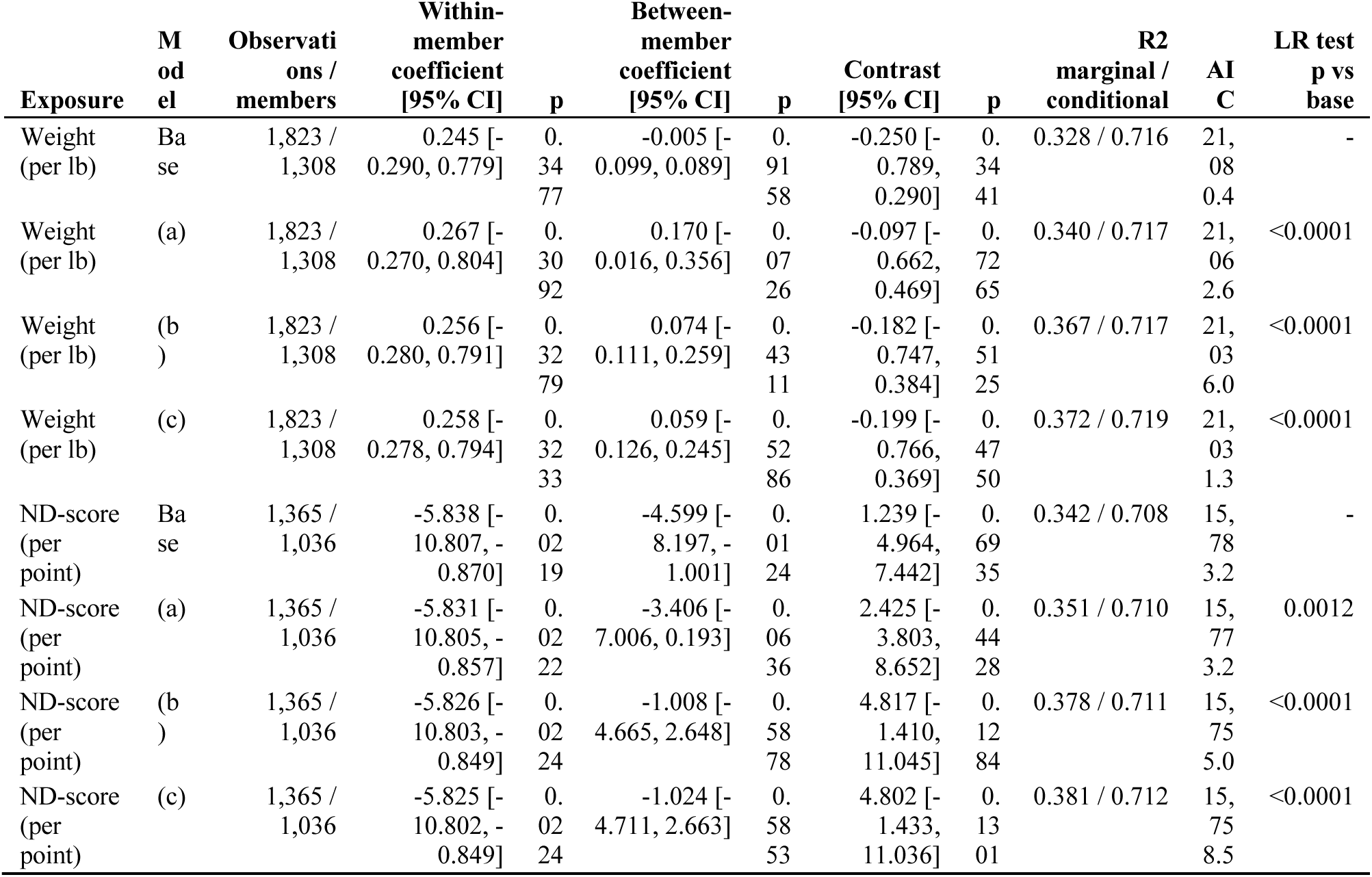
Covariate-extended within-member and between-member models of IBS-SSS for weight and ND-score (respondents; one common occasion sample per exposure, so that models are nested on identical data). Base: baseline IBS-SSS score, time, within and between exposure, member random intercept. (a) adds age, sex and baseline BMI; (b) adds ethnicity, household-income stratum and engagement days; (c) adds baseline IBS-SSS stratum and intake-list medication. Coefficients are IBS-SSS points per lb (weight) or per ND point; contrast is between minus within. Cluster-robust CR2 errors. R2, marginal and conditional (REML); AIC and likelihood-ratio test from maximum-likelihood refits; the test compares each model with the base. Associations only.

| Exposure | Model | Observations / members | Within-member coefficient [95% CI] | p | Between-member coefficient [95% CI] | p | Contrast [95% CI] | p | R2 marginal / conditional | AIC | LR test p vs base |
| --- | --- | --- | --- | --- | --- | --- | --- | --- | --- | --- | --- |
| Weight (per lb) | Base | 1,823 / 1,308 | 0.245 [-0.290, 0.779] | 0.3477 | -0.005 [-0.099, 0.089] | 0.9158 | -0.250 [-0.789, 0.290] | 0.3441 | 0.328 / 0.716 | 21,080.4 | - |
| Weight (per lb) | (a) | 1,823 / 1,308 | 0.267 [-0.270, 0.804] | 0.3092 | 0.170 [-0.016, 0.356] | 0.0726 | -0.097 [-0.662, 0.469] | 0.7265 | 0.340 / 0.717 | 21,062.6 | <0.0001 |
| Weight (per lb) | (b) | 1,823 / 1,308 | 0.256 [-0.280, 0.791] | 0.3279 | 0.074 [-0.111, 0.259] | 0.4311 | -0.182 [-0.747, 0.384] | 0.5125 | 0.367 / 0.717 | 21,036.0 | <0.0001 |
| Weight (per lb) | (c) | 1,823 / 1,308 | 0.258 [-0.278, 0.794] | 0.3233 | 0.059 [-0.126, 0.245] | 0.5286 | -0.199 [-0.766, 0.369] | 0.4750 | 0.372 / 0.719 | 21,031.3 | <0.0001 |
| ND-score (per point) | Base | 1,365 / 1,036 | -5.838 [-10.807, -0.870] | 0.0219 | -4.599 [-8.197, -1.001] | 0.0124 | 1.239 [-4.964, 7.442] | 0.6935 | 0.342 / 0.708 | 15,783.2 | - |
| ND-score (per point) | (a) | 1,365 / 1,036 | -5.831 [-10.805, -0.857] | 0.0222 | -3.406 [-7.006, 0.193] | 0.0636 | 2.425 [-3.803, 8.652] | 0.4428 | 0.351 / 0.710 | 15,773.2 | 0.0012 |
| ND-score (per point) | (b) | 1,365 / 1,036 | -5.826 [-10.803, -0.849] | 0.0224 | -1.008 [-4.665, 2.648] | 0.5878 | 4.817 [-1.410, 11.045] | 0.1284 | 0.378 / 0.711 | 15,755.0 | <0.0001 |
| ND-score (per point) | (c) | 1,365 / 1,036 | -5.825 [-10.802, -0.849] | 0.0224 | -1.024 [-4.711, 2.663] | 0.5853 | 4.802 [-1.433, 11.036] | 0.1301 | 0.381 / 0.712 | 15,758.5 | <0.0001 |

**TABLE S20.** Sensitivity summary: pooled and 12-week estimates of change in IBS-SSS (points; negative = improvement) without and with inverse-probability weighting and with medication terms added to the RTM-corrected model. RTM-corrected estimates are centred at 171.42.

| Analysis | n | Estimate [95% CI] | p |
| --- | --- | --- | --- |
| Pooled, unweighted, unadjusted | 1,592 | -40.50 [-45.25, -35.75] | <0.0001 |
| Pooled, unweighted, RTM-corrected | 1,592 | -41.04 [-45.24, -36.83] | <0.0001 |
| Pooled, IPW-A, unadjusted | 1,592 | -37.39 [-42.31, -32.47] | <0.0001 |
| Pooled, IPW-A, RTM-corrected | 1,592 | -39.48 [-43.82, -35.13] | <0.0001 |
| Pooled, IPW-B, unadjusted | 1,592 | -38.81 [-43.82, -33.79] | <0.0001 |
| Pooled, IPW-B, RTM-corrected | 1,592 | -39.12 [-43.51, -34.73] | <0.0001 |
| 12 weeks, RTM-corrected, antidepressant term added | 430 | -47.76 [-55.52, -39.99] | <0.0001 |
| 12 weeks, RTM-corrected, GLP-1 receptor agonist baseline term added | 430 | -47.87 [-55.62, -40.12] | <0.0001 |
| 12 weeks, RTM-corrected, antidepressant and GLP-1 terms added | 430 | -47.58 [-55.31, -39.85] | <0.0001 |
| 12 weeks, RTM-corrected, all medication terms added | 430 | -47.68 [-55.42, -39.94] | <0.0001 |

**TABLE S21.** Remission stratum as a negative control: unadjusted and RTM-corrected change, with the reliability-formula correction at the retest reliability (0.719, treated as fixed). Each 0.1 of reliability moves the pooled expected shift by 13.78 points.

| Reading set | n | Unadjusted change<br>[95% CI] | RTM-corrected change<br>[95% CI] | p | Reliability formula,<br>rho 0.719 [95% CI] |
| --- | --- | --- | --- | --- | --- |
| 12 weeks | 107 | +9.93 [-1.24, +21.09] | -37.49 [-63.57, -11.42] | 0.005 | -29.39 [-40.10, -18.67] |
| Pooled, first reading at<br>14 days or later | 344 | +24.08 [+17.32, +30.84] | -19.12 [-38.71, +0.47] | 0.056 | -14.64 [-21.25, -8.02]<br>(expected shift +38.72) |

**TABLE S22.** Drop-one covariate analysis of RTM-corrected change (models without stratum terms; complete-case samples).

| Reading set | Model | n | RTM-corrected mean change<br>[95% CI] | Shift from full model |
| --- | --- | --- | --- | --- |
| 12 weeks | Full | 407 | -50.95 [-59.53, -42.38] | – |
| 12 weeks | Drop baseline score | 407 | -45.84 [-55.64, -36.03] | 5.12 |
| 12 weeks | Drop age | 407 | -50.77 [-59.33, -42.20] | 0.19 |
| 12 weeks | Drop sex | 407 | -52.11 [-59.91, -44.31] | -1.15 |
| 12 weeks | Drop baseline BMI | 407 | -50.97 [-59.54, -42.41] | -0.02 |
| Pooled | Full | 1,592 | -39.06 [-43.61, -34.52] | – |
| Pooled | Drop baseline score | 1,592 | -41.30 [-46.50, -36.11] | -2.24 |
| Pooled | Drop age | 1,592 | -38.84 [-43.42, -34.27] | 0.22 |
| Pooled | Drop sex | 1,592 | -41.06 [-45.22, -36.89] | -1.99 |
| Pooled | Drop baseline BMI | 1,592 | -39.06 [-43.61, -34.52] | -0.0 |

**TABLE S23.**
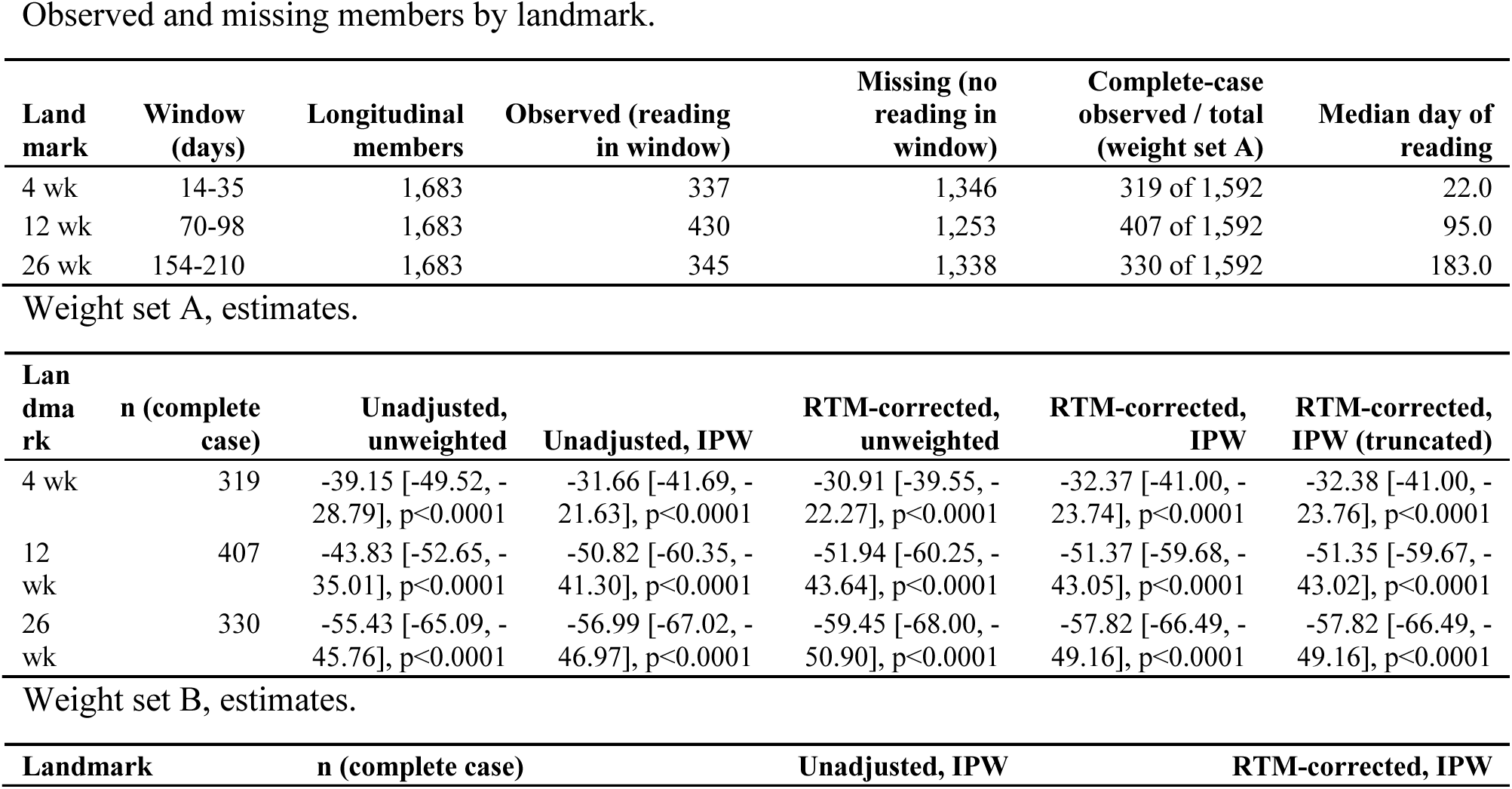
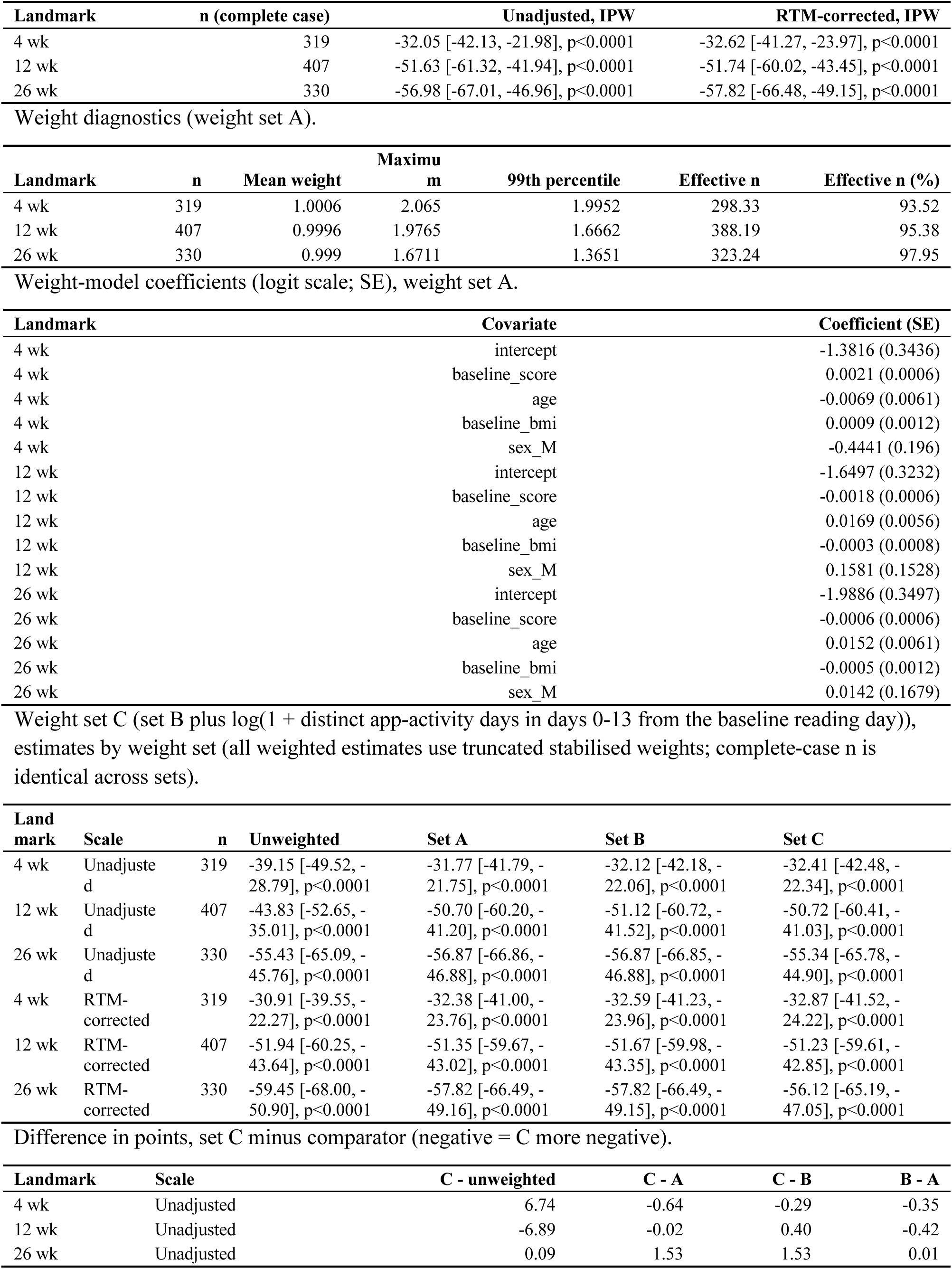
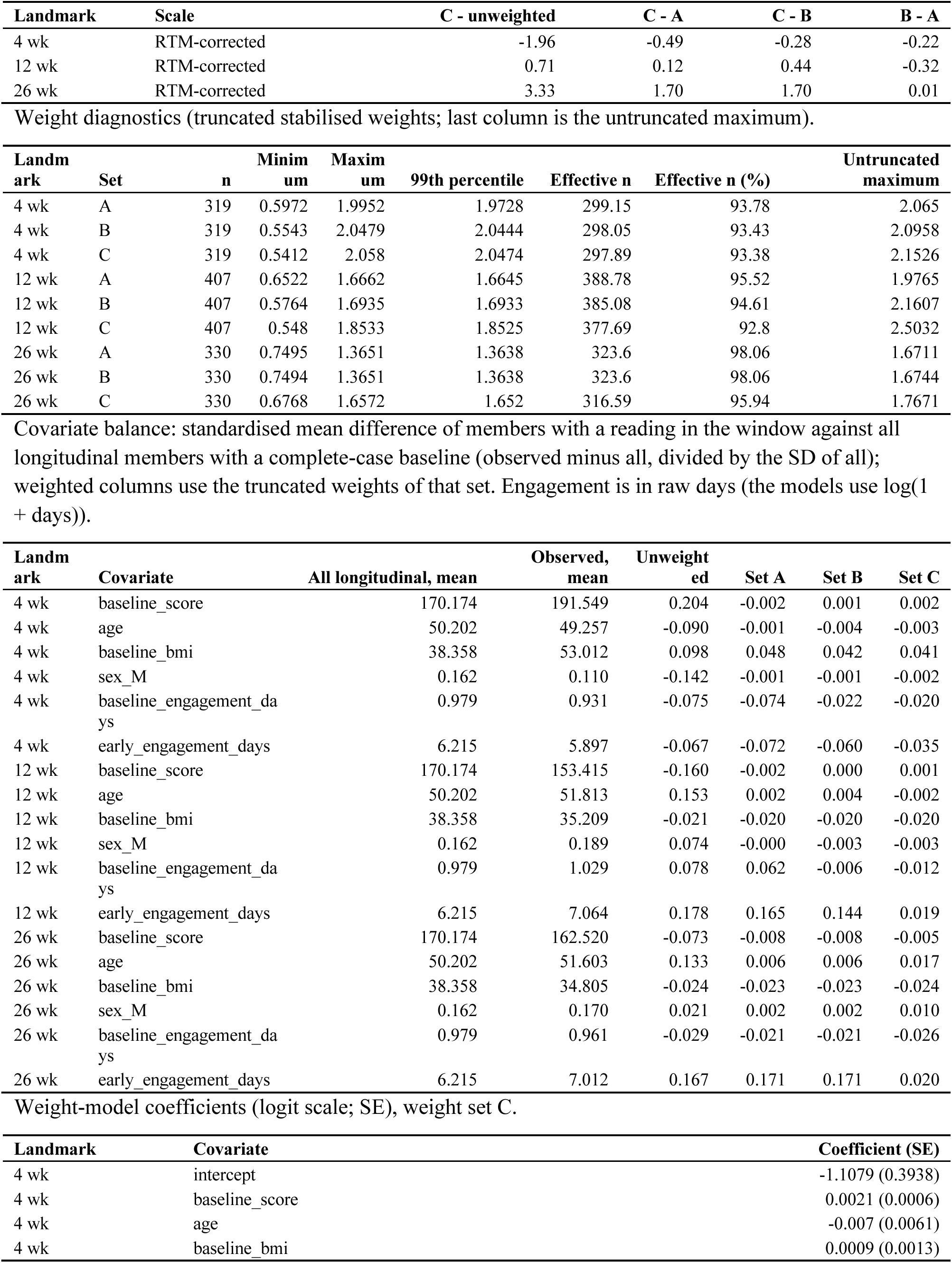
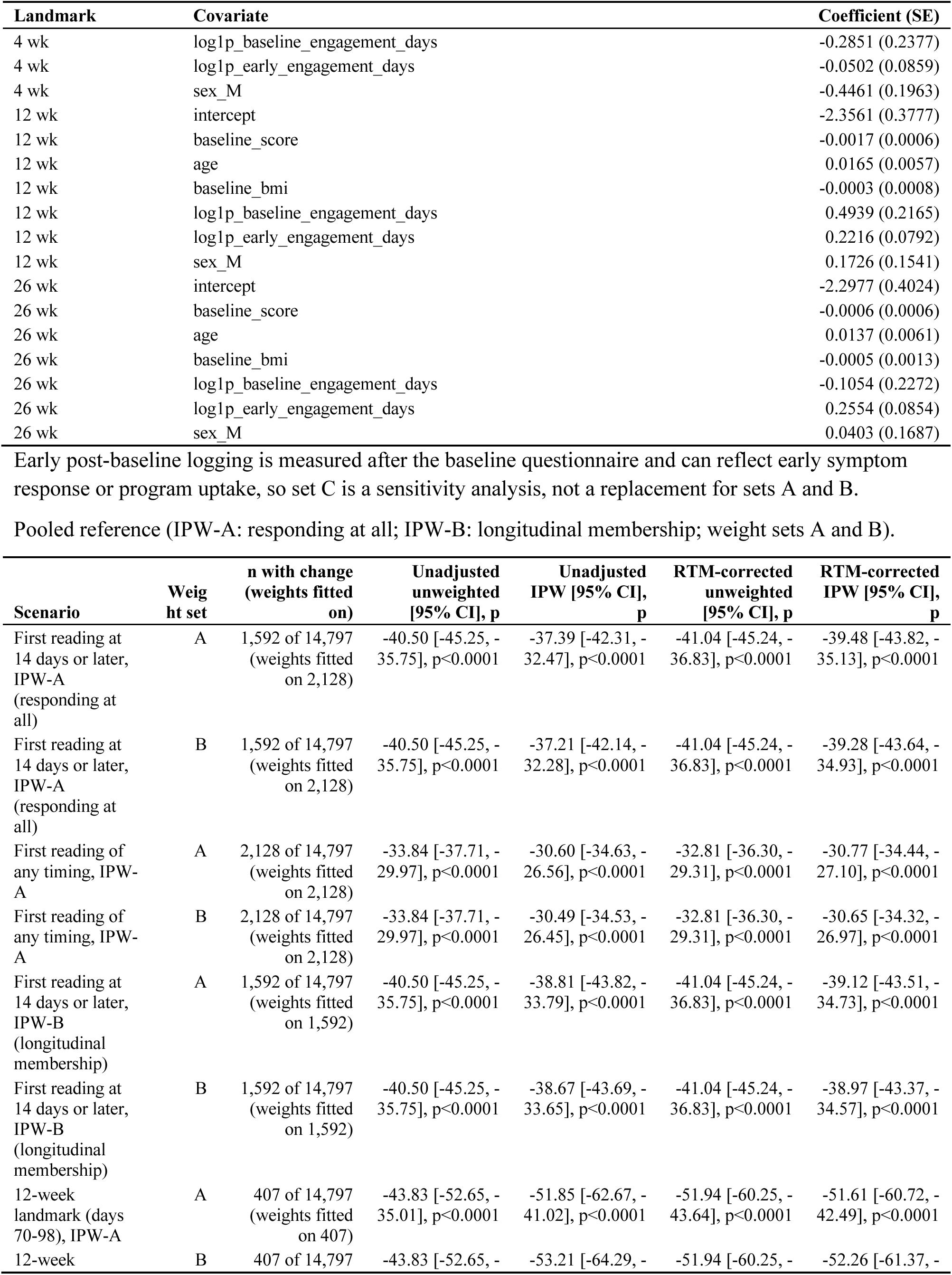
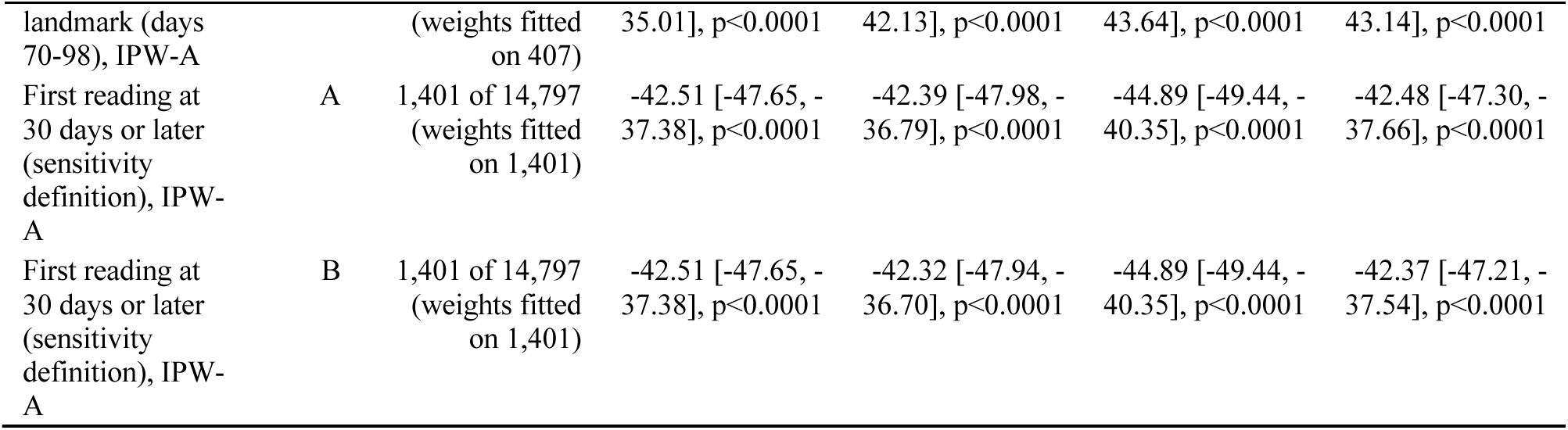
Inverse-probability weighting. Upper blocks: per-landmark analysis within the longitudinal sample (n = 1,683; complete-case 1,592), with the number observed in each landmark window, weighted and unweighted estimates, weight diagnostics and weight-model coefficients (weight set A; set B reported for IPW; set C, which adds early post-baseline engagement, in the final blocks of the per-landmark analysis). Lower block: pooled reference models for responding at all (IPW-A) and longitudinal membership (IPW-B) among 14,797 in-window members. Estimates are mean change [95% CI].

**TABLE S24.** Delta-based tipping-point sensitivity analysis for data missing not at random. Upper block: delta at which the mean change over all 1,683 longitudinal members reaches zero at each landmark, with the ratio to the 50-point benchmark. Middle block: pooled mean change by delta at each landmark (unadjusted; single imputation, so intervals are anti-conservative). Lower block: pooled reference from the original design, in which all 14,250 non-observed members were imputed the observed pooled mean (–40.54) plus the delta.

| Landmark | Observed n | Missing n | Observed mean, unadjusted | Delta to zero, unadjusted | Delta / 50-point benchmark | Observed mean, RTM-corrected | Delta to zero, RTM-corrected | Delta / 50-point benchmark | Zero reached within the benchmark (RTM-corrected) |
| --- | --- | --- | --- | --- | --- | --- | --- | --- | --- |
| 4 wk | 337 | 1,346 | -39.25 | 49.08 | 0.98 | -31.48 | 39.36 | 0.79 | yes |
| 12 wk | 430 | 1,253 | -43.88 | 58.93 | 1.18 | -51.47 | 69.13 | 1.38 | no |
| 26 wk | 345 | 1,338 | -54.69 | 68.79 | 1.38 | -58.29 | 73.31 | 1.47 | no |

Mean change by delta (unadjusted).
| Delta (points) | 4 weeks: pooled mean change [95% CI] | 4 weeks: p | 12 weeks: pooled mean change [95% CI] | 12 weeks: p | 26 weeks: pooled mean change [95% CI] | 26 weeks: p |
| --- | --- | --- | --- | --- | --- | --- |
| 0 | -39.25 [-41.25, -37.25] | 0.0000 | -43.88 [-46.06, -41.70] | 0.0000 | -54.69 [-56.60, -52.78] | 0.0000 |
| 5 | -35.25 [-37.26, -33.25] | 0.0000 | -40.15 [-42.34, -37.97] | 0.0000 | -50.71 [-52.63, -48.80] | 0.0000 |
| 10 | -31.25 [-33.27, -29.24] | 0.0000 | -36.43 [-38.62, -34.24] | 0.0000 | -46.74 [-48.66, -44.82] | 0.0000 |
| 15 | -27.25 [-29.28, -25.23] | 0.0000 | -32.71 [-34.91, -30.51] | 0.0000 | -42.76 [-44.70, -40.83] | 0.0000 |
| 20 | -23.26 [-25.30, -21.22] | 0.0000 | -28.99 [-31.21, -26.77] | 0.0000 | -38.79 [-40.74, -36.84] | 0.0000 |
| 25 | -19.26 [-21.32, -17.20] | 0.0000 | -25.26 [-27.51, -23.02] | 0.0000 | -34.81 [-36.79, -32.84] | 0.0000 |
| 30 | -15.26 [-17.34, -13.17] | 0.0000 | -21.54 [-23.81, -19.27] | 0.0000 | -30.84 [-32.84, -28.84] | 0.0000 |
| 35 | -11.26 [-13.37, -9.15] | 0.0000 | -17.82 [-20.12, -15.52] | 0.0000 | -26.86 [-28.89, -24.84] | 0.0000 |
| 40 | -7.26 [-9.41, -5.12] | 0.0000 | -14.10 [-16.43, -11.76] | 0.0000 | -22.89 [-24.95, -20.83] | 0.0000 |
| 45 | -3.26 [-5.44, -1.08] | 0.0034 | -10.37 [-12.75, -8.00] | 0.0000 | -18.91 [-21.01, -16.81] | 0.0000 |
| 50 | 0.74 [-1.48, 2.96] | 0.5149 | -6.65 [-9.07, -4.23] | 0.0000 | -14.94 [-17.08, -12.80] | 0.0000 |

Pooled reference.
| Delta (points) | Imputed change for non-observed members | Pooled mean change | 95% CI | p |
| --- | --- | --- | --- | --- |
| 0 | -40.54 | -40.54 | [-41.02, -40.05] | < 0.0001 |
| 5 | -35.54 | -36.06 | [-36.55, -35.58] | < 0.0001 |
| 10 | -30.54 | -31.59 | [-32.08, -31.11] | < 0.0001 |
| 15 | -25.54 | -27.12 | [-27.61, -26.63] | < 0.0001 |
| 20 | -20.54 | -22.65 | [-23.14, -22.16] | < 0.0001 |
| 25 | -15.54 | -18.18 | [-18.67, -17.68] | < 0.0001 |
| 30 | -10.54 | -13.71 | [-14.21, -13.20] | < 0.0001 |
| 35 | -5.54 | -9.23 | [-9.74, -8.72] | < 0.0001 |
| 40 | -0.54 | -4.76 | [-5.28, -4.24] | < 0.0001 |
| 45 | 4.46 | -0.29 | [-0.82, 0.24] | 0.2833 |
| 50 | 9.46 | 4.18 | [3.64, 4.72] | < 0.0001 |

**TABLE S25.** Change in IBS-SSS by antidepressant medication at baseline (intake list or pharmacy claims) and by GLP-1 receptor agonist use at baseline, RTM-corrected, by landmark.

| Exposure | Landmark (weeks) | n exposed / not exposed | RTM-corrected, exposed [95% CI] | RTM-corrected, not exposed [95% CI] | Difference [95% CI] | p |
| --- | --- | --- | --- | --- | --- | --- |
| Antidepressant | 4 | 112 / 225 | -29.66 [-46.07, -13.24] | -36.95 [-49.87, -24.03] | +5.64 [-13.30, +24.58] | 0.55<br>8 |
| Antidepressant | 12 | 106 / 324 | -48.54 [-63.46, -33.61] | -52.73 [-63.63, -41.83] | +3.76 [-13.71, +21.24] | 0.67<br>2 |
| Antidepressant | 26 | 101 / 244 | -65.05 [-79.42, -50.67] | -55.48 [-66.72, -44.23] | -10.91 [-28.85, +7.02] | 0.23<br>2 |
| GLP-1 receptor agonist at baseline | 4 | 47 / 290 | -60.14 [-102.51, -17.77] | -31.48 [-41.65, -21.31] | -9.02 [-34.85, +16.81] | 0.49<br>3 |
| GLP-1 receptor agonist at baseline | 12 | 64 / 366 | -45.32 [-74.99, -15.65] | -52.37 [-61.42, -43.32] | +4.69 [-16.51, +25.89] | 0.66<br>4 |
| GLP-1 receptor agonist at baseline | 26 | 71 / 274 | -72.87 [-89.23, -56.51] | -55.14 [-64.65, -45.62] | -19.92 [-40.01, +0.17] | 0.05<br>2 |

**TABLE S26.**
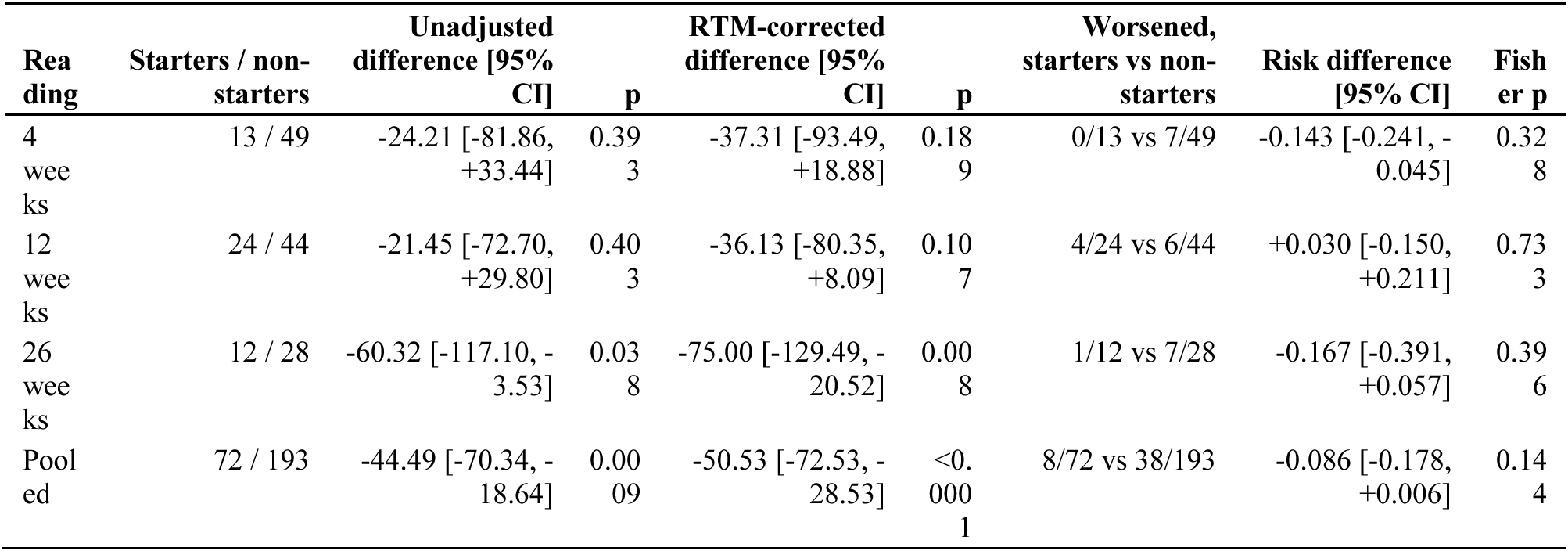
GLP-1 receptor agonist starters versus non-starters (members who were not baseline users and had a complete pharmacy window): change in IBS-SSS and worsening by 50 points or more. Differences are starters minus non-starters.

**TABLE S27.**
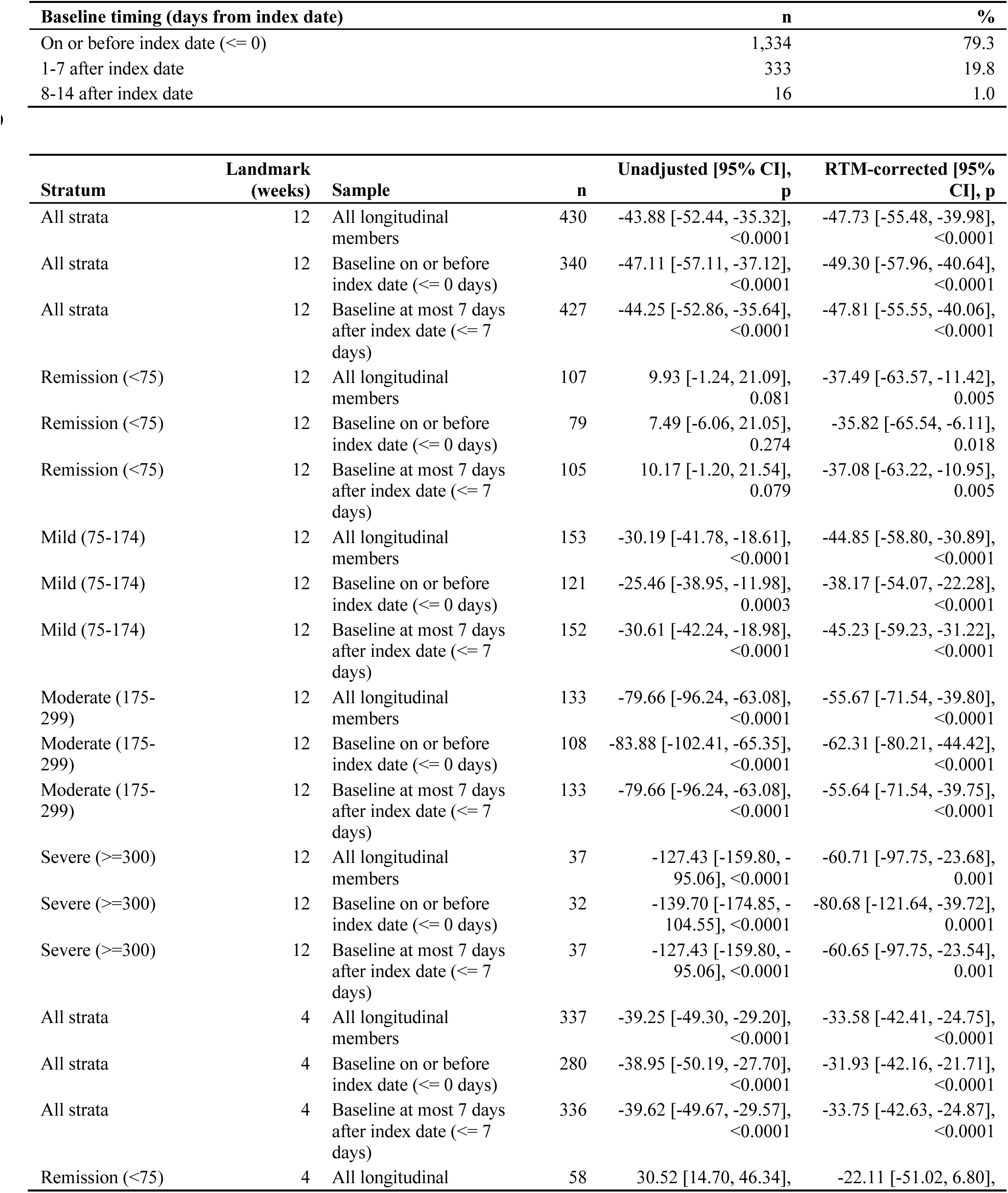
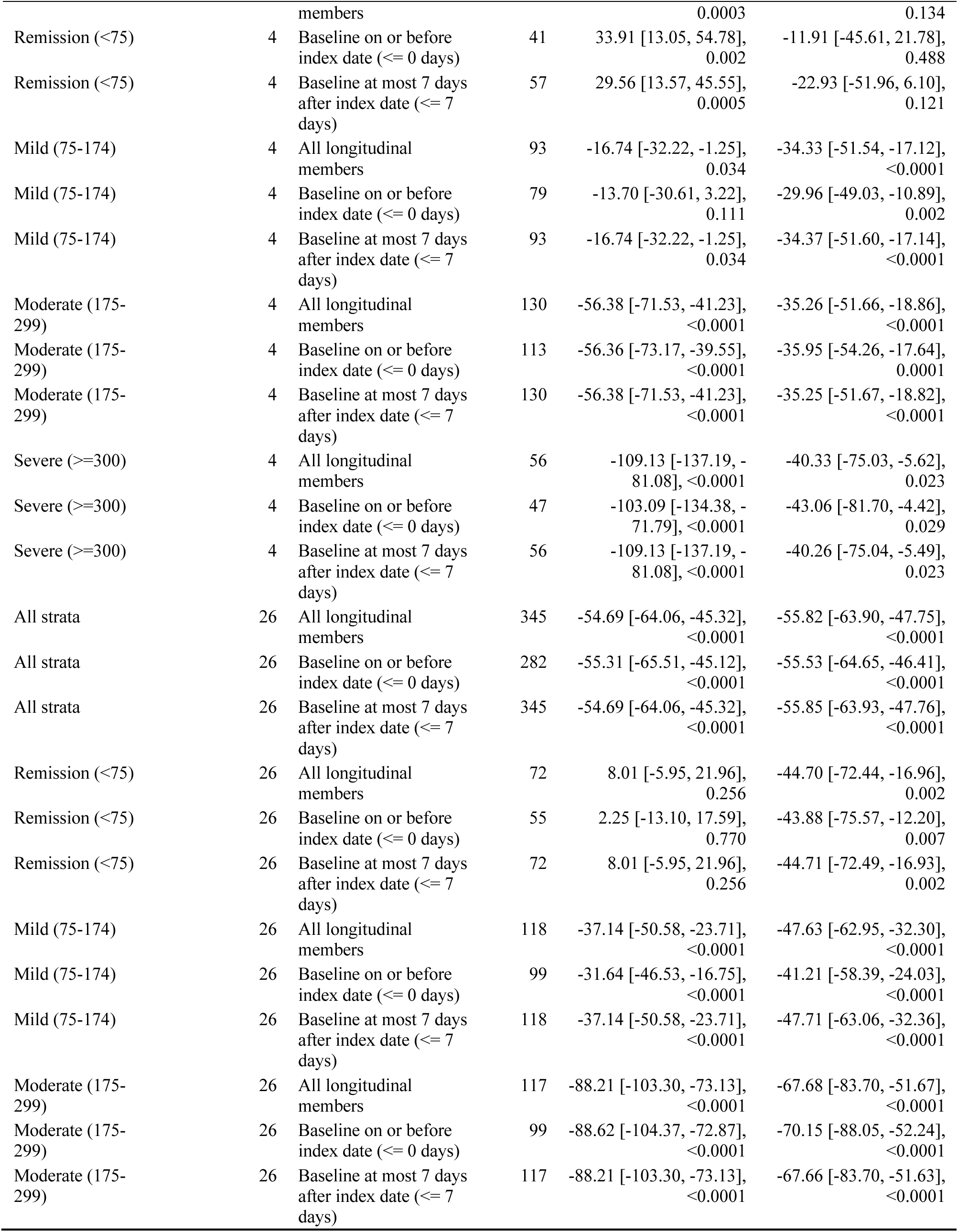
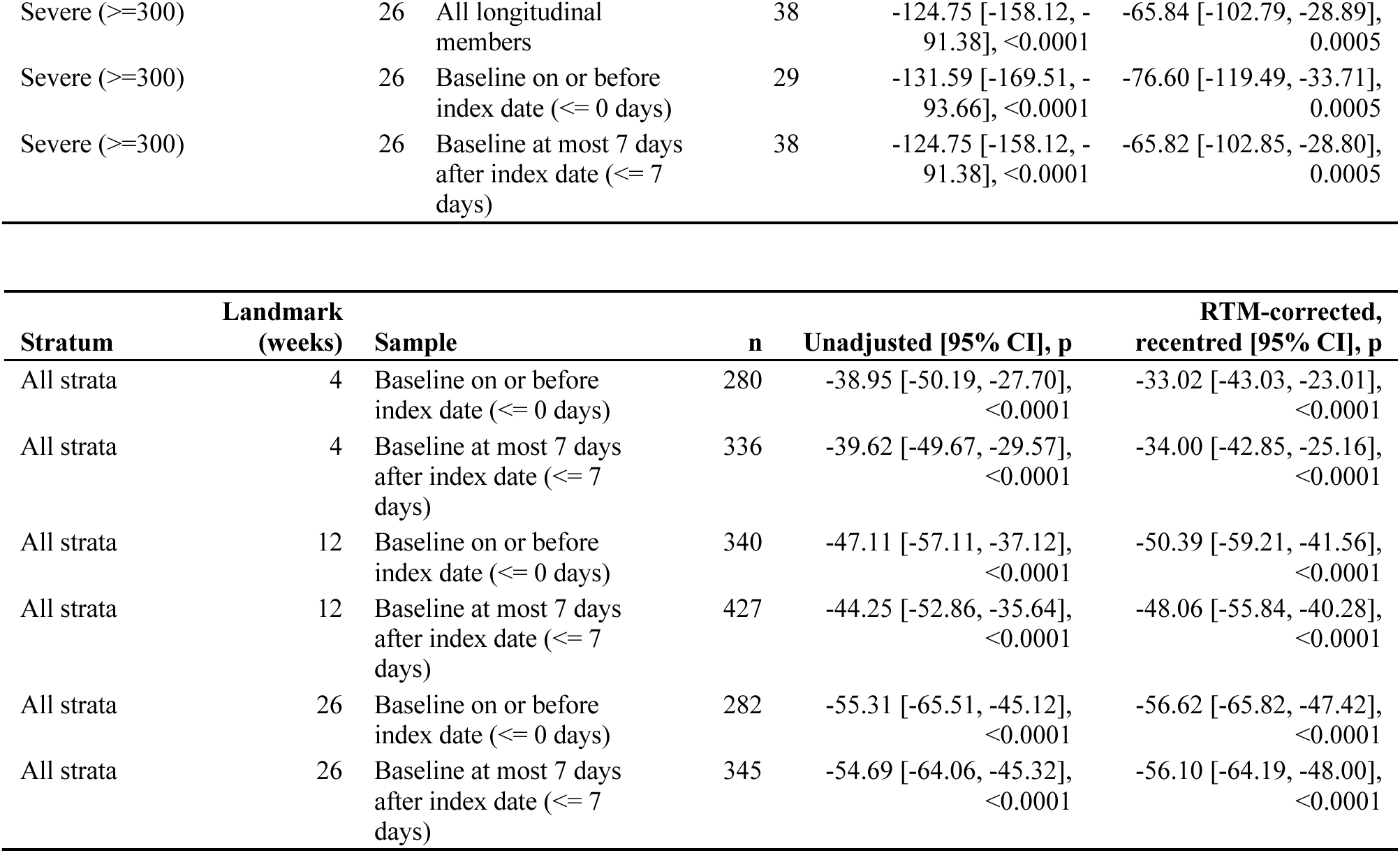
Baseline timing relative to the index date and landmark changes in IBS-SSS (points; negative = improvement) in the longitudinal sample and in members whose baseline reading was on or before the index date or at most 7 days after it. Landmark windows were defined from each member’s own baseline reading. RTM-corrected estimates are centred at the full longitudinal-sample mean (171.42). Upper block: baseline timing among the 1,683 longitudinal members (median 0, interquartile range 0 to 0 days). Middle block: change by stratum and landmark. Lower block: all strata, RTM-corrected estimates recentred at each restricted sample’s own mean baseline (full sample 171.42; <= 0 days 174.73; <= 7 days 172.11).

**TABLE S28.**
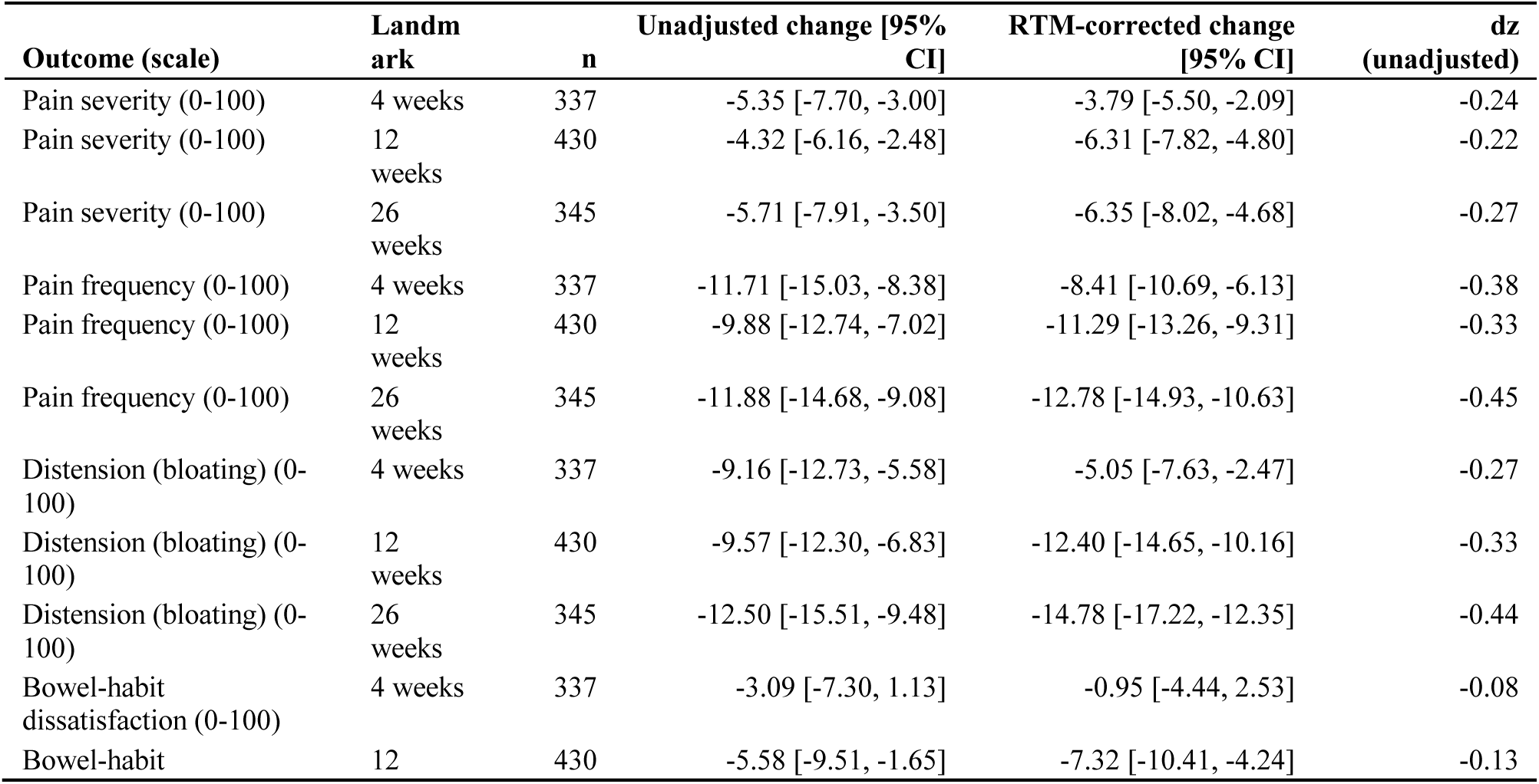
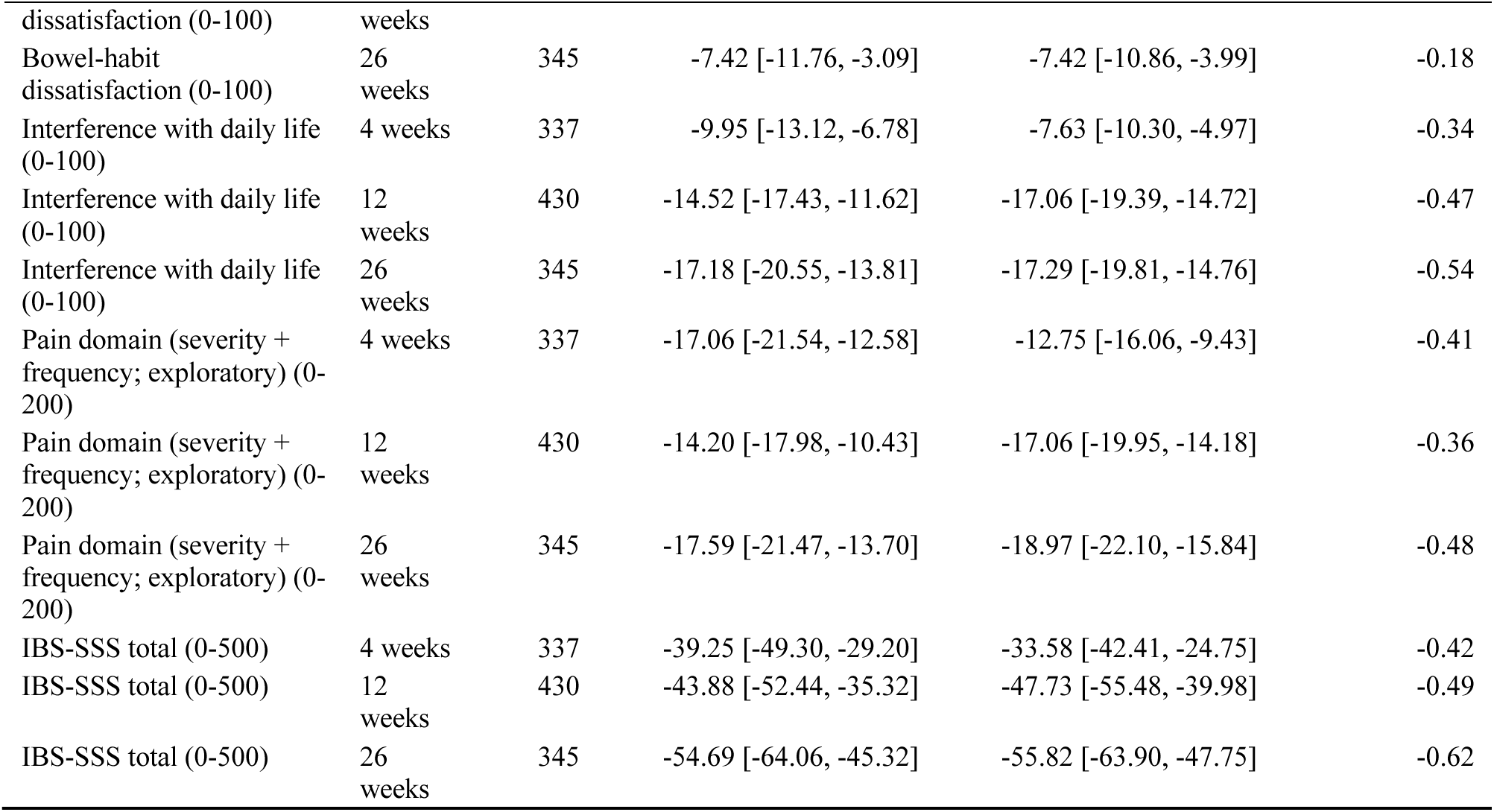
Change in IBS-SSS by item and domain at 4, 12 and 26 weeks (points; negative = improvement). Each item is scored 0-100; the pain domain (severity plus frequency) is an exploratory grouping. RTM-corrected estimates are centred at each outcome’s longitudinal-sample mean baseline (total 171.42). dz is the unadjusted mean change divided by the standard deviation of change. Exploratory; no multiplicity correction.

**TABLE S29.** Continuous-time (smooth model) change from baseline by item at 4, 12 and 26 weeks (days 28, 84 and 182), unadjusted and RTM-corrected (longitudinal sample, n = 1,683 members, 2,342 follow-up readings; points, negative = improvement).

| Outcome (scale) | Week (day) | Unadjusted, smooth [95% CI] | RTM-corrected, smooth [95% CI] |
| --- | --- | --- | --- |
| Pain severity (0-100) | 4 (28) | -4.1 [-5.3, -2.8] | -4.2 [-5.2, -3.2] |
| Pain severity (0-100) | 12 (84) | -4.2 [-5.3, -3.2] | -4.3 [-5.2, -3.5] |
| Pain severity (0-100) | 26 (182) | -4.5 [-5.6, -3.3] | -4.5 [-5.4, -3.7] |
| Pain frequency (0-100) | 4 (28) | -10.0 [-11.6, -8.3] | -8.7 [-10.3, -7.0] |
| Pain frequency (0-100) | 12 (84) | -9.9 [-11.3, -8.4] | -9.8 [-11.0, -8.6] |
| Pain frequency (0-100) | 26 (182) | -9.6 [-11.1, -8.1] | -10.8 [-12.2, -9.4] |
| Distension (bloating) (0-100) | 4 (28) | -8.7 [-10.5, -6.9] | -6.6 [-8.6, -4.6] |
| Distension (bloating) (0-100) | 12 (84) | -9.5 [-11.0, -8.0] | -10.0 [-11.4, -8.6] |
| Distension (bloating) (0-100) | 26 (182) | -11.0 [-12.6, -9.4] | -12.4 [-14.1, -10.7] |
| Bowel-habit dissatisfaction (0-100) | 4 (28) | -3.7 [-6.1, -1.4] | -3.5 [-5.5, -1.4] |
| Bowel-habit dissatisfaction (0-100) | 12 (84) | -4.5 [-6.5, -2.6] | -4.6 [-6.2, -3.1] |
| Bowel-habit dissatisfaction (0-100) | 26 (182) | -5.9 [-7.9, -3.9] | -6.5 [-8.2, -4.9] |
| Interference with daily life (0-100) | 4 (28) | -11.0 [-13.0, -8.9] | -9.9 [-11.8, -7.9] |
| Interference with daily life (0-100) | 12 (84) | -12.6 [-14.2, -11.1] | -12.9 [-14.3, -11.6] |
| Interference with daily life (0-100) | 26 (182) | -14.8 [-16.6, -13.0] | -15.7 [-17.4, -14.0] |
| Pain domain (severity + frequency; exploratory) (0-200) | 4 (28) | -14.2 [-16.5, -11.9] | -12.8 [-15.2, -10.3] |
| Pain domain (severity + frequency; exploratory) (0-200) | 12 (84) | -14.1 [-16.1, -12.1] | -14.2 [-15.9, -12.4] |
| Pain domain (severity + frequency; exploratory) (0-200) | 26 (182) | -14.0 [-16.0, -11.9] | -15.3 [-17.4, -13.2] |
| IBS-SSS total (0-500) | 4 (28) | -39.0 [-44.3, -33.6] | -32.8 [-39.1, -26.5] |
| IBS-SSS total (0-500) | 12 (84) | -41.0 [-45.6, -36.4] | -41.6 [-46.0, -37.2] |
| IBS-SSS total (0-500) | 26 (182) | -44.5 [-49.3, -39.8] | -49.5 [-54.9, -44.1] |

**TABLE S30.** Exploratory timing descriptors of the RTM-corrected smooth curve by item. Early share is the fitted change at 4 weeks divided by the fitted change at 26 weeks; the day to 75% is the first day at which 75% of the 26-week change was reached (14 is the first observable day). Intervals are member-level bootstrap percentile intervals (400 replicates). No pairwise difference between items in early share or in the day to 75% had a 95% interval excluding zero (10 comparisons each, no multiplicity adjustment). Item-by-time test over all five items: likelihood-ratio chi-square(44) = 50.7, p = 0.227; cluster-robust Wald chi-square(44) = 40.6, p = 0.617.

| Outcome | Fitted 26-week change (RTM-corrected) | Early share, 4 wk / 26 wk [bootstrap 95% CI] | Day to 75% of 26-week change [bootstrap 95% CI] |
| --- | --- | --- | --- |
| Pain severity | -4.5 [-6.3, -3.4] | 0.92 [0.40, 1.14] | 14 (first observable day) [14, 102] |
| Pain frequency | -10.8 [-13.0, -9.1] | 0.81 [0.59, 1.05] | 14 (first observable day) [14, 74] |
| Distension (bloating) | -12.4 [-14.1, -10.6] | 0.53 [0.30, 0.78] | 72 [14, 93] |
| Bowel-habit dissatisfaction | -6.5 [-9.5, -4.4] | 0.53 [-0.09, 0.80] | 97 [14, 133] |
| Interference with daily life | -15.7 [-17.8, -13.9] | 0.63 [0.46, 0.79] | 62 [14, 86] |
| Pain domain (severity + frequency; exploratory) | -15.3 [-18.5, -12.9] | 0.83 [0.57, 1.06] | 14 (first observable day) [14, 86] |
| IBS-SSS total | -49.5 [-57.3, -42.9] | 0.66 [0.50, 0.84] | 55 [14, 83] |

**TABLE S31.**
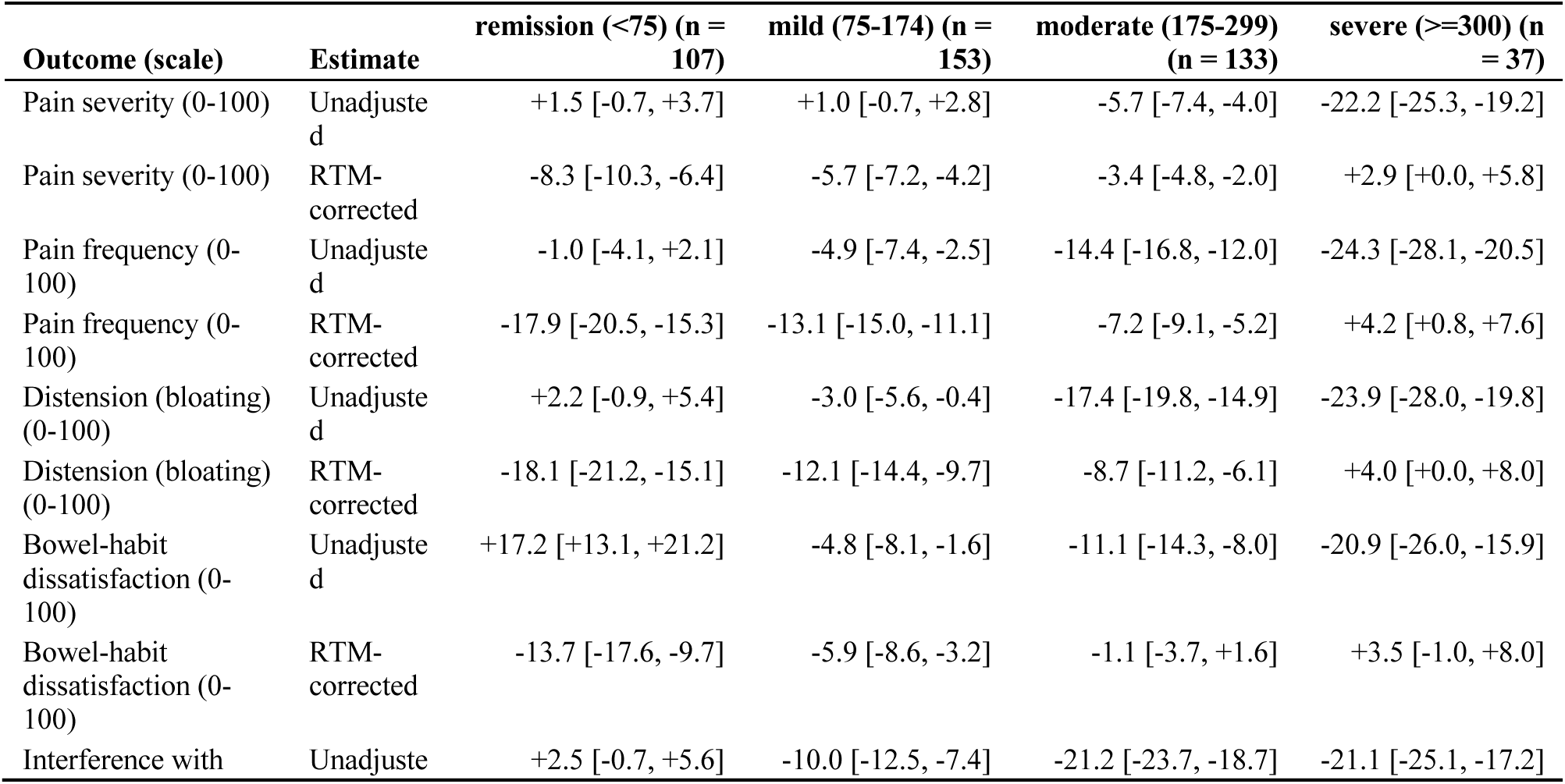
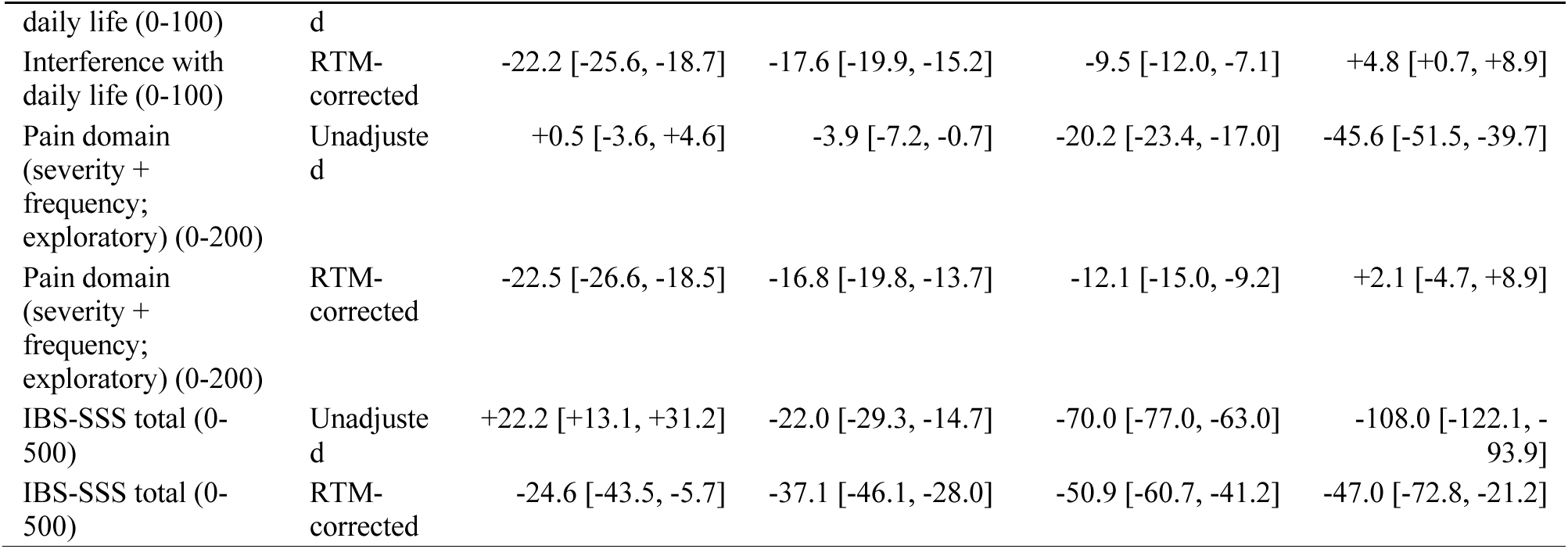
Smooth-model change from baseline at 12 weeks by item and baseline severity stratum, unadjusted and RTM-corrected (points; negative = improvement), with the number of members contributing in the 12-week window. RTM-corrected values are evaluated at the common baseline of the longitudinal sample, so that they extrapolate to a baseline far from the mean of the remission and severe strata; positive RTM-corrected values in the severe stratum are not evidence of worsening. Fitted values at 4 and 26 weeks are in the analysis tables of the item analysis.

**TABLE S32.** Tests of whether the profiles of the five items differ over time within each baseline severity stratum (change as a percentage of each item’s stratum baseline mean, RTM-corrected). Likelihood-ratio test and cluster-robust Wald test of the item-by-time interaction, with monthly or quarterly time bins. The cluster-robust test is not reliable when time cells hold few members (smallest cell shown) and is uninformative where it disagrees sharply with the likelihood-ratio test. No multiplicity adjustment.

| Stratum | Members | Time bins | Smallest cell (members) | Likelihood-ratio chi-square (df), p | Cluster-robust Wald chi-square (df), p |
| --- | --- | --- | --- | --- | --- |
| remission (<75) | 344 | monthly | 7 | 43.6 (44), 0.489 | 69.7 (44), 0.008 |
| remission (<75) | 344 | quarterly | 33 | 7.3 (12), 0.836 | 7.0 (12), 0.859 |
| mild (75-174) | 551 | monthly | 10 | 45.9 (44), 0.395 | 100.2 (44), <0.001 |
| mild (75-174) | 551 | quarterly | 58 | 9.2 (12), 0.685 | 29.0 (12), 0.004 |
| moderate (175-299) | 566 | monthly | 17 | 56.7 (44), 0.094 | 54.4 (44), 0.136 |
| moderate (175-299) | 566 | quarterly | 62 | 9.3 (12), 0.680 | 10.9 (12), 0.541 |
| severe (≥300) | 222 | monthly | 2 | 22.3 (44), 0.997 | 284.5 (44), <0.001 |
| severe (≥300) | 222 | quarterly | 30 | 4.1 (12), 0.982 | 4.4 (12), 0.974 |

**TABLE S33.**
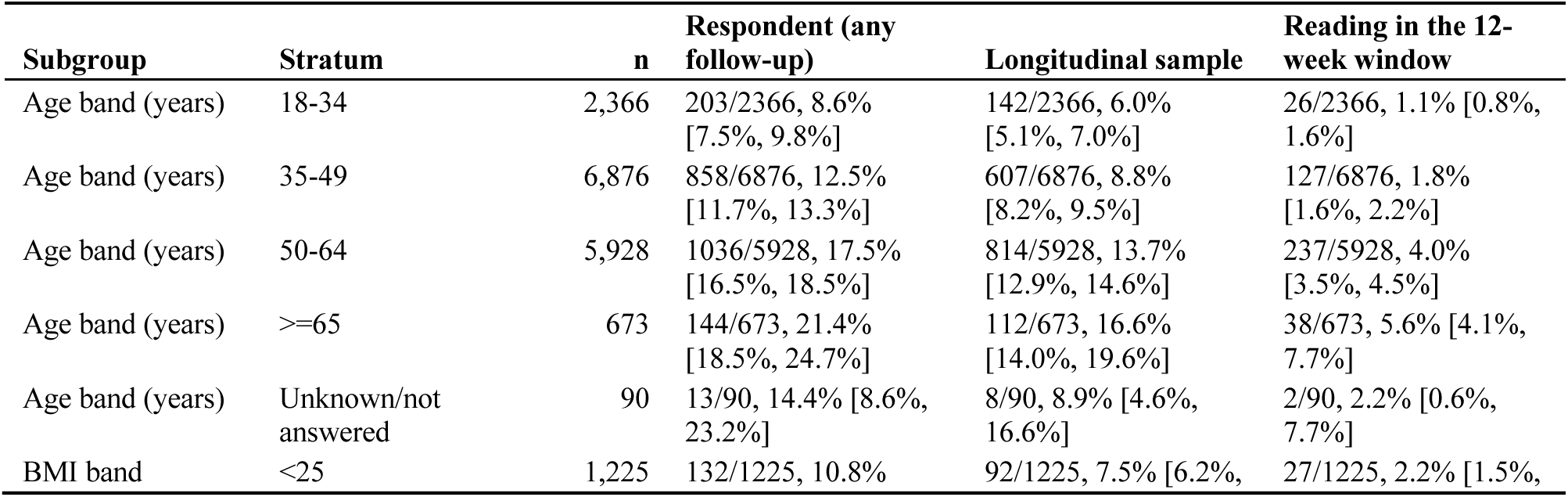
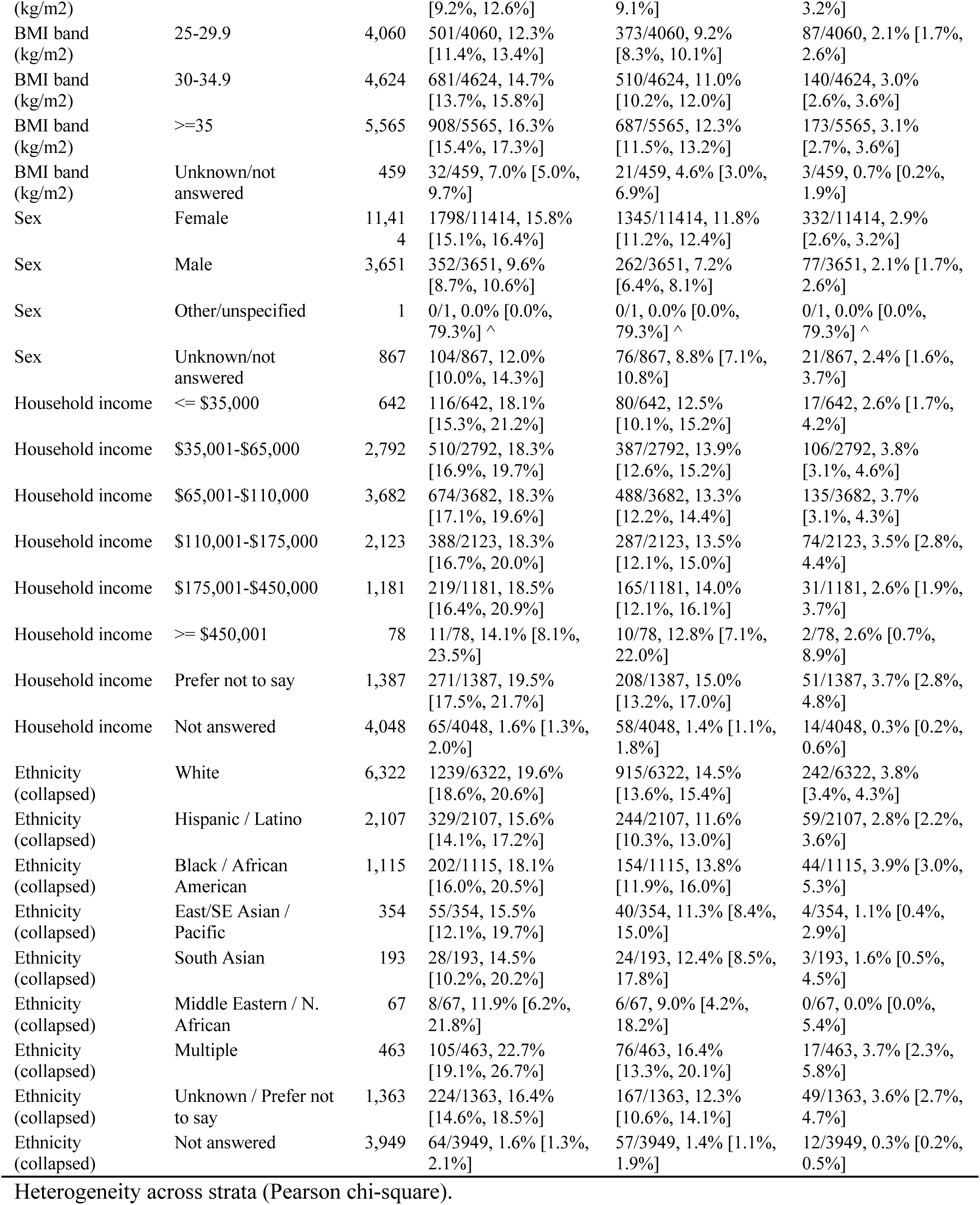
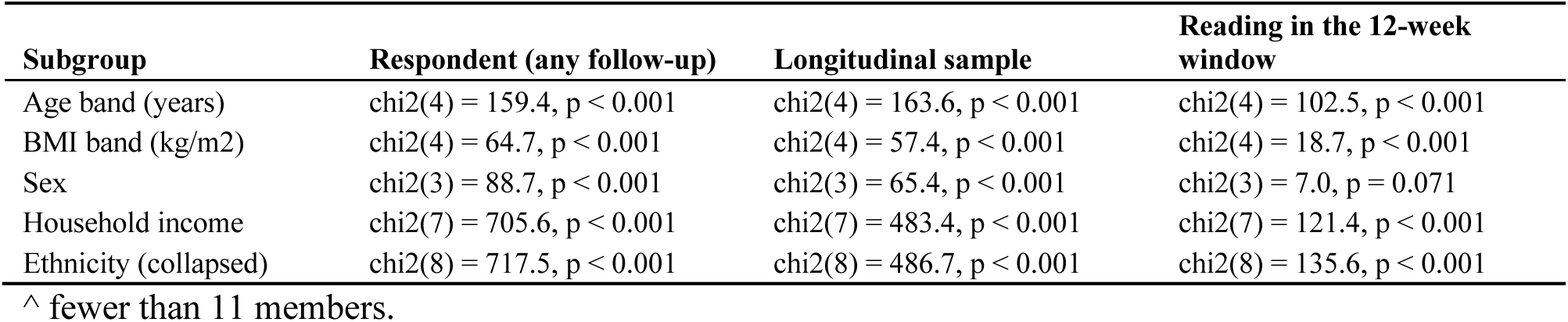
Follow-up by subgroup among the 15,933 members with a valid baseline: respondents (at least one follow-up reading of any timing), the longitudinal sample (at least one reading 14 days or later after baseline) and members with a reading in the 12-week window (days 70-98). Cells are n reaching the outcome / stratum n, rate [95% Wilson CI]. Heterogeneity is a Pearson chi-square test across the strata of each factor (last block). Unadjusted; exact counts, strata with fewer than 11 members are shown.

**TABLE S34.**
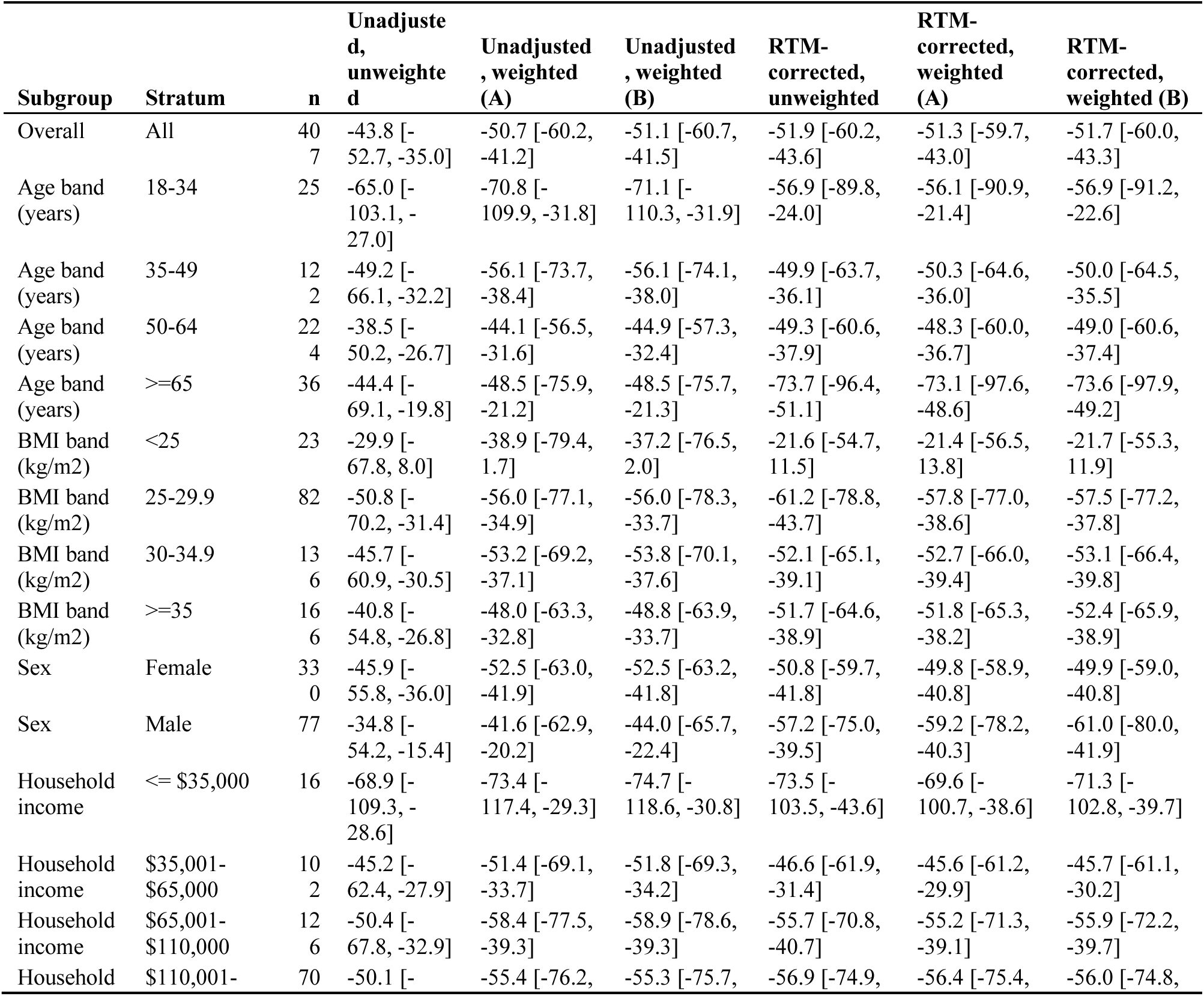
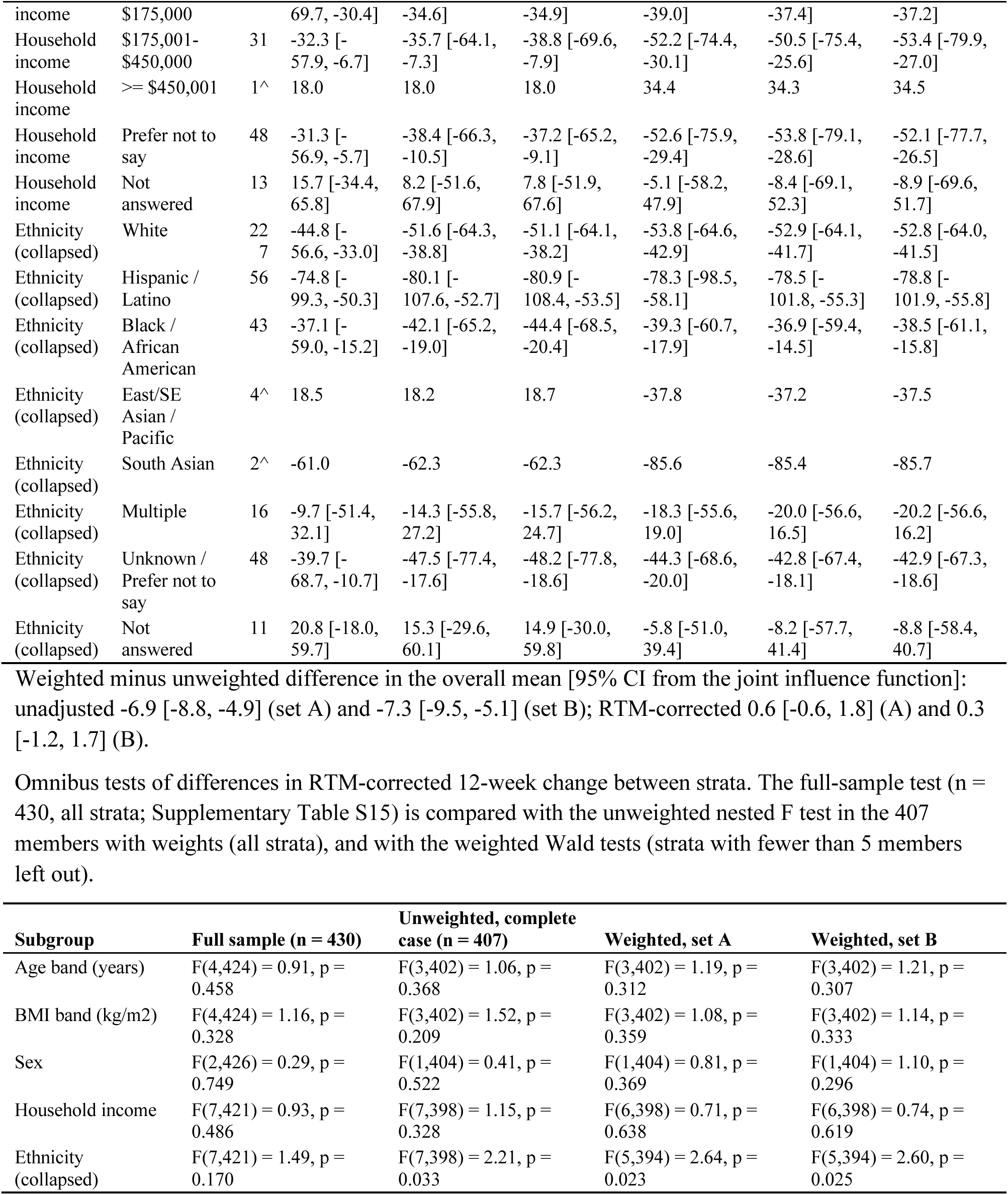
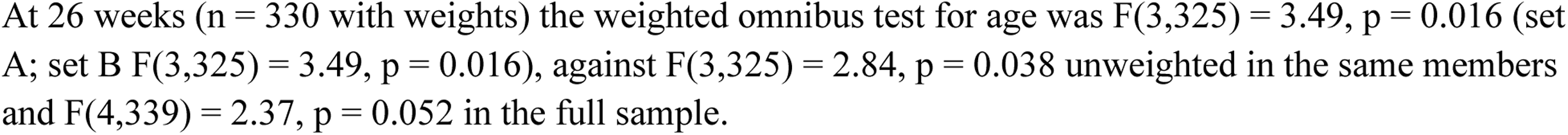
Inverse-probability-weighted and unweighted change in IBS-SSS at 12 weeks by subgroup (members with a reading in the window and complete age, body-mass index and sex, n = 407 of 430; the other 23 have no weight). Points; negative = improvement; mean change [95% CI]. Unweighted and weighted estimates use the same members; weights are the 12-week stabilised inverse-probability weights truncated at the 1st and 99th percentiles (set A: baseline score, age, BMI, sex; set B adds log(1 + pre-baseline engagement days)), applied by weighted least squares with HC1 intervals that do not propagate the weight-model error. RTM-corrected change is centred at 171.42. Strata with fewer than 5 members show point estimates only and are left out of the omnibus tests; ^ fewer than 11 members. Exploratory; no multiplicity correction.

**TABLE S35.** Responders and remission by baseline severity stratum and landmark. Cells are n/N = % [Wilson 95% CI]; no p value is given because none is defined for a proportion, and no multiplicity correction was applied. Observed, unadjusted proportions among members with a reading in the window: N differs by landmark, so columns are not one fixed group. Responder: baseline of 75 or more and a fall of 50 points or more. Remission: follow-up score below 75, defined at follow-up, so it is available for every baseline stratum above 75. Overall is each member’s first reading at 14 days or later (longitudinal sample); landmarks are 4 weeks (days 14 to 35), 12 weeks (days 70 to 98) and 26 weeks (days 154 to 210), using the reading closest to the target day. Members with a baseline below 75 cannot respond by definition; their N is shown. Moderate and severe combined is a descriptive subgroup (baseline 175 or above). The symptomatic row reproduces Table S6. Exploratory; association only.

| Baseline stratum | Overall (first reading at 14 days or later) | 4 weeks | 12 weeks | 26 weeks |
| --- | --- | --- | --- | --- |
| **Responder (fall of 50 points or more)** |  |  |  |  |
| Mild (75 to 174) | 226/551 = 41.0% [37.0, 45.2] | 31/93 = 33.3% [24.6, 43.4] | 64/153 = 41.8% [34.3, 49.8] | 58/118 = 49.2% [40.3, 58.1] |
| Moderate (175 to 299) | 336/566 = 59.4% [55.3, 63.3] | 67/130 = 51.5% [43.0, 60.0] | 77/133 = 57.9% [49.4, 65.9] | 86/117 = 73.5% [64.9, 80.7] |
| Severe (300 or above) | 158/222 = 71.2% [64.9, 76.7] | 41/56 = 73.2% [60.4, 83.0] | 28/37 = 75.7% [59.9, 86.6] | 28/38 = 73.7% [58.0, 85.0] |
| Moderate and severe combined (175 or above) | 494/788 = 62.7% [59.3, 66.0] | 108/186 = 58.1% [50.9, 64.9] | 105/170 = 61.8% [54.3, 68.7] | 114/155 = 73.5% [66.1, 79.9] |
| All symptomatic (75 or above) | 720/1,339 = 53.8% [51.1, 56.4] | 139/279 = 49.8% [44.0, 55.6] | 169/323 = 52.3% [46.9, 57.7] | 172/273 = 63.0% [57.1, 68.5] |
| Baseline below 75 (cannot respond) | not applicable (N = 344) | not applicable (N = 58) | not applicable (N = 107) | not applicable (N = 72) |
| **Remission (follow-up score below 75)** |  |  |  |  |
| Mild (75 to 174) | 231/551 = 41.9% [37.9, 46.1] | 33/93 = 35.5% [26.5, 45.6] | 67/153 = 43.8% [36.2, 51.7] | 54/118 = 45.8% [37.0, 54.7] |
| Moderate (175 to 299) | 124/566 = 21.9% [18.7, 25.5] | 24/130 = 18.5% [12.7, 26.0] | 30/133 = 22.6% [16.3, 30.4] | 27/117 = 23.1% [16.4, 31.5] |
| Severe (300 or above) | 16/222 = 7.2% [4.5, 11.4] | 4/56 = 7.1% [2.8, 17.0] | 4/37 = 10.8% [4.3, 24.7] | 5/38 = 13.2% [5.8, 27.3] |
| Moderate and severe combined (175 or above) | 140/788 = 17.8% [15.3, 20.6] | 28/186 = 15.1% [10.6, 20.9] | 34/170 = 20.0% [14.7, 26.6] | 32/155 = 20.6% [15.0, 27.7] |
| All symptomatic (75 or above) | 371/1,339 = 27.7% [25.4, 30.2] | 61/279 = 21.9% [17.4, 27.1] | 101/323 = 31.3% [26.5, 36.5] | 86/273 = 31.5% [26.3, 37.2] |

## Supplementary Figures

**FIGURE S1.**
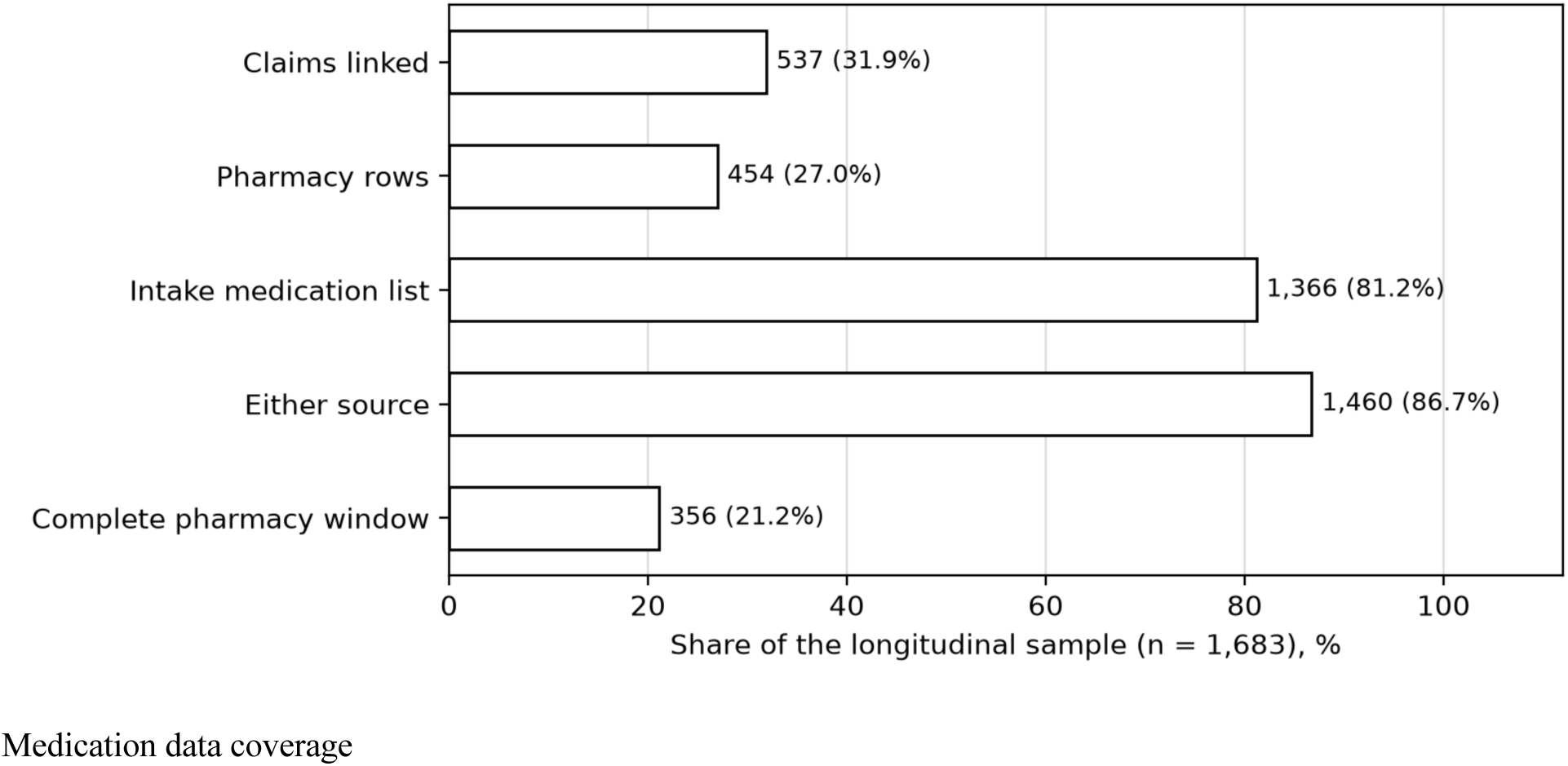
Medication data coverage in the longitudinal sample (n = 1,683): members with pharmacy claims linked (537), pharmacy rows (454), a baseline medication list (1,366), either source (1,460) and a pharmacy feed covering the 180 days before baseline through the last reading (356).

**FIGURE S2.**
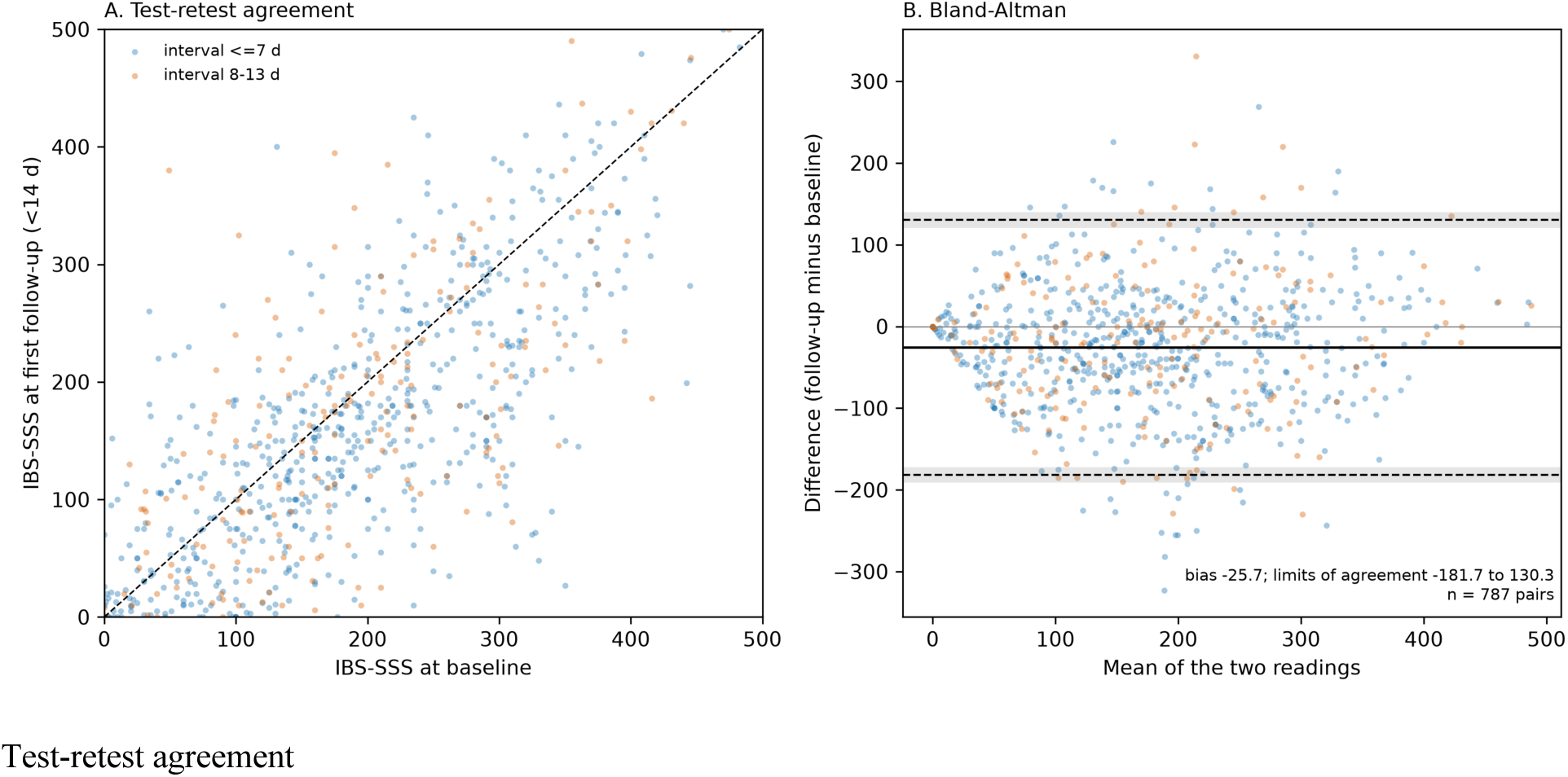
Test-retest agreement of IBS-SSS within 14 days (n = 787 members; 586 with an interval of 7 days or less, 201 with 8 to 13 days). Panel A, follow-up against baseline with the line of identity (dashed). Panel B, Bland-Altman plot; the solid line is the mean difference (–25.71; 95% CI –31.27 to – 20.14) and dashed lines are the 95% limits of agreement (–181.67 to 130.25). ICC(2,1) 0.709 (95% CI 0.668 to 0.748); Pearson r 0.729 (0.694 to 0.760).

**FIGURE S3.**
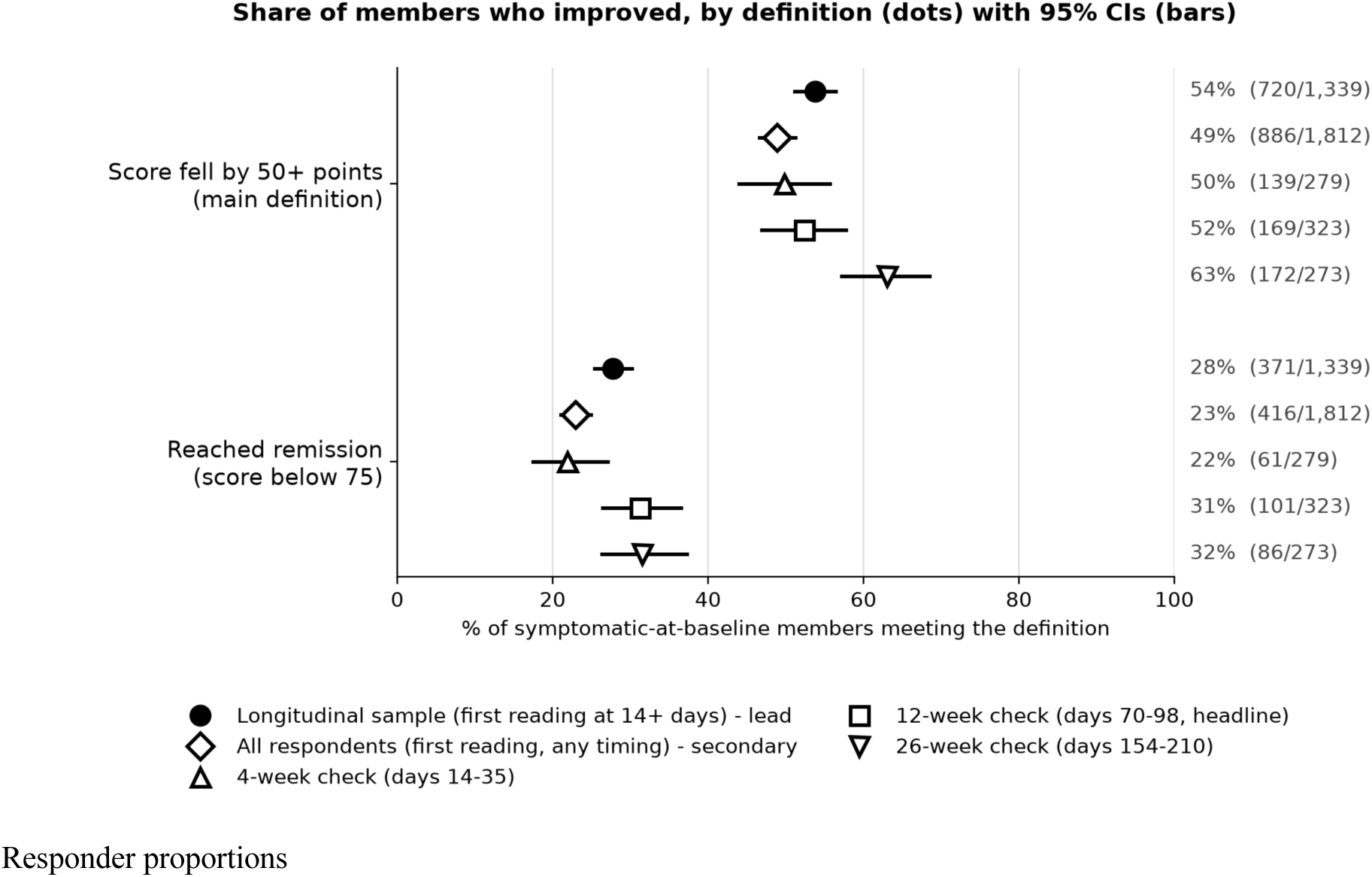
Responder proportions among symptomatic members (baseline 75 or more) by definition of response (50-point fall and remission). Dots are proportions with 95% Wilson CIs; numbers are responders/evaluable. Filled circles, longitudinal sample (n = 1,339); diamonds, all respondents with a first follow-up of any timing (n = 1,812); triangles and squares, 4-, 12- and 26-week landmarks (n = 279, 323, 273).

**FIGURE S4.**
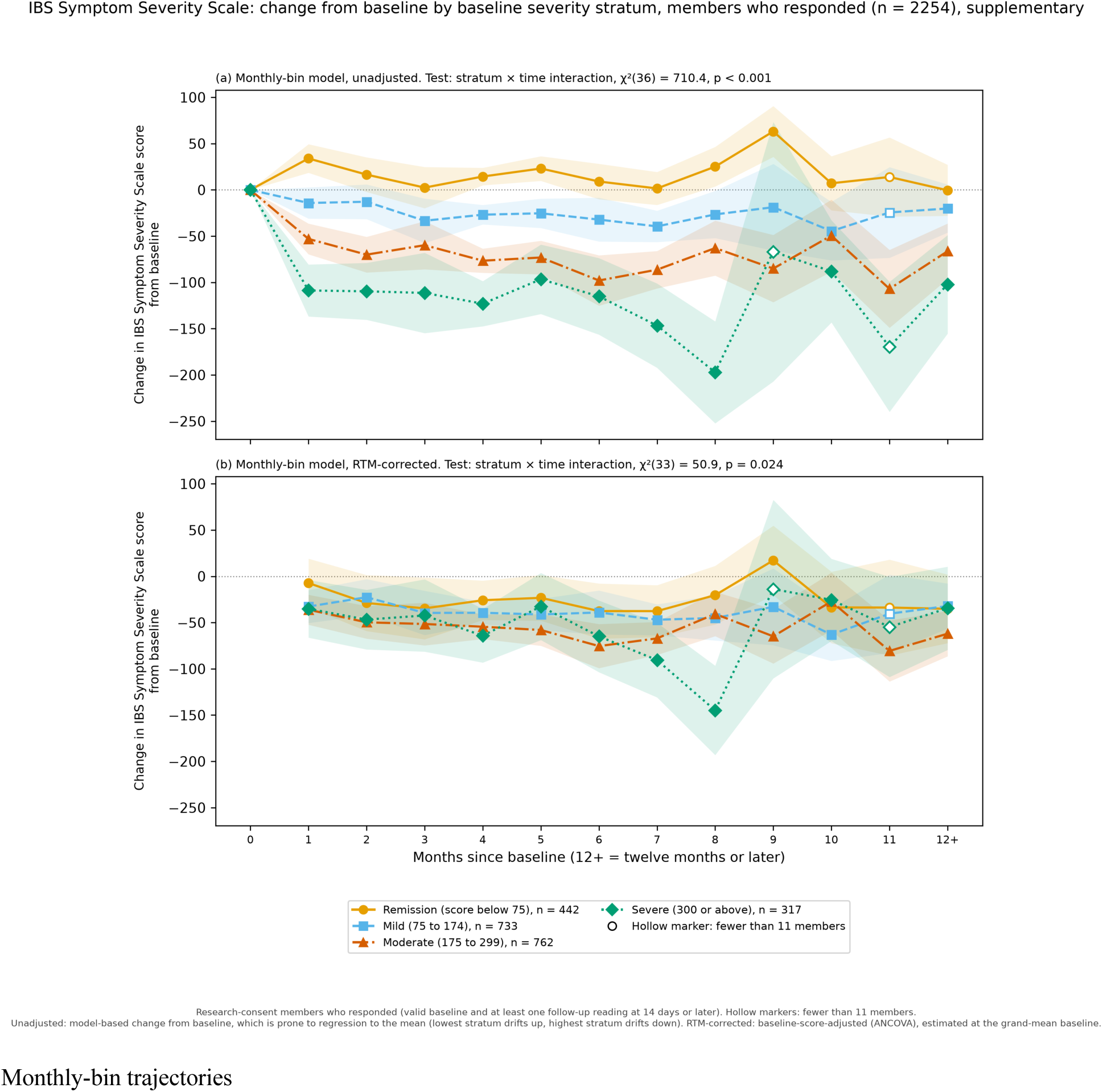
Monthly-bin trajectories of change in IBS-SSS by baseline severity stratum, unadjusted (panel a; 2,254 respondents, 4,596 rows; stratum-by-time chi-square(36) = 710.4, p < 0.001) and RTM-corrected (panel b; 1,683 members, 2,342 follow-up rows; chi-square(33) = 50.9, p = 0.024), with 95% CIs. Hollow markers indicate months with fewer than 11 members.

**FIGURE S5.**
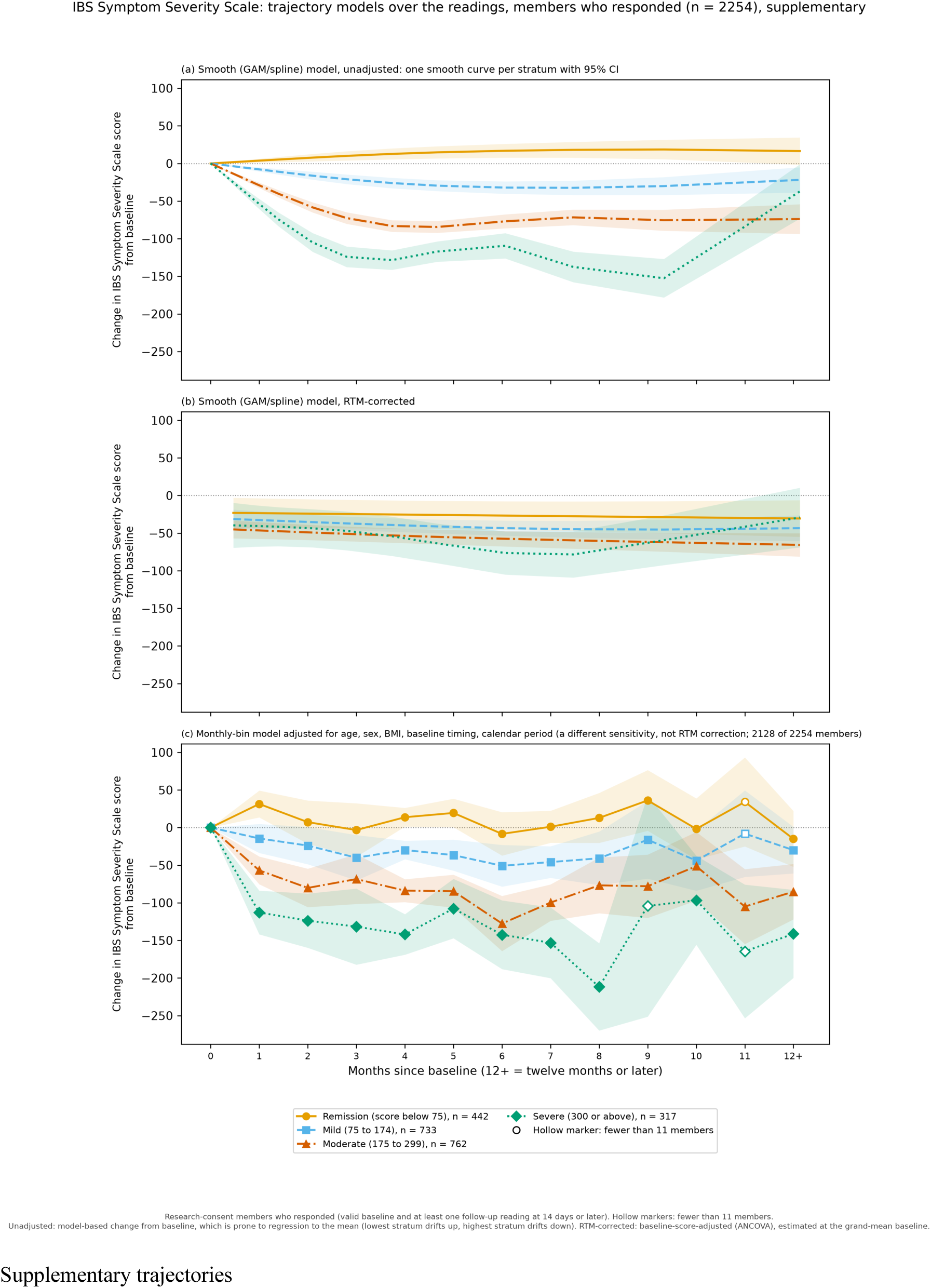
Smooth and covariate-extended trajectory models for IBS-SSS. Panels a and b, smooth additive mixed models, unadjusted and RTM-corrected; panel c, monthly-bin model additionally including age, sex, body-mass index, baseline timing and calendar period (2,128 of 2,254 members; a covariate sensitivity, not an RTM correction). Shaded areas are 95% CIs; hollow markers indicate months with fewer than 11 members.

**FIGURE S6.**
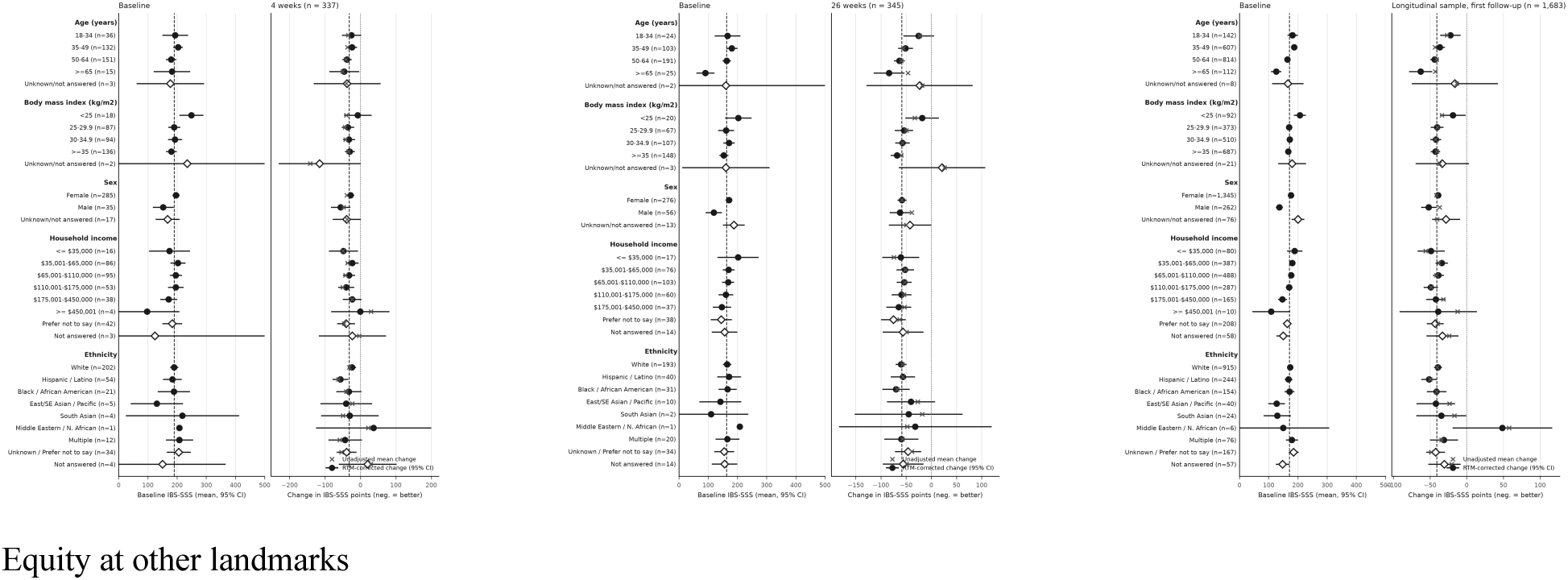
Change in IBS-SSS by subgroup at 4 weeks (n = 337), 26 weeks (n = 345) and pooled (n = 1,683). Conventions as in Figure 5; all panels are centred at the mean baseline of the longitudinal sample. Exploratory; no multiplicity correction.

**FIGURE S7.**
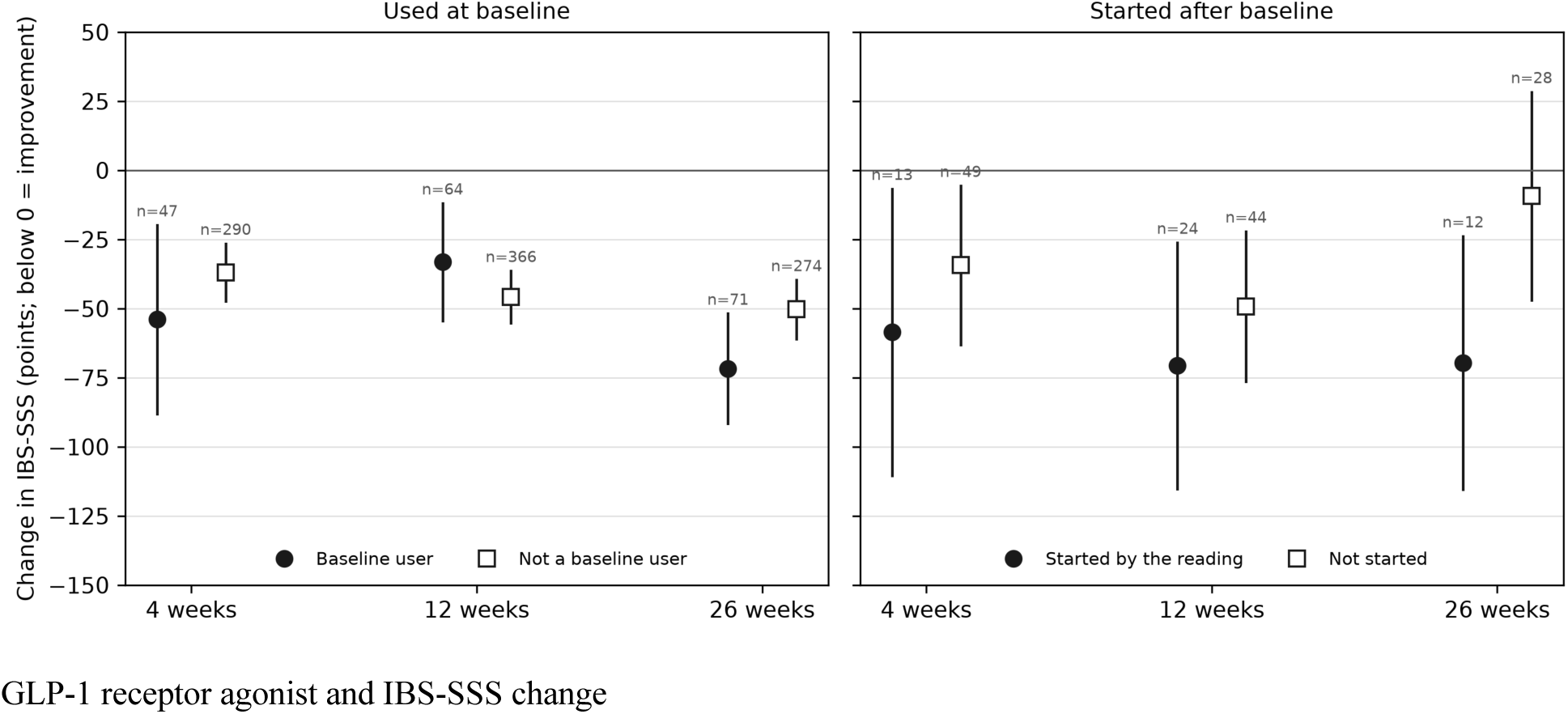
Change in IBS-SSS by GLP-1 receptor agonist status. Unadjusted mean change with 95% CIs at 4, 12 and 26 weeks, with exact n. Left, use at baseline; right, a start after baseline among members with a complete pharmacy window who were not baseline users. Associations only; medication data cover a minority of the sample.

**FIGURE S8.**
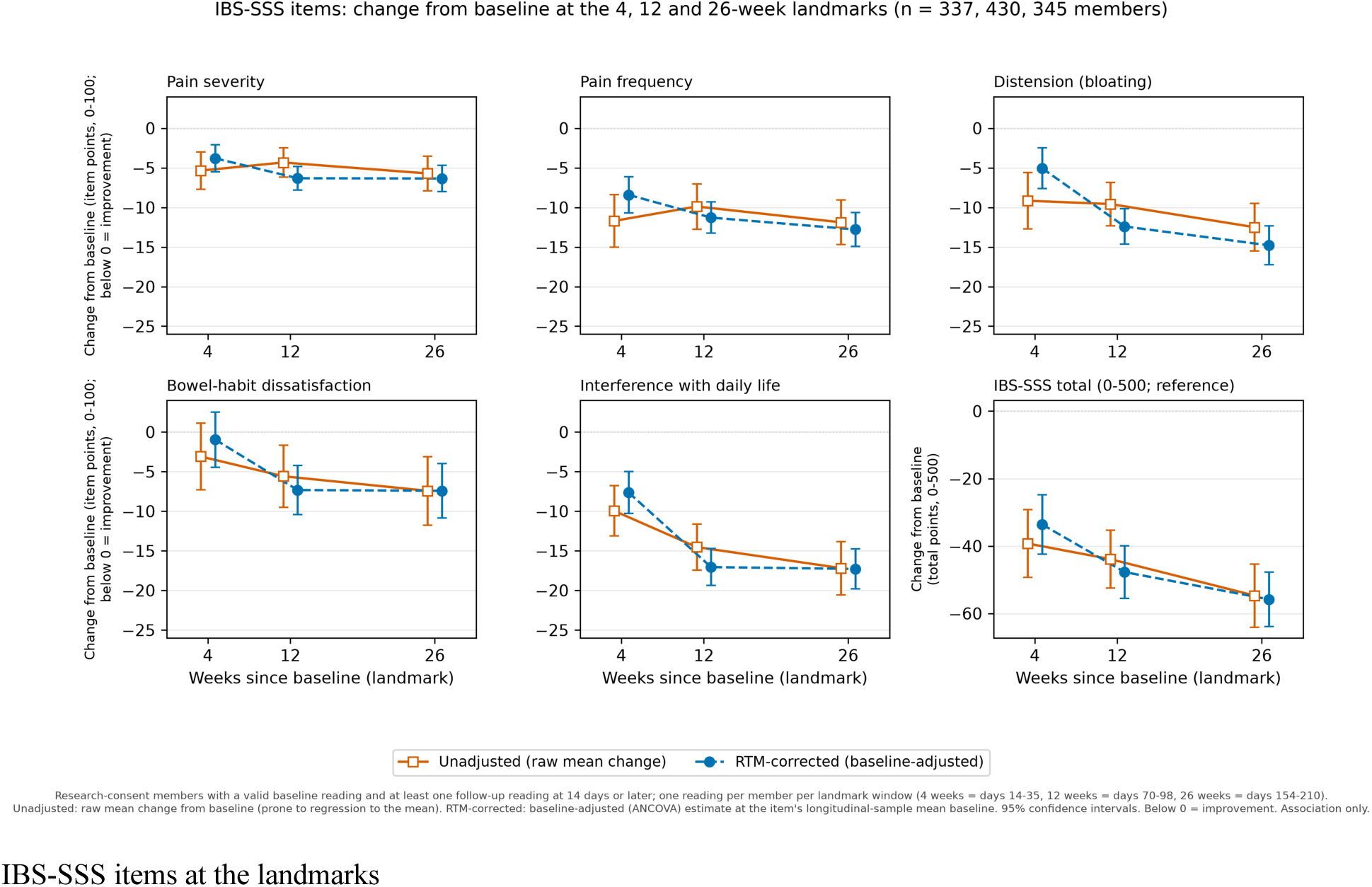
Change in IBS-SSS by item at 4, 12 and 26 weeks (n = 337, 430 and 345 members). Unadjusted raw mean change and RTM-corrected (baseline-adjusted) estimate with 95% CIs; items are scored 0-100 and the total (0-500) is shown for reference. Below 0 is improvement. Association only.

**FIGURE S9.**
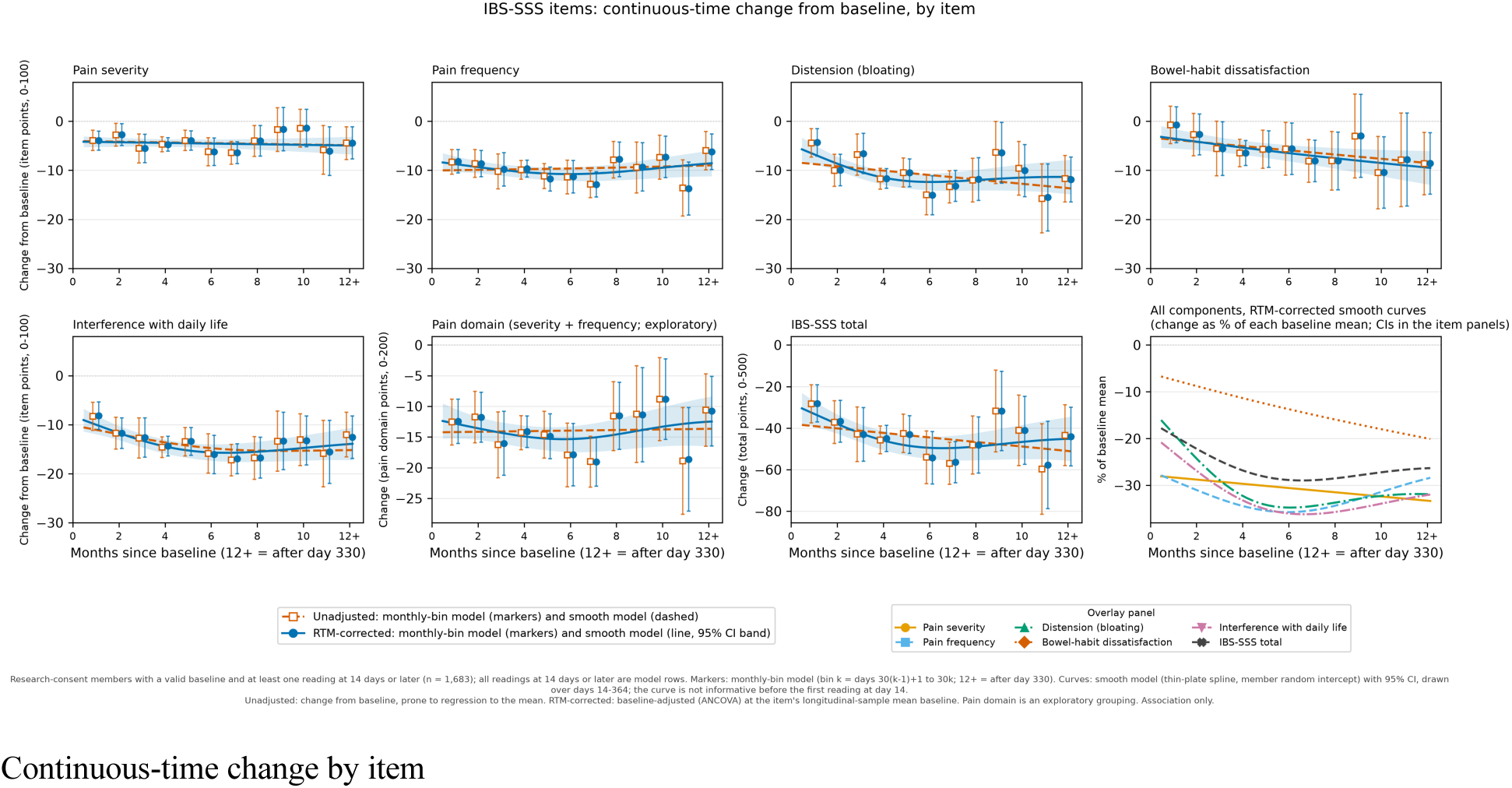
Continuous-time change from baseline by IBS-SSS item (1,683 members, 2,342 follow-up readings). Markers are monthly-bin model estimates and curves smooth-model estimates (RTM-corrected with 95% CI band, unadjusted dashed); the last panel overlays the RTM-corrected smooth curves as a percentage of each baseline mean. Curves are not informative before day 14 and are wide beyond about month 6. Exploratory; association only.

**FIGURE S10.**
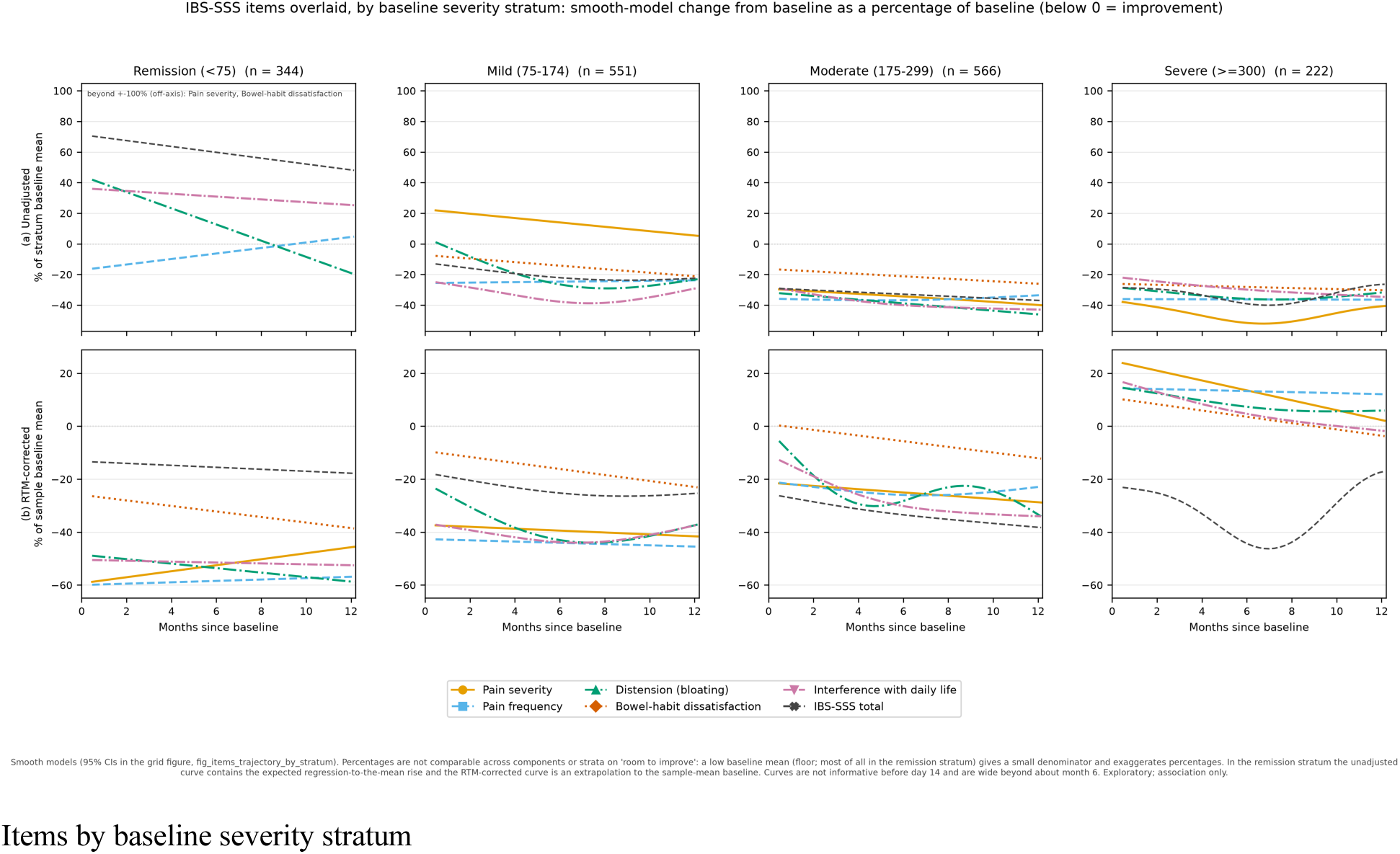
IBS-SSS items overlaid by baseline severity stratum: smooth-model change as a percentage of the stratum baseline mean (unadjusted, upper row) and of the sample baseline mean (RTM-corrected, lower row). Percentages are not comparable across items or strata because a low baseline mean (a floor, most of all in the remission stratum) gives a small denominator. In the remission stratum the unadjusted curve contains the expected regression-to-the-mean rise. Exploratory; association only.

